# The Metastatic Breast Cancer Project: leveraging patient-partnered research to expand the clinical and genomic landscape of metastatic breast cancer and accelerate discoveries

**DOI:** 10.1101/2023.06.07.23291117

**Authors:** Esha Jain, Jorge Gómez Tejeda Zañudo, Mary McGillicuddy, Daniel L. Abravanel, Beena S. Thomas, Dewey Kim, Sara Balch, John Navarro, Jakob H. Weiss, Tania G Hernandez, Michael Dunphy, Brett N. Tomson, Jorge Buendia-Buendia, Oyin Alao, Alyssa L. Damon, Simona Di Lascio, Shahrayz Shah, Ilan K. Small, Delia Sosa, Lauren Sterlin, Imani Boykin, Rachel E. Stoddard, Netsanet Tsegai, Ulcha F. Ulysse, Kolbe Phelps, Elizabeth Frank, Priti Kumari, Simone Maiwald, Katie Larkin, Sam Pollock, Andrew Zimmer, Parker S. Chastain, Taylor Cusher, Colleen Nguyen, Sarah Winnicki, Elana Anastasio, Eliezer M. Van Allen, Eric S. Lander, Todd R. Golub, Corrie A. Painter, Nikhil Wagle

## Abstract

Capturing the full complexity of the clinical experiences of metastatic breast cancer (MBC) patients treated in a variety of settings is needed to better understand this disease and develop new treatment modalities. Yet, challenges exist to establish and share a large MBC dataset that integrates genomic, clinical, and patient-reported data as it requires collecting information and samples from many geographically dispersed patients and institutions. We explored whether a patient-partnered research approach that uses online engagement could enable patients living across the United States and Canada to accelerate cancer research by sharing their samples, clinical information, and experiences. In collaboration with patients and patient advocates, the Metastatic Breast Cancer Project (MBCproject; www.mbcproject.org) was developed and launched in October 2015. As of March 2020, 3,246 MBC patients who received treatment at ∼1,700 institutions had consented for the MBCproject, providing patient-reported information via surveys, as well as access to medical records and biological samples. Through the collection and analysis of tumor and germline samples, medical records, and patient-reported data, the MBCproject generates and publicly releases clinically-annotated genomic data on primary and metastatic tumor specimens on a recurring basis.

Herein we describe the MBCproject cohort in detail and describe the clinico-genomic landscape of the MBCproject dataset. The complete dataset consists of whole exome sequencing (WES) for 379 tumors with matching germline from 301 patients, WES on germline samples from 377 patients, and transcriptome sequencing (RNA-seq) for 200 tumors from 141 patients, with clinical data from medical records and patient-reported information. A comparison of various clinical fields (diagnostic dates, tumor histology, tumor sites, treatments received) obtained from patient-reported data and the abstracted from medical records found a high degree of concordance, with multiple fields having over 90% concordance. Analysis of the somatic alterations in the 249 tumors taken after metastatic diagnosis found a significant enrichment of mutations in the cancer genes *TP53*, *PIK3CA*, *CDH1*, *PTEN, AKT1, NF1*, and *ESR1*, among others. Tumor evolutionary analysis of 14 patients with 3 or more samples identified oncogenic mutations in *ESR1*, *NF1*, and *TP53*, genes associated with MBC and/or resistance to endocrine therapy. Analysis of germline samples identified pathogenic variants in the cancer-associated genes *BRCA1, BRCA2*, *ATM,* and *PALB2*. Comparing the frequency of pathogenic variants in patients diagnosed before/at or after the age of 40 years old, we found that the presence of these variants in *BRCA1* or *BRCA2* was enriched in the younger group compared to the older group (9.2% vs 2.5%, p=0.0089; two-sided Fisher exact test). Transcriptome sequencing identified putatively oncogenic in-frame fusions in cancer genes such as *FANCD2*, *FGFR3*, *ESR1*, *BRAF* and *NCOR1*. Analysis of tumor’s intrinsic molecular subtype (research-based PAM50) found a depletion of the Luminal A subtype in MBCproject compared to The Cancer Genome Atlas, and a switch in molecular subtype in 15 out of 35 patients with 2 or more samples. A case study of a patient with sequencing data from 4 tumor biopsies obtained during the course of their metastatic disease is presented. An integrated analysis of the clinical and multi-omic data from this patient identified distinct drivers of resistance to endocrine therapy in each of these tumors.

The MBCproject clinico-genomic dataset is one of the largest available MBC patient cohorts This integrated dataset is poised for studying several understudied clinical cohorts (young women with breast cancer, *de novo* MBC), rare disease subtypes (e.g. lobular, metaplastic, extraordinary responders), biomarkers of response/resistance (e.g. CDK4/6 inhibitors), and real world patterns, among others, and will serve as an invaluable resource to accelerate discoveries.

## Introduction

Breast cancer is the most commonly diagnosed cancer among women globally and the second leading cause of cancer death among women in the United States^1, 2^. Multiple large scale studies have characterized the genomics of breast cancer and its disease subtypes and have identified several recurring genomic drivers that can be targeted clinically^3–5, 6–9^. However, most of these studies have focused on primary breast cancers, are often conducted by a single institution, and are limited to studying patient cohorts that receive cancer care at major cancer centers ^6, 7, 10–15^. This traditional approach of conducting research has potentially resulted in a skewed characterization of metastatic breast cancer (MBC) that does not capture the full range and diversity of cancer patients, as has been observed in other cancers^16–18^, and an unmet need to correct this.

To better capture the full range and diversity of disease experiences of metastatic breast cancer patients treated in a variety of clinical settings, an accessible approach that is available to patients regardless of where they live is required. We explored whether an approach of directly partnering with patients online to create a nationwide research project could overcome these challenges. Harnessing the power of social media and online communities that allow people to connect irrespective of geography, we engaged with members of the metastatic breast cancer community to listen to their experiences and understand their specific needs and perspectives. These initial relationships established with patients, advocates, and advocacy groups led to extensive discussions to frame and shape this new model of patient-partnered research, which has ultimately proven to be a successful research modality that has led to scientific discoveries with clinical impact in multiple cancer types^18, 19^.

Described herein, the resulting Metastatic Breast Cancer Project (MBCproject) was built and launched with deep patient involvement. By collecting biological samples (saliva, blood, and/or archived tumor tissue), medical records, and patient-provided information from surveys, the MBCproject generated an integrated and publicly available dataset of genomic (whole exome sequencing and RNA sequencing), clinical, and patient-reported information that is described and analyzed throughout this work.

## Results

### Building a patient-partnered project for metastatic breast cancer

In order to enhance the current understanding of the genomic and clinical landscape of metastatic breast cancer, we launched a patient-partnered research project that is facilitated by an online platform (https://mbcproject.org/), which allows for both online registration and consent (Fig. 1A). As a result, metastatic breast cancer patients living anywhere in the US and Canada can enroll in this study and provide informed consent online to share access to their samples, medical records, and information about their cancer experience. The study was launched in October 2015, with over 2600 patients registering and over 1500 patients providing consent to participate in MBCproject within the first year of the project’s launch (Fig. 1D). Much of the project’s early growth was attributed to patients and patient advocates sharing information about the project via social media platforms (Fig. 1D, Supp. Fig. S2). As of March 31st 2020, 3246 patients who have received medical care at over 1690 institutions across USA and Canada had registered and consented to participate in the study (Fig. 1B-C). This project was developed with input and in collaboration with the metastatic breast cancer community, including patients and patient advocates (Supp. Table S1, Supp. Fig. S1A-B) and continues to evolve with their input.

**Figure 1.**
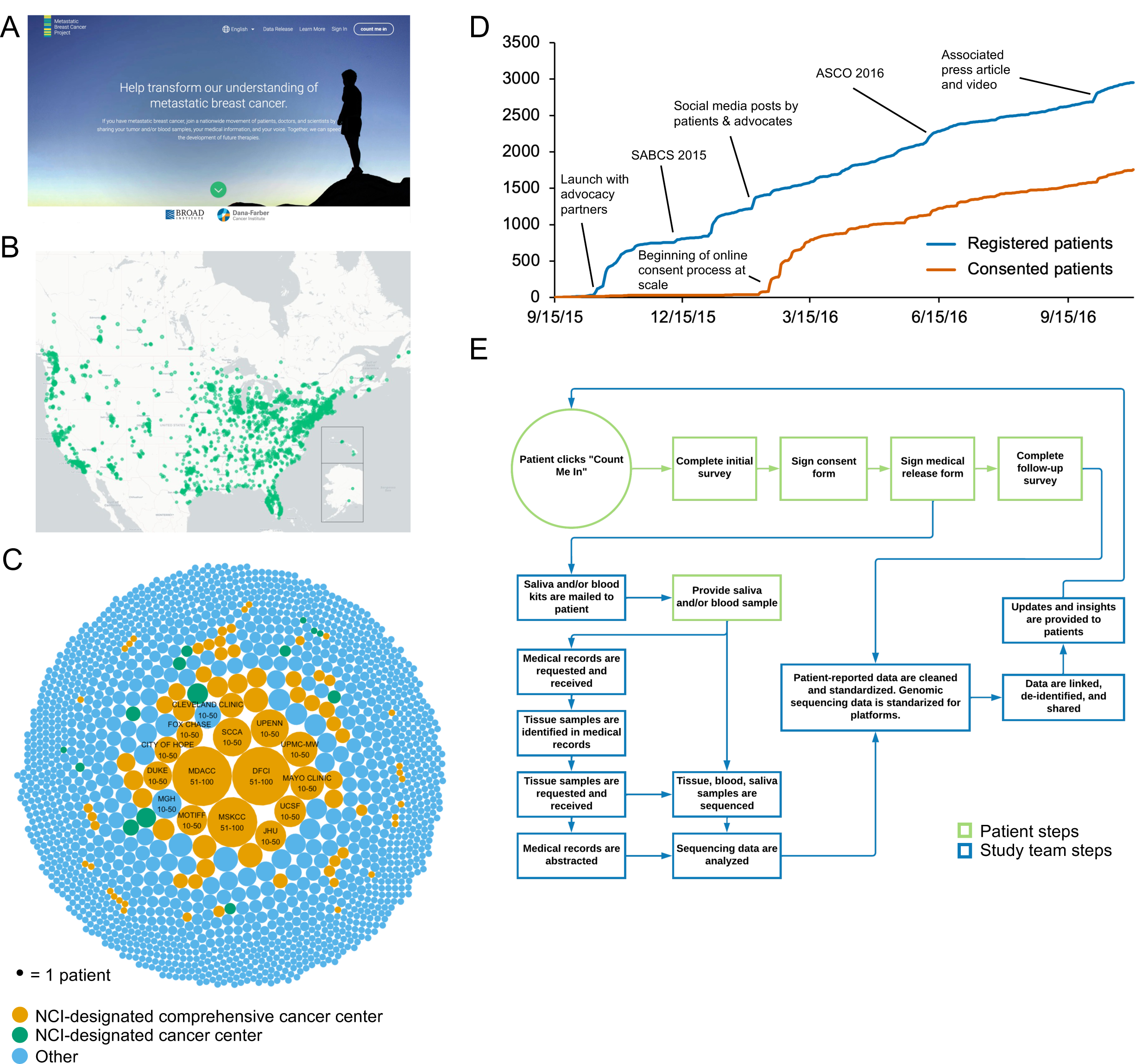
Building a patient-partnered project for metastatic breast cancer (N=3246). (A) MBCproject.org homepage, designed in partnership with patients. (B) Geographical distribution of the 3246 patients who provided consent for participation in the MBCproject between September 15, 2015 and March 31, 2020. (C) Medical institutions where participants reported receiving care for their breast cancer. Patients reported receiving care from over 1,600 distinct United States medical institutions. Each unique medical center is represented by a circle. The size of the circle corresponds with the number of patients who reported receiving care at said institution. Around 6% of reported medical institutions are National Cancer Institute (NCI) designated comprehensive cancer centers (denoted by an orange circle) or NCI designated cancer centers (denoted as a green circle). 1174 (∼69%) medical institutions were reported by only one patient each. (D) Cumulative number of patients registered (blue) and consented (red). (E) Overview of MBCproject processes. Steps outlined in green represent patient steps and steps outlined in blue represent study team steps. MBC, Metastatic Breast Cancer; MBCproject, The Metastatic Breast Cancer Project.

Patients provided online consent for participation in the MBCproject, which allowed for the acquisition of medical records and biological samples (tumor, saliva and/or blood), the analysis of WES from matched tumor and germline samples and of RNA-Seq from tumors, and the sharing of de-identified patient-reported, clinical, and genomic data on public databases multiple times (Table 1, Fig. 1E). Patients continue to be engaged with MBCproject and are provided with regular study updates via email newsletters and social media. (Fig. 1E, Supp. Fig. S1A-B).

**Table 1.** Number of requested, received and percentage yield of consent forms, medical records, saliva, tissue and blood samples collected for the MBCproject study. MBCproject, The Metastatic Breast Cancer Project.

| **Data type** | **Requested** | **Received** | **Yield** |
| --- | --- | --- | --- |
| **Dataset freeze (March 31, 2020)** |  |  |  |
| Consent Form | 4952 | 3246 | 66% |
| Saliva Sample | 2830 | 2092 | 74% |
| Medical Record | 2196 | 1360 | 62% |
| Tumor Sample | 852 | 608 | 71% |
| Blood (cfDNA) | 1463 | 1112 | 76% |
| **Current (September 30, 2022)** |  |  |  |
| Consent Form | 5537 | 3684 | 67% |
| Saliva Sample | 3182 | 2236 | 70% |
| Medical Record | 2208 | 1372 | 62% |
| Tumor Sample | 1114 | 703 | 63% |
| Blood (cfDNA) | 1464 | 1114 | 76% |

### Generating a clinco-genomic dataset for the MBCproject

The MBCproject (as of March 31st 2020) is comprised of 3246 patients who have provided their consent to participate in the study. All of these patients (N = 3246) provided self-reported information about their demographics and disease characteristics by responding to questions in the intake survey (Table 2, Suppl. File 1). A subset of these patients (N = 1604) provided additional information through a follow-up survey (Table 2, Supp. File 1). The study team requested (N = 2196 patients) and received (N = 1360 patients from over 650 institutions, yield = 62%) medical records, sent (N = 2830 patients) and received saliva kits (N = 2092 patients, yield = 74%) for the collection of germline DNA, sent requests (N = 852 samples) and received tumor tissue (N = 608 samples, yield = 71%) for the collection of tumor DNA, and sent (N = 1463 patients) and received blood kits (N = 1112 patients, yield = 76%) for the collection of germline blood and cell-free DNA (cfDNA) (Table 1, Supp. Fig. S3A). cfDNA samples (N = 954) underwent ultra low pass whole genome sequencing (ULP-WGS) to estimate the tumor content in these samples. The cfDNA samples with sufficient tumor content (N = 144) were sent for WES. Among the tumors that underwent WES, a total of 379 tumors (275 tissue biopsies, 104 circulating tumor DNA (ctDNA) samples) from 301 patients had a tumor purity of >=10% (Fig. 2, Supp. Fig. S3A) and were included in the final WES dataset for downstream analysis. Detailed clinical information was abstracted for these 301 patients from their medical and pathology records. Additionally, 228 tumor biopsies were submitted for RNA sequencing, of which 200 samples (141 patients) passed quality control and were included in the downstream analysis, with 157 samples (120 patients) having both WES and RNA-seq data. WES from germline samples without matching tumor WES were included in the downstream analysis (377 patients). Overall, the cohort is composed of genomic and transcriptomic data from somatic or germline samples from 379 unique patients with metastatic breast cancer. Throughout this work, we refer to the clinical and multi-omic data associated with the 379 tumors (from tissue or ctDNA) and 301 patients with WES as the MBCproject clinico-genomic dataset.

**Figure 2.**
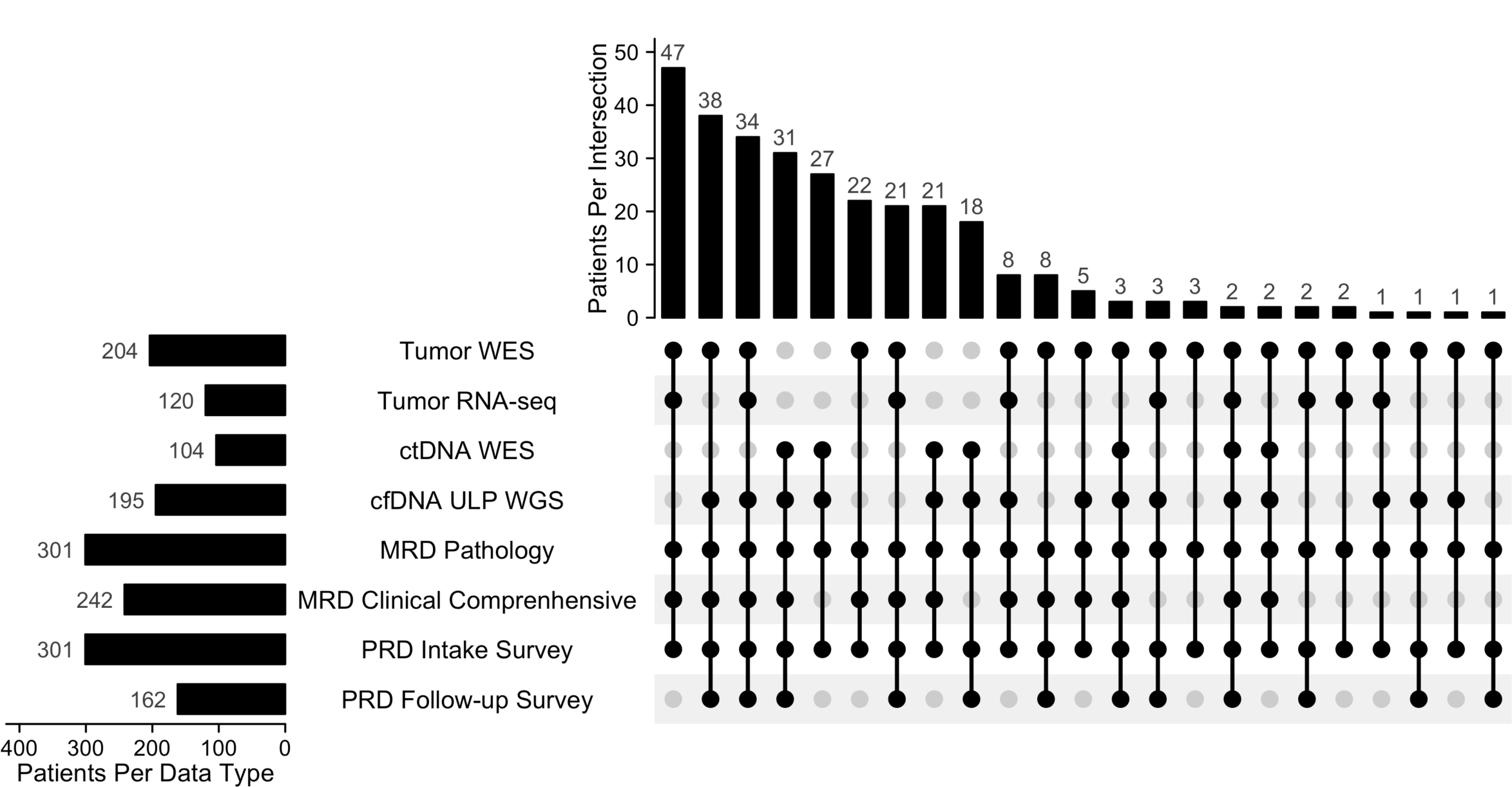
Multi-omic and clinical data types collected for the Metastatic Breast Cancer Project. Data types generated and number of patients with each data type for the MBCproject clinico-genomic dataset (N=379 tumor, 301 patients). MBC, Metastatic Breast Cancer; MBCproject, The Metastatic Breast Cancer Project; WES, Whole Exome Sequencing.

**Table 2.** Clinical data collection, biospecimen acquisition, and genomic data generation carried out to build the MBCproject dataset. MBCproject, The Metastatic Breast Cancer Project.

| **Data type** | **Number of patients or samples** |
| --- | --- |
| **MBCproject patient data collection** |  |
| Consent form signed | 3246 patients |
| Survey #1 submitted<br>(Intake survey of demographics, diagnosis details, receptor status, clinical experiences) | 3246 patients |
| Survey #2 submitted<br>(Follow-up survey of pathology details, metastatic sites, treatment types and timing) | 1604 patients |
| Medical record received (sufficient for full clinical review) | 1360 patients from 650 medical institutions |
| Saliva sample received | 2092 patients |
| Blood sample received | 1112 patients |
| Tumor sample(s) received | 608 samples (383 patients) |
| **MBCproject data generation** |  |
| Germline samples with completed WES | 391 germline samples |
| Tumor samples with completed WES | 391 tumor samples |
| Tumor samples with completed RNA-seq | 228 tumor samples |
| Tumor samples with corresponding H&E slide | 608 tumor samples |
| Cell-free DNA samples with completed ULP WGS | 955 blood samples |
| Circulating tumor DNA samples with completed WES | 144 blood samples |
| Pts with WES and medical records fully abstracted | 259 patients |

### Clinical characteristics of the MBCproject through patient reported data

We used patient reported data (PRD) to describe the patient, disease, and treatment characteristics of the MBCproject study participants. Patients shared information about their disease characteristics and clinical history (Fig. 3A-C, Suppl. Table S2) and their demographics (Suppl. Table S3) through an intake survey. A subset of patients (N = 1604, Table 2) provided information regarding disease histology, including rare subtypes (Suppl. Table S2), sites of metastasis (Fig. 3D), and treatments received (Fig. 3E-F) through a follow-up survey. The median age of primary breast cancer diagnosis of patients in the study was 45 yrs (mean - 45.8 yrs, range - 20-75 yrs), including 1077 patients (33.2%) who were diagnosed with breast cancer at 40 yrs of age or younger (Fig. 3A). Collectively, the study includes patients newly diagnosed with metastatic disease as well as long term survivors, with the time from primary to metastatic diagnosis ranging from 0 to > 11 yrs (median: 27.0 months, Fig. 3B). The time between metastatic diagnosis and registration in the study ranged between ≤1 to >10 yrs (median: 1.48 yrs, Fig. 3C).

**Figure 3.**
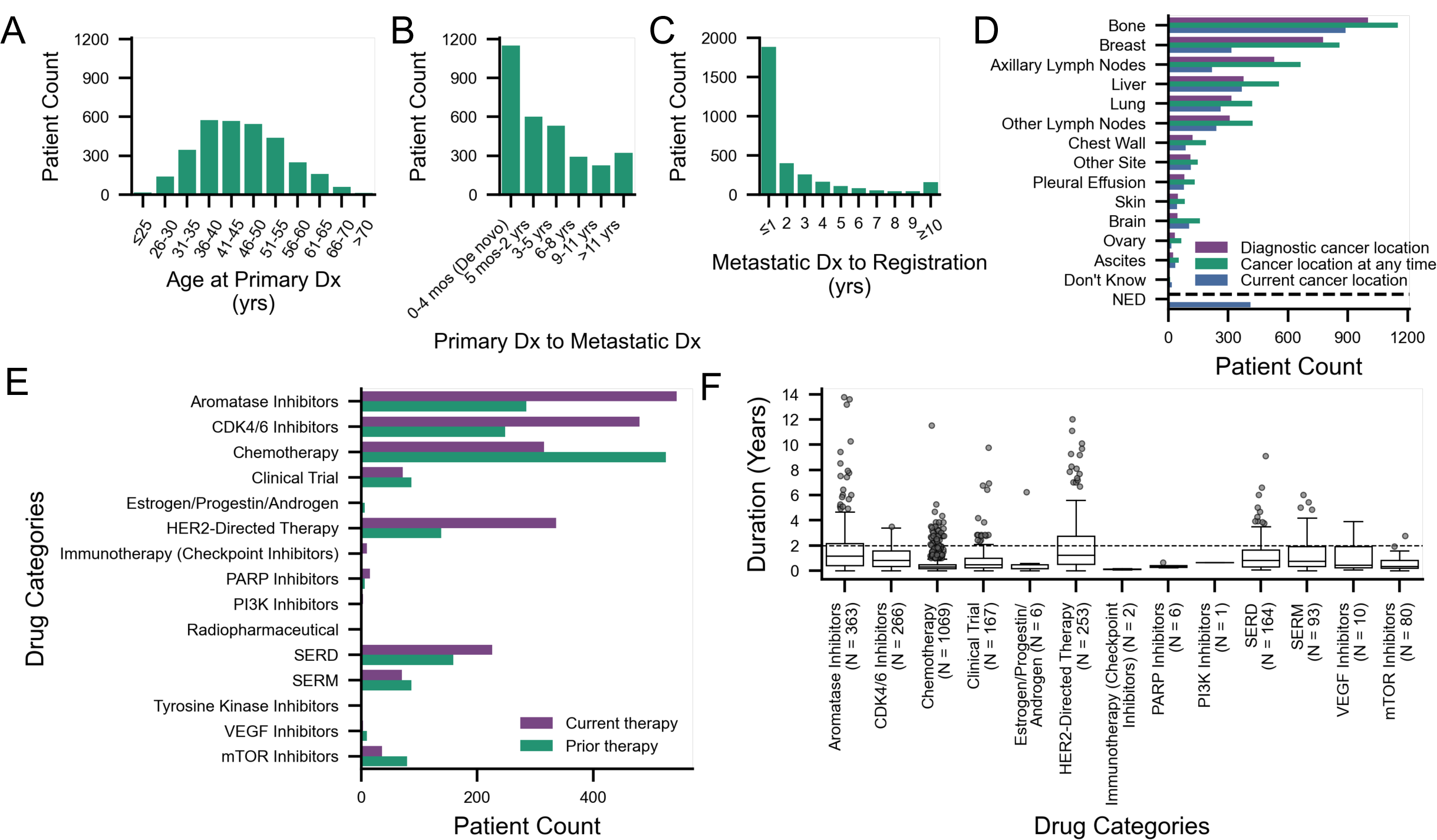
Clinical attributes from patient-reported data in the Metastatic Breast Cancer Project (N=3246 patients). Patient-reported data from Metastatic Breast Cancer Project participants obtained from an intake and follow-up survey. (A) Distribution of age at primary breast cancer diagnosis (N=3110). (B) Time between date of primary and metastatic diagnosis (N=3122). (C) Time between metastatic breast cancer diagnosis and registration for the MBCproject (N=3204). (D) Distribution of sites of metastatic breast cancer (N=1673). Patients selected location(s) of their cancer at the time of metastatic breast cancer diagnosis (purple), at time of survey completion (blue), and at any time during their experience with metastatic breast cancer (green). Patients could select more than one option. (E) Metastatic breast cancer treatment information for therapies received before (prior therapy, green) or at the time of survey completion (current therapy, purple) (N=1451). (F) Treatment duration for prior therapies received (N=839). Each data point denotes a treatment regimen reported by a patient. The horizontal dotted line denotes duration of 2 or more years. NED, No evidence of disease; MBC, Metastatic Breast Cancer; MBCproject, The Metastatic Breast Cancer Project.

Patients reported breast (diagnostic: 777, anytime: 859, current: 316), axillary lymph nodes (diagnostic: 531, anytime: 664, current: 218), bone (diagnostic: 1002, anytime: 1152, current: 888), and liver (diagnostic: 377, anytime: 556, current: 369) as the top 4 most frequent sites of metastases at diagnosis or at any time during their disease, as well as other distant metastatic locations (Fig. 3D). Notably, 411 patients self-reported as having no evidence of disease (NED) at the time of MBCproject survey submission. Treatment information from the follow-up survey (N = 1511 patients) was used to identify major treatment categories received by patients in this study (Suppl. Table S4). The top 5 most common prior or current therapies received were aromatase inhibitors (current: 544, prior: 285), CDK4/6 inhibitors (current: 480, prior: 248), HER2-directed therapies (current: 336, prior: 138), chemotherapy (current: 315, prior: 525), and SERDs (current: 226, prior: 159). Recently approved treatment regimens like immunotherapy (current: 10, prior: 2) and PI3K inhibitors (current: 3, prior: 1) were also reported (Fig. 3E). Evaluating prior treatments given under the metastatic setting and their associated treatment duration (N = 839 patients), we identified patients that exhibited extraordinary responses (treatment duration ≥ 2 years) to various drug categories. Overall, 31.1% (N = 261) of patients experienced an extraordinary response to a treatment, which spanned among various drug categories including aromatase inhibitors, SERDs, SERMs, CDK4/6 inhibitors, HER2-directed therapies, and clinical trials, among others (Fig. 3F).

### Clinical characteristics of the MBCproject through medical record abstraction

We analyzed the abstracted clinical and pathology records from medical record data (MRD) for the 379 tumor samples (301 patients) from the MBCproject clinico-genomic dataset (Suppl. Fig. S3). The age of diagnosis with breast cancer ranged from <25 to >65 years, with a median age of 44 years (Suppl. Fig. S4A). The median interval between primary and metastatic diagnosis was 24.5 months (range = 0 - 343 months, Suppl. Fig. S4B). The median interval between patient’s primary diagnosis and time of biopsy collection was 1.5 months (range = −0.7 - 408.3 months, Suppl. Fig. S4C). Of the 379 tumor biopsies, 73% (N = 275 samples, 204 patients) were collected from archived tissue (from 18 unique anatomical locations) and the remaining 27% (N = 104 samples, 104 patients) were collected from ctDNA (Fig. 4A, F). Biopsies were collected both before metastatic disease diagnosis (N = 114 samples, 30.1%, 91 patients) and after metastatic disease diagnosis (N= 249 samples, 65.7%, 209 patients) (Fig. 4B). 45.9% (N = 174) of the biopsies collected were treatment naive (not exposed to any breast cancer related treatment), while 33.8% (N=128) of the tumors were collected after treatment exposure (Fig. 4C). The number of tumor biopsies and timepoints per patient ranged between 1 and 5, with 56 patients having more than 1 tumor biopsy (Suppl. Fig. S3B, Suppl. Table S6A) and 49 patients having tumor biopsies from more than 1 timepoint (Suppl. Fig. S3B, Suppl. Table S6B). Additional information was abstracted for each tumor biopsy which included tumor histology (IDC, N = 167, 44.1%; ILC, N = 30, 7.9%; Mixed, N = 26, 6.9%), receptor status (HR+/HER2-, N = 78, 20.6%; HR+/HER2+, N = 27, 7.1%; HR-/HER2+, N = 10, 2.6%; TNBC, N=10, 2.6%) (Fig. 4D-E, Suppl. Fig. S5A-C), and tumor location/site (Fig. 4F-G). Of the cancer sites of metastases, bone (metastatic diagnosis: 164, anytime: 196, primary diagnosis: 1) and liver (metastatic diagnosis: 65, anytime: 101, primary diagnosis: 0) were the two most frequent sites (Fig. 4G).

**Figure 4.**
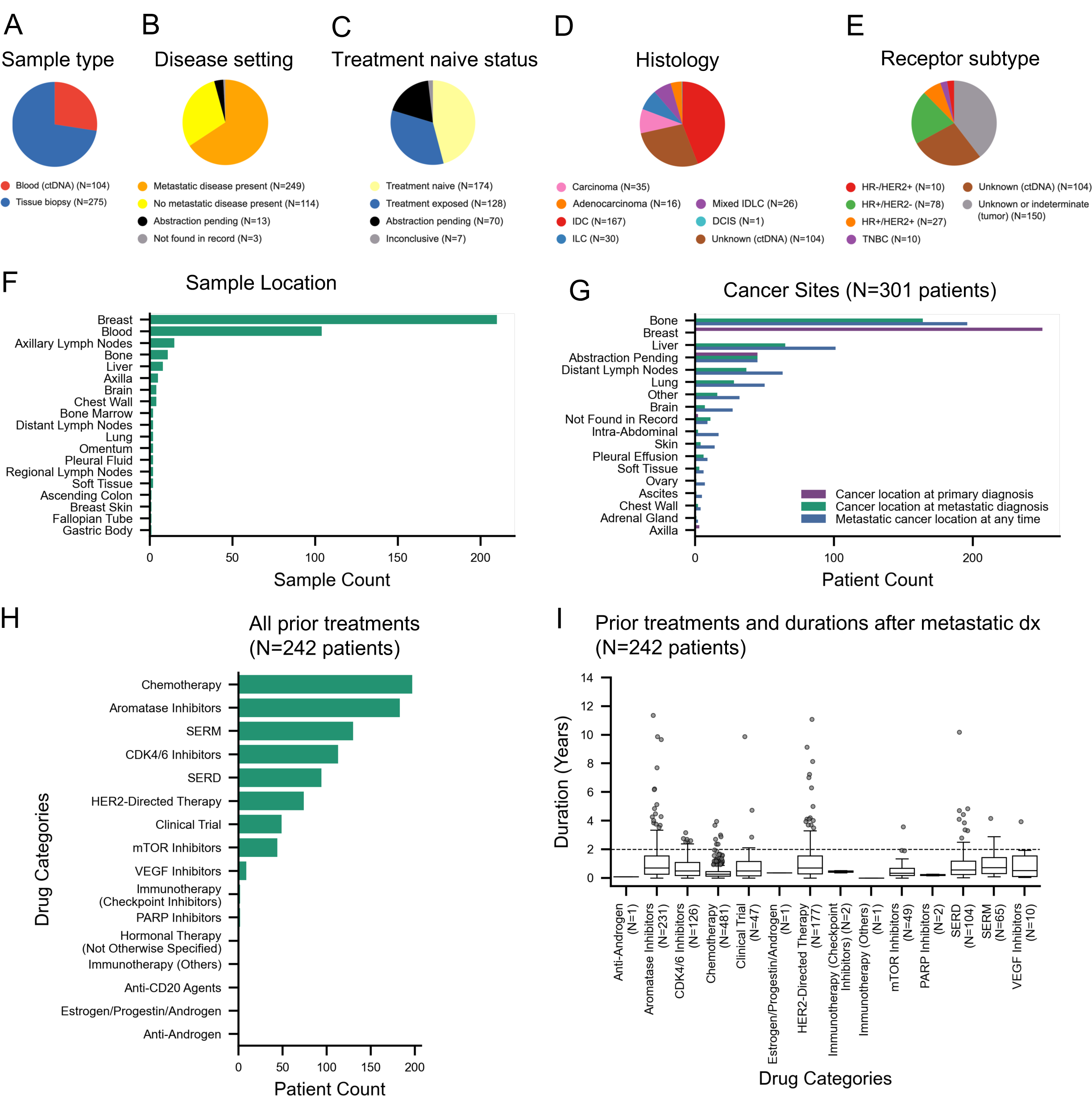
Clinical attributes from medical records in the Metastatic Breast Cancer Project clinico-genomic dataset (N=379 tumors, 301 patients). (A) Distribution of sample type based on source of tumor: FFPE tissue biopsy or ctDNA from blood. (B) Distribution of disease setting based on whether a sample was taken before metastatic diagnosis (‘no metastatic disease present’) or after metastatic diagnosis (‘metastatic disease present’). (C) Distribution of treatment naive status of samples, based on whether a sample was collected before (‘treatment naive’) or after (‘treatment exposed’) the patient received systemic treatment. (D) Distribution of histology of samples. Samples were classified as IDC, ILC, mixed IDLC, DCIS, carcinoma and adenocarcinoma. Histology information is unknown for ctDNA samples. (E) Distribution of receptor subtype of samples. Receptor subtype (HR+/HER2-, HR+/HER2+, HR-/HER2+ and TNBC) was assigned based on the ER, PR, and HER2 receptor status of the samples. Receptor subtype information is unknown for ctDNA samples. (F) Distribution of anatomical locations from which samples were obtained. (G) Distribution of cancer sites at the time of primary diagnosis, metastatic diagnosis, and any time throughout the course of the disease for each patient. (H) Distribution of prior treatments received by drug category for each patient. Treatment information was abstracted for N=242 patients out of 301 patients, with abstraction pending for the rest of the patients. (I) Prior treatments received per patient and treatment duration. The dotted line denotes a treatment duration of 2 years, which we used to define treatment for which there was an extraordinary response. WES, Whole Exome Sequencing; ctDNA, circulating tumor DNA; HR, Hormone Receptor; HER2, Human Epidermal Growth Factor Receptor 2.

Detailed treatment information, including treatments, start and stop dates of treatment regimens, treatment mode (e.g. adjuvant), and response to treatment were abstracted for 242 patients (Suppl. Fig. S3B, Fig. 4H-I). Similarly to what was observed in the PRD, 32.6% of patients (N = 79) exhibited extraordinary responses (treatment duration ≥ 2 years), which spanned multiple drug categories (Fig. 4I).

### Medical record and patient reported data in the MBCproject are highly concordant

To evaluate the consistency of the clinical data collected from PRD and MRD, we compared several clinical parameters between them. Direct comparison of values, for patients’ date of birth (N = 248, 97.5% concordance), date of primary disease diagnosis (N = 242, 93% concordance), date of metastatic disease diagnosis (N = 234, 94.9% concordance) and time to metastatic disease (N = 238, 90.8% concordance) yielded a high concordance rate (Suppl. Fig. S6A, Suppl. Table S5A). Similarly, a high concordance was observed when comparing PRD and MRD information of a patients’ receptor status (hormone receptor status: N = 290, concordance = 92.1%; HER2 receptor status: N = 272, concordance = 87.9%; triple negative status: N = 93, concordance = 91.4%) and histology (IDC/ILC: N = 80, concordance = 91.2%; mixed histology: N = 80, concordance = 87.5%) during their disease course (Suppl. Fig. S6A, Suppl. Table S5B). These analyses yielded a median concordance rate of 91.4% (range 87.5%-97.5%) (Suppl. Fig. S6A).

We additionally compared the PRD and MRD of clinical data related to metastatic sites and treatment received, using MRD as a reference to calculate the precision and sensitivity of PRD. When comparing the presence of specific metastatic sites of interest at the time of metastatic diagnosis, we found a high concordance between MRD and PRD (bone only: sensitivity = 72.7%, precision = 90.6%; visceral metastatic sites: sensitivity = 90%, precision = 80.4%; brain: sensitivity = 100%, precision = 66.7%, Suppl. Table S5C). When comparing the top 3 most frequent treatments according to MRD given following metastatic diagnosis (letrozole, fulvestrant, and palbociclib), we also found a high concordance (letrozole: sensitivity = 76.7%, precision = 80.7%; palbociclib: sensitivity = 94.3%, precision = 76.9%; fulvestrant: sensitivity = 60.9%, precision = 73.7%) (Suppl. Fig. S6F). A qualitatively similar result was obtained when comparing the top 10 most frequent treatments given following metastatic diagnosis, which showed a median sensitivity of 71.2% (range: 37.9 - 94.3%) and a median precision of 86.4% (range: 59.5% - 95.2%).

Overall, we found that there was a high degree of concordance between various types of clinical data obtained from MRD and PRD (diagnostic dates, tumor histology, tumor sites, treatments received). The concordance between PRD and MRD from MBCproject highlights the potential each of them has to study the clinical characteristics of MBC patients.

### The somatic genomic landscape of primary and metastatic tumors from the MBCproject revealed gene alterations and mutational processes associated with metastatic disease and treatment resistance

The MBCproject clinico-genomic dataset consisted of 379 tumor samples from 301 patients. These tumors had a median purity of 0.35 (Suppl. Fig. S7A-B) and a median ploidy of 2.32 (Suppl. Fig. S7A-D). Among tumor sites, purity was higher in tumors obtained from tissue (median 0.4) than in those obtained from ctDNA (median 0.23) (Suppl. Fig. S7A). Among the most common tumor sites obtained from tissue (breast, axillary lymph nodes, bone, and liver), most had a similar median purity (medians between 0.39 and 0.41), the exceptions being bone sites, which had a lower purity (median 0.26) (Suppl. Fig. S7B).

Analysis of 157 treatment naive tumors from the breast and breast-adjacent sites (axilla or axillary lymph nodes) revealed significant enrichment of mutations in 11 genes, including known cancer-related genes (using MutSig2CV^20^) (Fig. 5A). *TP53*, *PIK3CA*, *CDH1*, *GATA3*, *MAP3K1*, *PALB2*, *PTEN, ARID1A*, *FANCD2* were amongst the top frequently mutated genes in this cohort. 17 cancer genes, including *TP53*, *PIK3CA*, *ERBB2*, *CDH1*, *ESR1*, *AKT1, PTEN,* and *NF1,* were significantly altered in 249 metastatic tumors (Fig. 5B). Mutations in *ESR1* were enriched in treatment exposed tumors obtained after metastatic diagnosis as compared to treatment naive tumors obtained before metastatic diagnosis (22.2% vs 3.4%, p=1.1 x 10^-4^; two-sided Fisher exact test) (Suppl. Fig. S8A). Comparison of treatment naive breast and breast-adjacent tumors obtained before metastatic diagnosis with treatment exposed tumors from distant metastatic sites revealed an enrichment in the latter of mutations in both *ESR1* (26.6% vs 3.4%, p=2.2 x 10^-5^; two-sided Fisher exact test) and *AKT1* (6.6% vs 0%, p=2.0 x 10^-2^; two-sided Fisher exact test) (Suppl. Fig. S8B). Alterations in metastatic tumors were classified into therapeutic levels based on OncoKB^21^. 39% of tumors collected in metastatic setting (97 samples from 92 patients) harbored clinically actionable mutations across 11 genes, which included mutations in canonical oncogenic pathways like PI3K signaling (*PIK3CA, AKT1, TSC2*), receptor tyrosine kinase (RTK)/RAS/MAPK signaling (*ERBB2*) and DNA damage repair (*ATM*) (Fig. 5C)^21, 22^.

**Figure 5.**
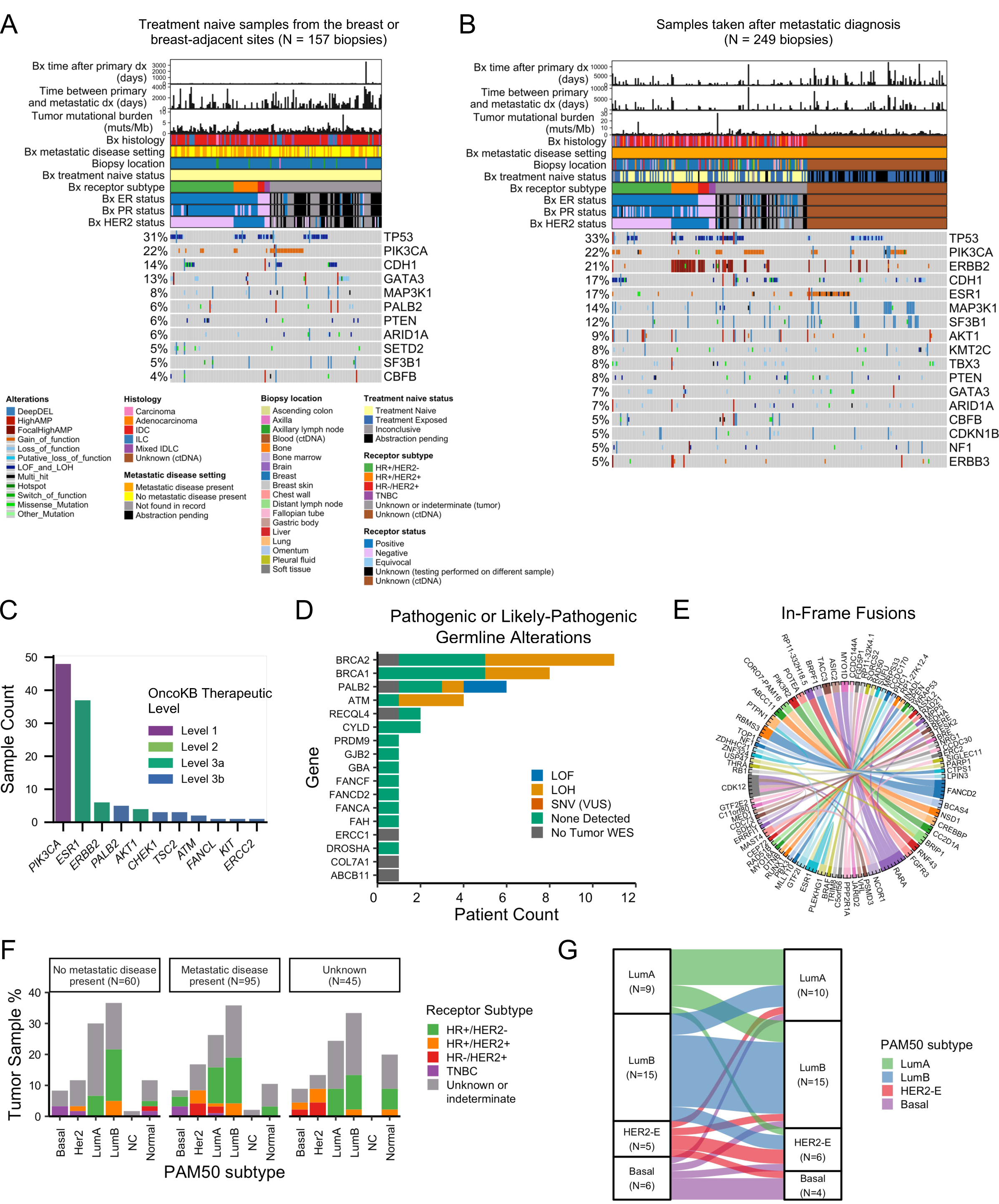
Genomic and transcriptomic features of metastatic breast cancer. (A) Genomic landscape of treatment naive samples from breast and breast-adjacent sites (N=157 tumors). (B) Genomic landscape of tumors taken after metastatic diagnosis (N=249 tumors, 209 patients). For panels A and B, Somatic alterations (SNVs, indels, and copy number variations) across cancer related genes that were significantly altered (based on Mutsig2CV) in each of these cohorts are shown, along with the tumor mutation burden per samples calculated from the non-synonymous somatic alterations. Clinical features abstracted from medical records associated with each sample are shown. Clinical features shown are tumor histology, receptor subtype, biopsy location, metastatic disease setting, treatment naive status, time between primary diagnosis and sample collection, time between primary and metastatic diagnosis. (C) Distribution of clinically actionable mutations per gene in samples taken after metastatic diagnosis (N=249 samples, 209 patients). Clinically actionable mutations are based on OncoKB. Levels 1 and 2 are standard-of-care categories that include mutations recognized by the FDA or other professional organizations as predictive of response to FDA-approved drugs. Levels 3a and 3b are investigational categories that include mutations predictive of response to a drug in breast cancer or an FDA-approved drug in another indication, respectively. (D) Distribution of pathogenic/likely-pathogenic germline variants (based on CharGer) in cancer predisposition genes (N=377 patients). The number of germline variants with a concurrent somatic event (loss of heterozygosity, loss of function mutations, and variants of unknown significance) in the tumor samples are shown. (E) Landscape of oncogenic or likely-oncogenic in-frame fusions (based on OncoKB) (N=200 tumors, 141 patients). Each line of unit width denotes a unique gene fusion and the ends of the line point to the gene pairs in the fusion. (F) Distribution of transcriptional subtypes (research-based PAM50) based on metastatic disease setting (N=200 tumors, 141 patients). The number of samples for each receptor subtype is shown. (G) Transcriptional subtype switching in paired tumors from the same patient (N=35 patients). The width of the line denotes the number of patients in which the specified subtype switch was observed. Tumors with a Normal subtype or not classified were excluded. SNV, Single nucleotide variant; WES, Whole exome sequencing; ctDNA, circulating tumor DNA; FDA, Food and drug administration; ER, Estrogen receptor; PR, Progesterone receptor;HER2, Human epidermal growth factor receptor 2; LumA, Luminal A; LumB, Luminal B; HER2-E, HER2-enriched; NC, Not classified.

To explore whether the genes we found to be with associated metastatic disease and treatment exposure are acquired or enriched during metastatic disease progression and treatment, we performed tumor evolutionary analysis (Table 3) (using ABSOLUTE^23^ and PhylogicNDT^24^). The analysis was performed on the 14 patients with 3 or more tumor biopsies, which included 12 patients with biopsies from more than 1 timepoint and 2 patients with multiple biopsies from the same timepoint (Table 3, Suppl. Table S6). Shared clonal non-synonymous mutations in cancer-related genes (i.e., putative truncal mutations) among tumors were found for 13/14 of these patients. These putative truncal mutations included oncogenic mutations in breast cancer genes such as *PIK3CA*, *GATA3*, *TP53*, *CDH1*, and *PTEN*. Acquired non-synonymous mutations in cancer-related genes with respect to the earliest sequenced sample were found for 11/12 of the patients with biopsies from more than 1 timepoint. These acquired mutations included oncogenic mutations in genes previously found to be enriched in metastatic breast cancer and/or associated with resistance to endocrine therapy^5, 7, 25, 26^, including *ESR1* (acquired in 3 patients), *NF1* (acquired in 1 patient), and *TP53* (acquired in 1 patient). Private clonal mutations in cancer-related genes between tumors were found in the 2 patients with multiple biopsies from the same timepoint. For one of these patients (patient 734), there were oncogenic *PIK3CA* and *ARID1A* clonal mutations private to only a subset of the 3 breast biopsies, indicative of the clonal heterogeneity across these breast tumor sites.

**Table 3.** Tumor heterogeneity and tumor evolution in patients with three or more tumor samples (N=14 patients). Mutations in selected genes shared and private/acquired across biopsies sequenced at different timepoints or locations from a patient, highlighting tumor heterogeneity and evolution in these patients.

| Patient ID | Sample IDs | Reference sample | No. of samples | No. of timepoints | Relative bx time (days) | Bx location | Shared clonal mutations in key genes | Private clonal mutations (tumor heterogeneity) or acquired mutations (tumor evolution) in key genes |
| --- | --- | --- | --- | --- | --- | --- | --- | --- |
| **Tumor Heterogeneity** |  |  |  |  |  |  |  |  |
| 734 | T1A, T1B, T1K |  | 3 | 1 | 46, 46, 46 | BREAST, BREAST, BREAST | CDKN2A R80*, RECQL5 L895M | ARID1A S1338fs, PIK3CA N1044Y, POLG R1071C |
| 6109 | T1A, T1B, T1C |  | 3 | 1 | 10, 10, 10 | BREAST, BREAST, BREAST | APC A474T, GATA3 G334fs, PTPRD P155A, SETDB1 A560G, UBR5 R284C | ARID1A Q1145H, PBRM1 K1074E, POLK F659Y, RAD23A A91P, UBR5 R285H |
| **Tumor Evolution** |  |  |  |  |  |  |  |  |
| 2991 | T1A, T1B, T2 | T1A | 3 | 2 | 93, 93, 360 | BREAST, BREAST, BREAST | PIK3CA N345K | CDH1 Q448*, CDH1 T199fs, KMT2C R4225*, MAP3K1 I1461M, MAP3K1 K1463R, MAPK1 E81V, NCOR1 Q113*, NF1 D633fs, PIK3CA E545K |
| 3896 | T2, T4, T5 | T2 | 3 | 3 | 0, 350, 410 | BREAST, AXILLARY LYMPH NODE, PLEURAL FLUID | ESR1 Y537N, PIK3CA H1047R, TP53BP1 Q26* | APEX1 L244fs, ESR1 E380Q, MDM4 A348P, RARA D221G |
| 857 | T1, T2, T3 | T1 | 3 | 3 | 0, 34, 186 | BREAST, LIVER, BREAST | PABPC1 R436C | COL1A1 E1297*, JAK2 R138G |
| 95 | T1, T2, T3 | T1 | 3 | 3 | 0, 39, 263 | BREAST, LIVER, BREAST | RECQL5 E308D, TP53 P98fs, USP7 G916V | ERBB4 G917* |
| 128 | T1, T2, T3 | T1 | 3 | 3 | 0, 37, 2071 | BREAST, BREAST, BONE | KRAS G12A, PIAS3 R63* | ESR1 D538G, EZR P118L, RARA R272W |
| 1160 | T1, T2, T3 | T1 | 3 | 3 | 0, 858, 1011 | BREAST, SOFT TISSUE, SOFT TISSUE | TP53 R175H, TP63 R343fs | AKT3 H236Y |
| 3717 | T1, T2, T3, BLOOD | T1 | 4 | 4 | 2015, 2282, 2689, 3503 | LYMPH NODE DISTANT, AXILLARY LYMPH NODE, AXILLARY LYMPH NODE, BLOOD | KDR L840F, SETD2 S1001F, TP53 R248Q | ESR1 E380Q, FANCA R914K |
| 3946 | T1, T2, T3, T4 | T1 | 4 | 4 | -21, 44, 798, 805 | AXILLA, BONE, ASCENDING COLON, FALLOPIAN TUBE | CDH1 Q23*, POLR2B D1017N | SETDB1 A1014S |
| 63 | T1, T2, T3A, T3B | T1 | 4 | 3 | 0, 16, 161, 161 | BREAST, BONE, BREAST, BREAST | CDH1 Q23*, PTEN VL317fs |  |
| 5856 | T0A, T0B, T0C, T2 | T0B | 4 | 2 | 0, 0, 0, 198 | BREAST, BREAST, AXILLARY LYMPH NODE, BREAST | CDH1 F267fs | DNMT3A H506Y |
| 203 | T1B, T1C, T1D, T2, T3 | T1B | 5 | 3 | 0, 0, 0, 166, 1211 | BREAST, BREAST, BREAST, BREAST, BONE | DDX5 M134I | ATM L2492R, PRDM2 ED282del, PRDM2 P1006L |
| 4384 | T1, T2, T3, T4, BLOOD | T1 | 5 | 5 | 3419, 4658, 5288, 5754, 7989 | BREAST, AXILLA, AXILLA, CHEST WALL, BLOOD |  | ASXL1 G949D, KMT2D Q1479L, MTOR D2424N, TP53 C135R |

*De novo* mutational signature analysis identified an APOBEC dominant signature, a homologous recombination (HR)-dominant signature, and a clock-like dominant mutational signature in the cohort (using SignatureAnalyzer^27^) (Suppl. Fig. S9A). Tumors with the APOBEC and HR signatures as their dominant signature were enriched in samples taken after metastatic diagnosis when compared to those taken before metastatic diagnosis (APOBEC: 23.7% vs 11.4%, p = 0.0068; HR: 41.4% vs 21.9%, p = 0.00036; two-sided Fisher exact test) (Suppl. Table S7A, Suppl. Fig. S9B)^5, 8^. The opposite effect was seen in tumors with the Clock-like signature as their dominant signature (Clock-like: 31.7% vs 53.5%, p=0.00012; two-sided Fisher exact test). A similar enrichment of APOBEC and HR signatures and a depletion of Clock-like signatures was seen when comparing treatment naive tumors taken before metastatic diagnosis and treatment exposed tumor taken after metastatic diagnosis (Suppl. Table S7B), and when comparing the strength of COSMIC signatures 1 (Clock-like), 3 (HR), 2 (APOBEC), and 13 (APOBEC) between groups (using deconstructSigs^28^) (Suppl. Fig. S9B-C).

No statistically significant differences in tumor mutational burden (TMB) were observed between tumors based on their metastatic diagnosis (Suppl. Fig. S9D) or treatment exposure (Suppl. Fig. 9E), although the few hypermutated tumors identified (TMB >=10 mutations/Mb) were all in the metastatic setting (3/249 tumors in the metastatic setting) (Suppl. Fig. S9D). Comparing the TMB of tumors based on their dominant signature, we found that tumors with APOBEC as their dominant signature have a higher TMB (p<0.0001 for all cases; Wilcoxon two-sided test) (Suppl. Table S9F), consistent with what has been reported previously^29^.

### The germline genomic landscape of MBCproject revealed variants associated with cancer risk and cancer diagnosis at a young age

We analyzed germline samples from 377 patients in order to identify pathogenic/likely-pathogenic variants in genes associated with cancer risk^30^. 11.4% (N=43/377) of patients had at least one pathogenic/likely-pathogenic variant in cancer predisposition genes (using CharGer^30^), numerically higher than what was found in the TCGA breast cancer cohort (9.6%, N=103/1076^30^) (p = 0.32, two-sided Fisher exact test). The most prevalent genes were among 12 established breast cancer predisposition genes^31^ and are *BRCA1* (2.1%, N=8)*, BRCA2* (2.9%, N=11), *BRCA1* or *BRCA2* (4.8%, N=18), *ATM* (1.1%, N=4) and *PALB2* (1.6%, N=6) (Fig. 5D). The frequency of patients with variants in any of these 12 breast cancer predisposition genes in MBCproject (7.4%, N=28/377) was numerically higher than in the TCGA breast cancer cohort (5.6%, N=60/1076) (p = 0.21, two-sided Fisher exact test), consistent with the modest increase in prevalence seen in MBC compared to non-metastatic BC^32^. 5.0% (N=15 of 301 patients with a tumor exome) of patients were found to have pathogenic/likely-pathogenic variants in cancer predisposition genes along with a concurrent somatic event (loss of heterozygosity, loss of function mutations, or variants of unknown significance) in the same gene (Fig. 5D).

To evaluate whether there is an association between the age of cancer onset and the presence of pathogenic/likely-pathogenic germline variants, we compared the age at primary breast cancer diagnosis of patients with or without pathogenic/likely-pathogenic germline variants. We found a statistically significant difference in age distribution between the groups when considering pathogenic/likely-pathogenic variants in *BRCA1* or *BRCA2* (D=0.39, p=0.011; two-sided, two-sample Kolmogorov–Smirnov test) (Suppl. Table S8A), with patients with these variants having a lower median age than others (median age of 37 and 45 years). A similar result was obtained when considering pathogenic/likely-pathogenic variants in cancer predisposition genes, with the median age of patients with these variants being lower than those without (median age of 41 and 45 years) and the age distribution between these groups being statistically significant (D=0.26, p = 0.0098; two-sided, two-sample Kolmogorov–Smirnov test) (Suppl. Table S8A). We found similar results when comparing the frequency of pathogenic/likely-pathogenic variants in patients diagnosed before/at or after the age of 40 years old, with patients with variants in the gene groups considered being enriched in the younger compared to the older group, and with the enrichment being statistically significant in the *BRCA1* or *BRCA2* (9.2% vs 2.5%, p=0.0089; two-sided Fisher exact test) but not for the cancer predisposition gene case (16.2% vs 9.1%, p=0.06; two-sided Fisher exact test). These results are consistent with recent work finding that the presence of pathogenic/likely-pathogenic germline variants in cancer associated genes, particularly *BRCA1* or *BRCA2*, is associated with the diagnosis of breast cancer at a young age^33^.

### The transcriptomic landscape of MBCproject revealed the occurrence of transcriptional subtype switching across a patient’s tumors and the presence of in-frame fusions in cancer-related genes

The MBCproject transcriptomic dataset consisted of 200 tumor samples from 141 patients. 59 and 91 of these tumors were taken before and after metastatic diagnosis, respectively (Suppl. Fig. S10B). The number of biopsies and timepoints per patient ranged between 1 and 5, with 44 patients having more than 1 tumor biopsy (Suppl. Fig. S3B, Suppl. Table S9A) and 22 patients having tumor biopsies from more than 1 timepoint (Suppl. Fig. S3B, Suppl. Table S9B). These tumors had a median number of genes detected of 19,888 (Suppl. Fig. S10A), which was similar among the most common tumor sites (breast, axillary lymph nodes, bone, liver) (medians between 19,336 and 20,541) (Suppl. Fig. S10C).

We analyzed the transcriptome of these tumors to identify gene fusions that could be driving oncogenic behaviors in these tumors. 80.5% (161) of tumors were found to have at least one fusion (Suppl. Fig. S10D), with a median of 5 fusions per tumor. 12% (24) of tumors had an in-frame fusion classified as oncogenic or likely-oncogenic by OncoKB, including fusions in breast cancer genes such as *FANCD2* (N=3 tumors, 1 patient), *FGFR3* (N=2 tumors, 1 patient), *ESR1* (N=1 tumor), *BRAF* (N=1 tumor), and *NCOR1* (N=1 tumor) (Fig. 5E). 13% (26) of tumors had in-frame fusions involving a kinase gene (e.g. *FGFR3, BRAF, ERBB2, IGF1R*), of which 5 fusions across 5 tumors were classified as oncogenic or likely-oncogenic (e.g. *BRAF*-*MRPS33*, *RARA*-*CDK12*, and *FGFR3*-*TACC3*) (Suppl. Fig. S11A, Suppl. Table S10A). 16% (32) of tumors had in-frame fusions involving cancer genes not found in fusion databases, which involved genes such as *RB1*, *NF1*, and *FOXO3* (Suppl. Fig. S11B, Suppl. Table S10B).

To characterize the transcriptional subtype of these tumors, we applied a research-based PAM50 classifier to obtain their intrinsic molecular subtype. The distribution of molecular subtypes was 27.0% (54) Luminal A, 35.5% (71) Luminal B, 14.5% (29) HER2-Enriched, 8.5% (17) Basal, 13% (13) Normal, and 1.5% (3) not classified (NC) (Suppl. Fig. S12A). The molecular subtype of these tumors was consistent with their receptor subtype: HR+/HER2-tumors were primarily Luminal A or Luminal B, HER2 positive tumors were primarily HER2-Enriched, and TNBC tumors were primarily Basal (Fig. 5F). Consistent with these results, *ESR1* expression was the highest in Luminal A or Luminal B tumors compared to other subtypes (Supp. Fig. S12B), and was higher in HR-positive compared to HR-negative tumors (Supp. Fig. S12C).

Comparing the frequency of molecular subtypes in MBCproject with that of The Cancer Genome Atlas Breast Invasive Carcinoma (TCGA BRCA), which is composed primarily of primary tumors not in the metastatic setting, we found a statistically significant depletion of the Luminal A subtype in MBCproject with respect to TCGA BRCA (27.0% vs 42.1%, p=5.6e-05; two-sided Fisher exact test) (Suppl. Table S11A). A depletion of the Luminal A subtype was also observed when comparing MBCproject tumors after (26.3%) or before (30.0%) metastatic diagnosis with TCGA BRCA primary tumors (42.5%) (p=0.0022 and p=0.060, respectively; two-sided Fisher exact test) (Suppl. Table S11A). This depletion of Luminal A tumors in MBCproject is consistent with the high prevalence of Luminal A subtype switching between paired primary and metastatic tumors found in previous work^8, 34, 35^.

To study the switching and heterogeneity of molecular subtypes across a patient’s tumors, we analyzed the subtype of paired tumors from patients with 2 or more samples (35 patients, excluding samples with Normal or NC subtype) (Fig. 5G). Note that, unlike prior work on subtype switching, paired tumors in this study can correspond not only to paired primary/metastatic samples, but also paired metastatic samples from distinct sites or paired concurrent samples from the primary tumor. We observed a switch in molecular subtype in 15 (43%) patients (Suppl. Table S11B). Luminal A and Luminal B were the most common subtypes involved in switching (4 patients switched from Luminal A, 5 patients switched from Luminal B). Switching from the Basal and HER2-E subtypes, which is less common, was also observed (3 patients switched from HER2-E, 3 patients switched from HER2-E) and involved switching to all other subtypes. These findings are consistent with previous work on subtype switching^8, 34, 35^, including the observed high rate of switching (43% in our case, >30% in other studies).

### A patient case study leveraging the breadth and depth of clinico-genomic data identified putative drivers of endocrine therapy resistance during disease progression

To illustrate the type of insights on disease progression and treatment resistance that the clinco-genomic dataset enables, we present the case study of patient 3717 (Fig. 6). This patient had an initial diagnosis of ductal carcinoma *in situ* (DCIS) and developed metastatic disease 5.5 years later (Fig. 6A). This patient received multiple lines of treatment during the course of their metastatic disease (9 distinct therapies, including endocrine therapy, CDK4/6 inhibitors, HER2-directed therapies, and chemotherapy). The patient died 5.3 years after their metastatic diagnosis, 10.7 years after their primary diagnosis. As part of the MBCproject dataset, we have 4 samples collected over this patient’s disease course: WES and RNA-seq for 3 metastatic tumors and WES for a ctDNA sample. One of these samples was collected at metastatic diagnosis (3717_T1, 5.5 years after primary diagnosis) and the rest were collected following metastatic diagnosis (3717_T2, 3717_T3, and 3717_BLOOD_P, collected 6.3, 7.4, and 9.6 years after primary diagnosis, respectively) (Fig. 6B).

**Figure 6.**
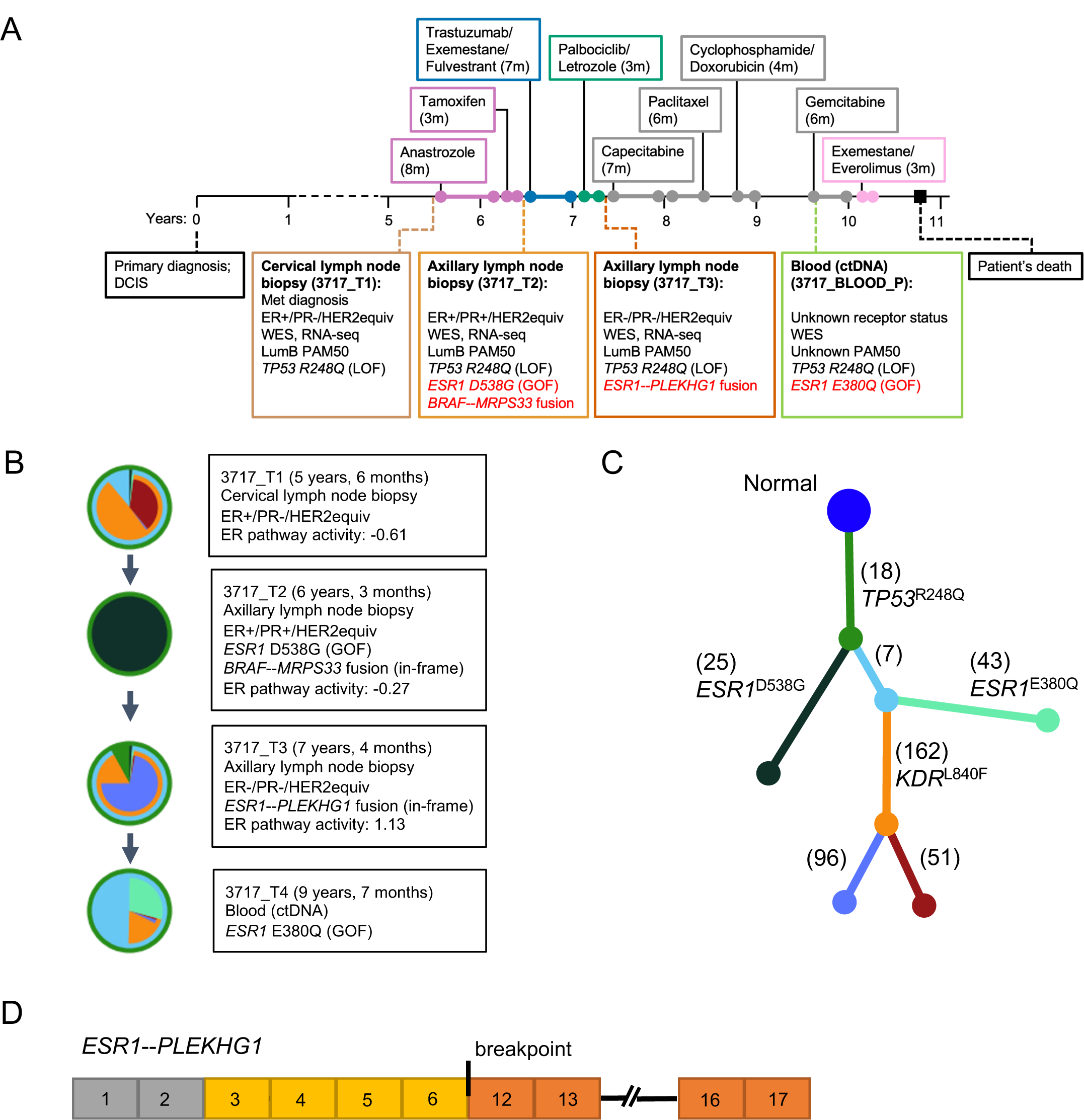
Patient case study leveraging the range of clinico-genomic data types from Metastatic Breast Cancer Project. Case study of patient 3717, a patient in the MBCproject clinico-genomic dataset, which includes a detailed treatment timeline and associated genomic and transcriptomic features. (A) Timeline depicting treatments received, diagnostic information, and sequenced biopsies for this patient. Diagnostic information for each biopsy is included in the top of each panel. Selected oncogenic mutations, fusions, and intrinsic molecular subtype identified from WES and RNA-seq are included in the bottom of each panel. (B, C) Tumor evolutionary analysis of this patient’s tumors. Pie charts (panel B) denote the relative abundance of each clone in the tumor (each clone is assigned a different color), with the phylogenetic tree (panel C) defining the mutations that characterize each clone. The boxes adjacent to each pie chart describe the clinico-genomic features associated with the tumor. (D) Fusion structure of the *ESR1*--*PLEKHG1* in-frame fusion identified in tumor 3717_T3. The diagram depicts the fusion structure formed between *ESR1* (shown in yellow and gray) and *PLEKHG1* (shown in orange), with a breakpoint located after exon 6 of *ESR1*. MBCproject, The Metastatic Breast Cancer Project; WES, Whole exome sequencing; RNA-Seq, RNA sequencing.

During the course of this patient’s disease, there was a loss of ER expression based on IHC, with the first and second biopsies (3717_T1, 3717_T2) being ER-positive and the third biopsy (3717_T3) being ER-negative (Fig. 6A, Suppl. Table S12). Subsequent biopsies from this patient, which were not sequenced, were found to be ER-negative (7.9 years after primary diagnosis) and ER-positive (9.5 years after primary diagnosis). From the transcriptome of the sequenced tumors, we found that all three tumors had a Luminal B subtype (Fig. 6A) and *ESR1* expression consistent with that of ER-positive samples (*ESR1* log2(TPM+1) > 5.5 in patient 3717’s tumors vs an interquartile range of 5.17-6.72 and 1.63-3.33 in ER-positive and ER-negative tumors, respectively) (Suppl. Table S12). From the activity of ER pathway transcriptional signatures (Hallmark gene sets^36^), there is also a mismatch between their ER status and the expected pathway activity (ER pathway activity in the ER-negative interquartile range of −1.33 to −0.03 for 3717_T1 and 3717_T2, and in the ER-positive interquartile range of 0.16 to 2.12 for 3717_T3) (Suppl. Table S12). The inconsistency between ER IHC, ER pathway transcriptional data, and molecular subtype could reflect the heterogeneity in the tumor sample that resulted in both ER-positive and ER-negative metastatic tumors.

Tumor evolutionary analysis of the 4 sequenced tumors showed the emergence of clones with distinct driver oncogenic mutations throughout the disease, all of which share truncal mutations that include an oncogenic TP53 mutation (Fig. 6B-C). Among the driver oncogenic mutations are an activating near-clonal *KDR*^L840F^ mutation in the first tumor (3717_T1) that is present in all but the second tumor (3717_T2), a clonal activating *ESR1*^D538G^ mutation in the second tumor that was almost undetectable in the first tumor, and a subclonal activating *ESR1*^D5380Q^ mutation found only in the fourth sample (3717_BLOOD_P). The acquisition of these activating *ESR1* mutations is consistent with the tumors becoming resistant to endocrine therapy following the multiple lines received. Exome analysis alone did not reveal a clear genomic driver of endocrine therapy resistance in the third tumor (3717_T3), although it was found to be ER-negative by IHC.

To look for additional drivers of endocrine therapy resistance, we leveraged the transcriptome of these tumors. For the endocrine resistant tumor without *ESR1* mutations (3717_T3), we found an in-frame *ESR1-PLEKHG1* fusion as a putative resistant driver^37^. For the *ESR1*^D5380Q^-mutant tumor (3717_T2) we found an in-frame *BRAF*-*MRPS33* fusion, which could be contributing to the resistance phenotype. The presence or absence of *ESR1* alterations were found to be consistent with the activity of ER pathway transcriptional signatures, with the activity in the *ESR1* wildtype tumor (3717_T1, ER pathway activity: −0.61) being lower than that of the *ESR1*^D5380Q^-mutant (3717_T2, ER pathway activity: −0.27) and *ESR1-PLEKHG1*-fusion (3717_T3, ER pathway activity: 1.13) tumors (Suppl. Table S12). Overall, this joint genomic and transcriptomic analysis was able to identify putative drivers of endocrine resistance in each of the patient’s tumors.

## Discussion

In this work, we introduced the Metastatic Breast Cancer Project (MBCproject), a patient-partnered research project that aims to accelerate discoveries that impact the clinical outcomes of patients with metastatic breast cancer and beyond. The MBCproject was built with deep patient involvement and allows patients to share their samples, clinical experiences, and their voices. From October 2015 to March 2020, 3,246 patients diagnosed with metastatic breast cancer and treated at over 1,700 institutions across the United States and Canada consented to participate in the MBCproject and share their samples and clinical data. By collecting tumor and germline samples (saliva, blood, and archived tumor tissue), medical records, and patient-provided information from surveys, the MBCproject generated an integrated and publicly available dataset of genomic, clinical, and patient-reported information that was described throughout this work. The clinico-genomic MBCproject dataset consists of whole exome sequencing of 379 (germline-matched) tumors from 301 patients, RNA sequencing of 200 tumors from 141 patients, and 377 germline samples from 377 patients, with clinical data from medical records and patient-reported information.

Compared to other large-scale genomic studies of metastatic breast cancer (Table 5), key distinguishing characteristics of MBCproject is the availability of both transcriptomic and whole exome sequencing data (Fig. 2, Fig. 5), the ability to study multiple tumor samples from various timepoints in a patient’s disease (Fig. 5G, Fig. 6, Suppl. Table S6, Suppl. Table S9), and the comprehensive clinical data from patient-reported information (Fig. 3) and medical records (Fig. 4). Given these characteristics, the MBCproject dataset is poised for studying several understudied clinical cohorts (e.g. young women with breast cancer, *de novo* metastatic breast cancer, extraordinary responders to treatments), rare disease subtypes (e.g. invasive lobular carcinoma, metaplastic breast cancer), biomarkers of response and resistance (e.g. CDK4/6 inhibitors), real world patterns, among others (Table 4). In addition to the clinically relevant observations presented here, the data that continues to be generated from this ongoing study will further enable a more detailed molecular characterization of metastatic breast cancer. We expect that the MBCproject, in conjunction with the increasing number of MBC genomic studies and datasets, will expand our current understanding of the genomics of metastatic breast cancer, help uncover new mechanisms of resistance to therapies, and ultimately advance precision oncology research (Table 5).

**Table 4.** Specific use cases and examples of scientific questions that can be addressed using MBCproject data. MBCproject, The Metastatic Breast Cancer Project.

| **Types of use cases of MBCproject data** | **Specific examples of scientific questions that can be addressed using MBCproject data** |
| --- | --- |
| Genomic landscape of MBC | What genes show frequent alterations in MBC patients? How do somatic alterations and/or RNA expression of genes compare in primary versus metastatic breast cancer tumor settings? |
| Specific clinical cohorts | Examples; de novo MBC, young women with breast cancer, triple negative breast cancer |
| Rare disease subtypes or patient outliers | For a rare MBC subtype (e.g., lobular or metaplastic) are there any unique genomic features or correlated clinical attributes? What can be learned from disease features of long-term MBC survivors? Are there predictors of extraordinary response to a particular therapy? |
| Tumor evolution and heterogeneity at the patient level | How heterogenous are MBC tumors? Does tumor heterogeneity show concordance with other clinical aspects such as receptor status or response to a given treatment? How do cancers evolve in different disease settings or under specific treatment pressures? |
| Biomarkers of response/resistance | Are there mutational or gene expression correlates among patients who show a response (or develop resistance) to a given treatment? What are the underlying mechanistic pathways of resistance to a treatments like CDK4/6 inhibitors? |
| 'Real world' patterns | What medications are people getting in community healthcare settings? |

**Table 5.**
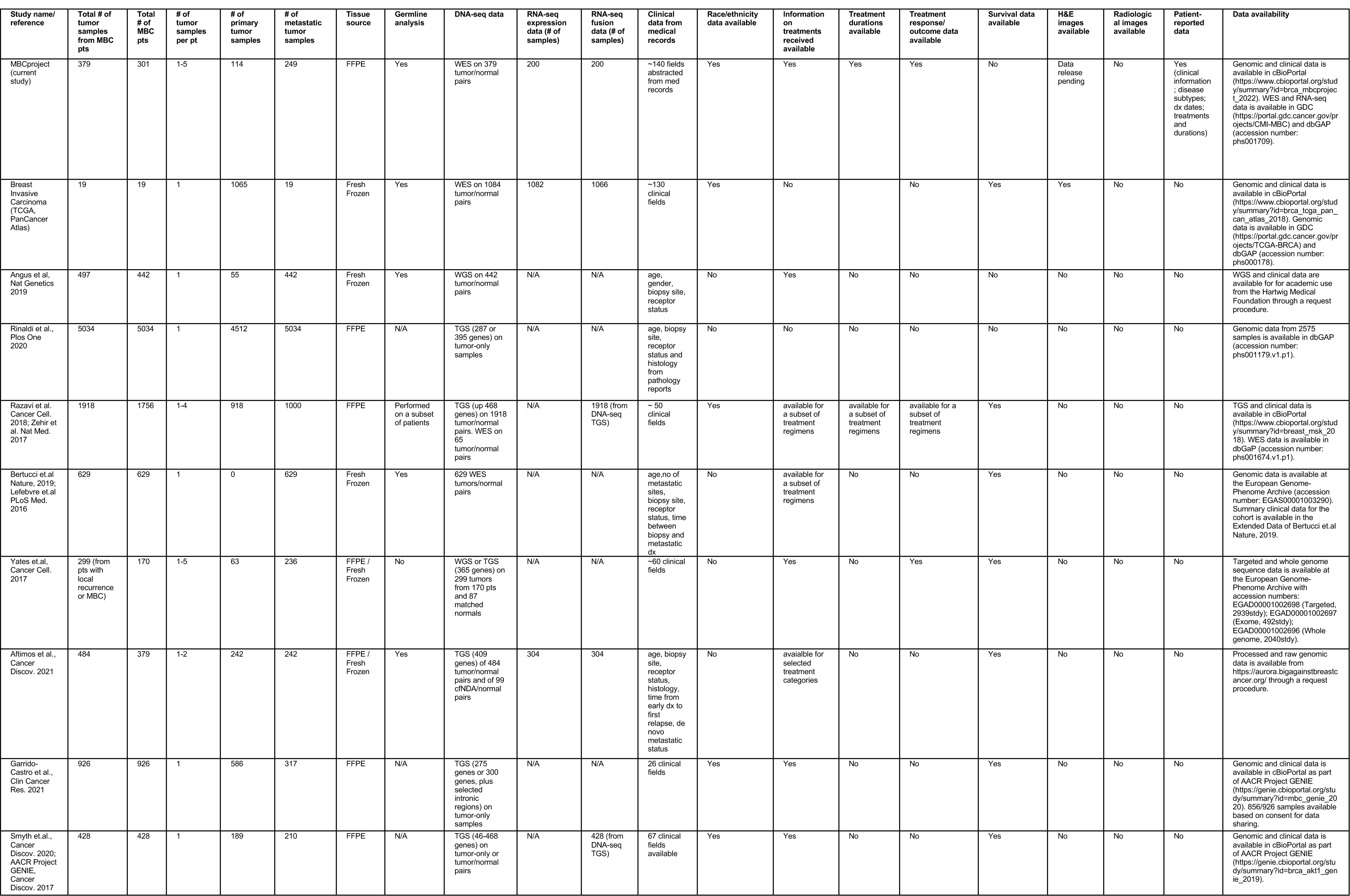
Summary of data types available in clinco-genomic datasets that include patients with metastatic breast cancer.

The MBCproject demonstrates that a patient-partnered study can build a large-scale and high-quality clinco-genomic dataset of real world patients through the remote acquisition of biological samples, patient-reported data, and medical records. To continue to pursue this approach in patient-partnered cancer research beyond breast cancer, the nonprofit organization ‘*Count Me In’* was established in 2018 (www.joincountmein.org). Since August 2021, Count Me In allows any patient in the United States and Canada that have ever been diagnosed with cancer to participate in this research initiative. Count Me In continues to advance efforts to engage with patients across different cancer types, such that every cancer patient regardless of where they live is empowered to contribute to accelerate research discoveries.

A current limitation of MBCproject is that the demographics of its patient population is younger than expected (median age at primary diagnosis of 44 years for the clinico-genomic cohort vs 62 years for the US population diagnosed with breast cancer in 2022^38^ and an estimated 60-69 years for the US population diagnosed with MBC in 2013^39^) and is enriched in patients identifying as non-Hispanic white as compared to the US population (84.4%, N=254/301 in the clinico-genomic cohort vs 57.8% in the 2020 US census^40^). The younger age of the MBCproject cohort was also true when compared to other MBC genomic cohorts (e.g. median age of ≥50 years, N=1756^7^, and 54 years, N=617^5^) and is particularly notable for patients diagnosed before being 40 years old (31.2%, N=79/301 here vs 17.6%, N=309/1756^7^). Potential reasons for the younger ages within the MBCproject are the project’s extensive online engagement approaches associated with patient enrollment (around 50% of patients indicated social media networks like Facebook and Twitter as a referral source) (Suppl. Fig. S2) and a self-selection of patients with less aggressive disease or better responses to treatment (e.g. 30.1%, N=977/3246 of patients reported being on a therapy for over 2 years and 50.0%, N=1622/3246 reported one of their therapies resulting in no evidence of disease) (Suppl. Table S2C), which might be enriched in younger patients. The over-representation of non-Hispanic white patients is consistent with that seen in other cancer cohorts like TCGA^16^ and part of a historical and currently known challenge of unequal representation in clinical research^41^. To facilitate a more diverse patient representation, we have partnered with over 30 non-profit breast cancer advocacy groups (Suppl. Table S1). There exist several community engagement efforts underway to directly reach patients in underrepresented communities, including partnerships with faith-based organizations and colleges/universities, as well as targeted engagement with the Black/African American community. An example of this is the *Amplifying Black Voices Across Cancer* digital initiative (‘#AmpBlackVoices’, www.BlackCancerVoices.org), which was launched in direct collaboration with Black cancer patients, loved ones/caregivers, and advocates. In addition, in partnership with Hispanic and Latino/Latina/Latinx/Latine patients, advocates, and researchers (Suppl. Fig. S1A), the MBCproject was translated/rewritten in Spanish (www.MBCprojectenEspañol.org) and was launched in June 2021^42^.

A distinguishing factor of the MBCproject compared to other clinico-genomic cancer datasets is the availability of clinical data from patient-reported data (N=1604 patients) and medical records (N=301 patients). Even though the traditional approach to obtain clinical data in a research study is through the abstraction of medical records, it is becoming clear (e.g. through All of Us research program^43^) that patient-reported data from surveys can provide information not readily available in medical records, complement the available data, and reach a scale not available otherwise^44, 45^. Yet it is still an open question how to harmonize and compare these two data types and the degree of similarity one should expect given their inherent biases^44, 46^. In the MBCproject, the clinical data of patients obtained from both patient-reported data and medical records gave us a unique opportunity to compare their degree of similarity. We observed a good degree of concordance between them for various types of clinical data (diagnostic dates, receptor status, tumor histology, tumor sites, treatments received). In particular, diagnostic dates and the occurrence of a receptor subtype or tumor histology during their disease course showed a high degree of concordance (concordance range 87.5%-97.5%, Suppl. Fig. S6A-E, Suppl. Table S5A-S5B). We also observed a high but lower degree of concordance when looking at sites at metastatic diagnosis (sensitivity range 72.7%-100%, Suppl. Table S5C) and more variable degree of concordance for metastatic treatments received (median sensitivity 71.2% and range 37.9% - 94.3% for 10 most common treatments, Suppl. Fig. S6F). Despite the lower concordance observed for some of the treatments, it is important to note that these and other discrepancies are not necessarily due to an error in the PRD or MRD. The discrepancies between the MRD and PRD results could be due to either (1) an error in them, (2) not having a complete medical record, (3) the available medical record not being up-to-date with respect to the PRD, or (4) the PRD preceding the obtained medical record. Given the high degree of concordance observed for diagnostic dates and tumor histology/subtype, we hypothesize that the discrepancies are most likely not due to an error, but further work is needed to pinpoint the source of these discrepancies.

Analysis of the genomic (somatic and germline) and transcriptomic dataset was able to recapitulate findings of prior MBC studies, which had separately analyzed each of these data types. For the somatic genomic dataset, we found an enrichment of alterations in the metastatic setting known to be associated with metastatic disease at the gene-level^5, 7, 9, 47^ (*ESR1*, *AKT1*, *PTEN*, *ERBB2*, and *NF1*; which are related to endocrine therapy resistance) (Fig. 5A-B) and at the mutational signature-level^5, 29^ (APOBEC and HR signatures) (Suppl. Figure S9B-C, Suppl. Table S7). We also found known metastatic and/or endocrine therapy associated oncogenic alterations acquired with respect to the earliest available sample (oncogenic mutations in *ESR1*, *NF1*, and *TP53*) (Table 3, Fig. 6) or enriched in treatment exposed distant tumor sites with respect to breast or breast-adjacent treatment naive sites^5, 7, 9^ (alterations in *ESR1* and *AKT1*) (Suppl. Fig. S8B). For the germline genomic dataset, we found that the age of breast cancer diagnosis for patients with pathogenic/likely-pathogenic germline variants in *BRCA1* or *BRCA1* is lower compared to the ones without one (Suppl. Table S8A, Fig. 5D), consistent with what others had found^33, 48^, and found that the prevalence of these mutations is similar to what was recently reported in MBC^32^ (4.8%, 18/377 here vs 5.0%, 129/2595). The prevalence of germline mutations in breast cancer predisposition genes was numerically higher than in TCGA (7.4%, N=28/377 here vs 5.6%, N=60/1076 in TCGA), consistent with recent work^32^. For the transcriptomic dataset, we found that the Luminal A subtype was depleted with respect to the TCGA breast cancer cohort (Suppl. Table S11A, Fig. 5F) and that transcriptional subtype switching between tumors from the same patient was common (42.9%, 15/35 patients with 2 or more tumor samples) (Fig. 5G, Suppl. Table S11B), consistent with recent findings on subtype switching of matched primary and metastatic tumors^8, 34, 35^.

In addition, our multi-omic analysis made potential novel discoveries. We described the landscape of fusions in the MBC transcriptome - to our knowledge, the first large-scale MBC transcriptome-based fusion landscape - and found that putatively oncogenic fusions in cancer related genes are not uncommon: 12% (N=24) of tumors had in-frame fusions classified as oncogenic by OncoKB and 13% (N=26) had in-frame fusions in kinases, which included fusions involving known breast cancer genes *FANCD2*, *FGFR3*, *ESR1*, *BRAF*, and *ERBB2* (Fig. 5E, Suppl. Fig. S11A, Suppl. Table S10B). In particular, the *ESR1* fusion (*ESR1-PLEKHG1*) appeared to be driving endocrine resistance in the tumor it was identified, since it was the only endocrine resistant tumor in this patient without an activating *ESR1* mutation and had no other acquired driver mutations (Fig. 6).

In conclusion, the MBCproject was built using a patient-partnered research approach and resulted in the generation of one of the largest metastatic breast cancer clinico-genomic datasets available. The MBCproject dataset distinguishes itself because of the availability of multiple omic data types (somatic and germline whole exome sequencing, tumor transcriptome sequencing, multiple tumors samples and timepoints per patient) and clinical data sources (from medical record abstraction and patient reported data). The work presented here shows how the MBCproject dataset was able to recapitulate multiple aspects of the current knowledge of metastatic breast cancer genomics from distinct studies. We expect that the richness of the MBCproject dataset and its availability in public data repositories (cBioPortal^49, 50^, dbGap, and the Genomic Data Commons) will make it become an essential resource for studying the genomics of metastatic breast cancer and lead to discoveries to better treat metastatic breast cancer.

## Methods

Please refer to Supplemental Methods for additional details of the methods used in this study.

### Patient and public involvement

A core pillar of the Metastatic Breast Cancer Project is patient engagement. There are several ways the Count Me In team engages with patients and community members. These mechanisms include engagement on social media, by partnering with trusted advocacy organizations, and – for patients who want to be more deeply involved – the development of Project Advisory Councils. In addition, Count Me In and the Metastatic Breast Cancer Project have several efforts aimed at connecting with members of the community who have been traditionally underrepresented in research to listen, learn, and build relationships.

### The Metastatic Breast Cancer Project website

The Metastatic Breast Cancer Project website (MBCproject.org) was developed after extensive collaboration with patients and advocates from the metastatic breast cancer community. The website enables MBC patients across the United States and Canada to learn about the project, register to participate remotely, sign an electronic consent form, and provide information about themselves and their experience via surveys.

### Informed consent

Informed consent (Suppl. File 1) was provided by all patients on a web-based consent form as approved by the Dana-Farber/Harvard Cancer Center Institutional Review Board (DF/HCC Protocol 15-057B). Patient consent allowed the research study team to acquire copies of medical records for abstraction, to send a kit for saliva sample acquisition, to perform sequencing analysis, and to share de-identified linked, clinical, genomic, and patient-reported data publicly. Patients could also opt in to consent to provide a blood sample and/or allow procurement of archived tumor samples for sequencing of germline and tumor DNA. The analyses conducted for this study were performed with information and samples from patients who consented between 09-15-2015 and 03-31-2020.

### Patient-reported data (PRD)

After providing consent, patients have the opportunity to fill out an intake survey questionnaire that asks information about dates of birth, primary and metastatic breast cancer diagnoses, receptor status subtypes, race and ethnicity details (updated with more categories and subcategories and a combined race and ethnicity question to capture more comprehensive demographic information in the latest version of survey), gender (updated to distinguish between gender identity and sex assigned at birth and added multiple options for gender identity in the latest version of survey), treatments associated with extraordinary response and referral source (Suppl. File 1).

In addition to an intake survey, a follow-up survey was deployed in September 2018. Patients who had previously consented for the project received an email notification to fill this survey. Patients who signed up after the survey was deployed, were directed to the survey immediately after completion of the medical release consent form. The follow-up survey asks questions related to the following: sites of metastases (at diagnosis,at any time during course of disease and current), histology and rare subtypes as well as details of current and past treatments received (Suppl. File 1). Patients were given additional survey questions through the blood biopsy consent form (Suppl. File 1). This survey enlisted questions regarding current and past treatments received by the patients along with details of treatment durations and dates (Suppl. File 1). All survey questions were optional except questions regarding country of residence. In all the surveys, responses to questions left blank were categorized as ‘Not reported’. If a participant did not complete a survey, they were categorized as “Participant did not complete survey.”

### Acquisition of medical records

For every patient who enrolled in the study, completed the medical release form (Suppl. File 1) and sent their saliva sample, the study staff requested medical records from institutions and physician offices at which the patient indicated that they received clinical care. Study staff electronically faxed a detailed medical record request form to each facility (Suppl. File 1). A subset of medical records that had not been received after several months were requested again in the same manner. Medical records were received by fax, mail, or secure electronic message. All medical records were saved on a secure drive.

### Acquisition of patient samples

Enrolled patients were mailed separate kits to provide saliva and blood samples. Blood kits were mailed to participants who provided informed consent by signing an additional associated consent form and provided a valid residential address in the US or Canada. Kits containing samples were mailed back in prepaid envelopes to the Broad Institute Genomics Platform. Blood samples were fractionated into plasma and buffy coats. Buffy coats were used to extract germline DNA for WES if no saliva sample was available. If the participant consented to the acquisition of tumor tissue, portions of stored clinical tumor tissue were requested. A form was faxed to each pathology department requesting one H&E-stained slide as well as either 5-μm unstained slides or one formalin-fixed paraffin-embedded tissue block. Requests explicitly stated that no sample should be exhausted in order to fulfill the request. Tissue samples were received at the Broad Institute by mail.

### Medical record abstraction of clinical data

For each medical record (MR) which was acquired by the study team over 100 clinical fields were abstracted following an established clinical data dictionary model (Suppl. File 2) by the study staff abstraction team.. The abstracted fields contain information about clinical and pathological features including pre-diagnostic, diagnostic, staging, surgery and treatments details/dates, information on other cancers and genomic tests details. Each parameter was abstracted to the greatest level of detail as available in the MR, and all dates reported publicly are in days relative to the date of primary diagnosis. This data is referred to as MRD Clinical Comprehensive in Fig. 2. See the Supplemental Methods for details and definitions of key clinical fields abstracted from the MRs. Clinical pathology data was acquired from the pathology reports obtained with each sample. This data is referred to as MRD Pathology in Fig. 2.

### Whole exome sequence data processing and analysis

#### Data processing and quality control

Whole exome sequences for each tissue/blood sample were captured using Illumina technology and the sequencing data processing and analysis was performed using the Picard and Terra pipelines at the Broad Institute. The Picard pipeline (http://picard.sourceforge.net) was used to produce a BAM file with aligned reads. This includes alignment to the GRCh37 human reference sequence using the BWA aligner ^51^ and estimation and recalibration of base quality score with the Genome Analysis Toolkit (GATK) ^52^.

A custom-made cancer genomics analysis pipeline was used to identify somatic alterations using the Terra platform (https://app.terra.bio/). The CGA WES Characterization pipeline developed at the Broad Institute was used to call, filter and annotate somatic mutations and copy number variation. See Supplemental Methods for details of the variant calling pipeline used in this study. GATK CNV^53^ was used for the generation of accurate relative copy-number profiles from the whole exome sequencing data and reference/alternate read counts at heterozygous SNP sites present in both the normal and tumor samples. After accurate proportional coverage profiles are generated for a sample, Allelic CapSeg tool ^54^ was used to generate a segmented allelic copy ratio profile. Allelic copy number profile and mutational call data were modeled jointly by ABSOLUTE^23^ to produce purity for the samples, a discrete copy number profile, and compute cancer cell fractions (CCF). Tumor samples which had a purity of 10% or more were used for downstream analysis.

#### Annotating oncogenic mutations

OncoKB^21^ was used to annotate known oncogenic mutations, identify their effect (e.g. loss of function or gain of function), and if they are known cancer hotspots. For the comutation plots, variants classified as silent or intron are not included. Variants included in the comutation plot that are not in a hotspot region and not assigned an oncogenic effect by OncoKB are classified as either missense, putative loss of function, or other mutation. The multi-hit classification is used if there are multiple alterations in the gene (excluding silent or intron variants).

#### Corrected quantification of copy number, gene deletions, and biallelic inactivations

The inference of gene amplifications, gene deletions, and biallelic inactivations were based on the copy number profile obtained from ABSOLUTE ^23^. To infer biallelic inactivations, mutational events that included both loss of heterozygosity (LOH) and a loss-of-function mutation (LOF) (loss-of-function or likely loss-of-function OncoKB-annotated mutation, or a Nonsense_Mutation, Nonstop_Mutation, Frame_Shift_Del, or Frame_Shift_Del mutation) were used. Gene amplifications and deep deletions were based on the purity corrected measure for the segment containing that gene. Genes in a segment-specific copy number of less than 0.5 were considered deep deletions (Deep DEL). To measure segment-specific copy number amplifications, the genome ploidy was subtracted for each sample to obtain the copy number above ploidy (CNAP). CNAPs of at least 3 are considered as amplifications (AMP); CNAPs above 1.5, but below 3 are considered low amplification (GAIN); CNAPs of at least 6 are considered high amplifications (High AMP), and CNAPs of at least 9 and no more than 100 genes are considered focal high amplification (Focal High AMP). In the figures, AMP or GAIN copy number amplifications were not included for any sample.

#### Significantly mutated genes

MutSig2CV^20^ was used to infer significantly recurrent mutated genes for specific sub cohorts. MutSig2CV uses patient- and gene-specific mutation rates to estimate a background model of predicted mutation incidence across the genome, and factors in biological covariates such as replication timing and gene-expression level on a gene-by-gene basis to account for the increased mutational rate of certain classes of genes. Genes were called significantly mutated if q<0.10. Mutations in cancer genes that were significantly mutated were manually checked to verify if they appeared to be artifacts and genes with a large number of mutations that appeared to be artifacts were removed.

#### Tumor mutational burden

Tumor mutational burden (TMB) was calculated as the total number of mutations (non-synonymous) detected for a given sample divided by the length of the total genomic target region captured with WES. A two-sided Wilcoxon test was used to calculate significance for differences in TMB across sub-cohorts.

#### Mutational signatures

SignatureAnalyzer^27^ was used to identify *de novo* mutational signatures in the cohort. deconstructSigs was used to calculate the strength of the COSMIC signatures for individual tumors^28^.

#### Tumor evolutionary analysis

Patients with >2 tumor or ctDNA WES samples collected from different timepoints/locations were used to study tumor evolution and tumor heterogeneity. To properly compare SNVs and indels in samples from the same patient, the union of all mutations called in each patient’s samples were considered. The reference and alternate reads in each patient’s samples were used as input for ABSOLUTE^23^ to compute cancer cell fractions. The clonal structure and the evolutionary history of the clones (phylogenic tree) was inferred with PhylogicNDT^24^ using only SNV sites and retaining only clones with cancer cell fraction (CCF) of more than 1%. For the evolutionary analysis in Table 3, mutations were denoted shared clonal if they had a CCF≥70% and a read count ≥3 in 2 or more samples, private clonal if they had a CCF≥70% and a read count ≥3 in only 1 sample, and acquired clonal if they had CCF≥20% and had a read count ≥3 in a sample and <3 reads in the reference sample.

### Germline analysis

Germline variants from blood and saliva WES samples were identified using GATK’s (version 4.1.4.0) haplotypecaller and joint genotyping module^52^. These variants were ranked for their likelihood of being pathogenic using a custom implementation of CharGer^30^. Variants with a CharGer score greater than 8 were considered pathogenic or likely pathogenic and only rare variants (population allele frequency < 0.05%, as in the TCGA germline study^30^) were considered. Pathogenic germline variants went through manual quality control and were removed if they appeared to be sequencing artifacts or did not pass the quality control filters of gnomAD v2.1.1. To compare the number of patients with germline variants in MBCproject with those from the TCGA germline study^30^, germline variants were obtained from the Supplemental Information, and filtered to those with a CharGer score greater than 8, resulting in 105 germline variants from 103/1076 patients. A Kolmogorov–Smirnov two-sided, two-sample test was used to compare the age distribution to evaluate the enrichment of pathogenic or likely pathogenic germline variants in age-based cohorts.

### Transcriptome sequencing data processing and analysis

#### Data processing and quality control

RNA-seq reads were mapped to the human genome (hg19) with STAR aligner^55^ with default parameters. Transcriptome quality was assessed using RNA-SeQC 2^56^ and expression quantification was conducted using RSEM^57^. Fusions were detected using the STAR-Fusion (version 1.8.1b) 39 for all tumor samples using default parameters. Fusion Annotator (https://github.com/FusionAnnotator/CTAT_HumanFusionLib/wiki) and Annotate Gene Fusion (AGFusion version 1.25)^58^ was used to annotate these fusions. Samples with <8,000 unique genes were removed from subsequent analysis. Gene expression was measured using log2(TPM+1) values.

#### Transcriptional signature activity

To calculate the activity of a transcriptional signature or gene set, we used single-sample gene set enrichment analysis (ssGSEA). ssGSEA was performed for all tumor samples using fgsea^59^ to calculate normalized enrichment scores for Hallmark gene sets from the Molecular Signatures Database^36^ using upper quartile normalized expression values.

#### PAM50 molecular subtype assignment

To assign research-based PAM50 subtypes, expression values were rescaled relative to those of a receptor status-balanced Metastatic Breast Cancer Project (MBCproject) cohort, in which samples were re-sampled to achieve the ER-positive to ER-negative receptor status ratio in the UNC training set, from which the PAM50 subtype centroids were derived^60, 61^. genefu was used to call research-based PAM50 subtypes^62^ using the rescaled expression values and spearman correlation to the PAM50 subtype centroids. Samples with a PAM50 centroid correlation <0.10 for each centroid were not assigned a PAM50 subtype (Not Classified, NC).

### Statistical tests and analysis

A *P* value of < 0.05 was considered to be statistically significant. A two-sided Fisher exact test was used to calculate significance for univariate frequency comparisons. Additional statistical tests used are described in the methods subsection of each data type. All statistical analysis was performed using R (version 4.0.3).

## Data availability

The clinically annotated genomic dataset of the MBCproject is shared publicly in order for all researchers to be able to utilize this data to better understand metastatic breast cancer. De-identified data has been shared on a recurring basis as the data is generated. Data from The Metastatic Breast Cancer Project is available at cBioPortal (https://www.cbioportal.org/study/summary?id=brca_mbcproject_2022), National Cancer Institute’s Genomic Data Commons (https://portal.gdc.cancer.gov/projects/CMI-MBC) and dbGaP (Study Accession phs001709). cBioPortal has the clinical and genomic data for the MBCproject clinico-genomic dataset (N=379 tumor, 301 patients). As of May 1st 2023, updating the Genomic Data Commons and dbGaP data repositories with the genomic data for all the tumor samples used in this study is in progress.

## Supporting information

Supplemental Material descriptions

## Acknowledgements

This study would not have been possible without the many metastatic breast cancer patients and loved ones of patients who have worked in deep partnership on this research project for many years. Through collaborations at conferences, your visits to the Broad Institute, and all of the interactions that we have had with you over the years, we are forever grateful to you for your generosity, knowledge, and enthusiasm. Thank you for always inspiring and educating us with your ideas, stories, and experiences. We are grateful to all of the advocacy partners who have worked together with us on the MBCproject. We appreciate many people from across the Broad Institute and Dana Farber for helpful support and scientific discussions. We thank the Broad Institute Communications, Development, & Compliance teams for their work in support of the MBCproject. We are deeply indebted to all former and current members of the Count Me In Team, the Wagle laboratory, the Broad Institute engineering team, the Broad Cancer Program, and the Broad Genomics Platform for their efforts to make this work possible. This research was supported by the non-profit organization Count Me In (joincountmein.org) and by anonymous philanthropic support to the Broad Institute.

## List of Supplemental Material

**Supplemental Figures S1-S12. Supplemental Tables S1-S12. Supplemental Files 1-2.**

