## Supplemental Material descriptions for "The Metastatic Breast Cancer Project: leveraging patient-partnered research to expand the clinical and genomic landscape of metastatic breast cancer and accelerate discoveries"

### Supplemental Text

**Supplemental Text. Additional information on The Metastatic Breast Cancer Project and Supplemental Methods.**

### Supplemental Figures

**Supplemental Figure S1. Key steps for carrying out patient-partnered research in MBCproject.** Overview of the key steps (panel A) and their application to develop a process for remote collection of blood samples from participants. (panel B).

**Supplemental Figure S2. Sources from which participants learned about MBCproject.** Patient-reported response to the question, "How did you hear about the MBCproject?" in the intake survey. Note: This question was added in January 2019. Participants who registered prior to that date did not have this question in their intake survey.

**Supplemental Figure S3. Process overview of all steps in the MBCproject required for data deposition, including attrition at each step** (A) Overview of MBCproject processes. Steps outlined in orange represent steps carried out by patients, while steps outlined in green indicate steps completed by the study team. (B) Data types available for each patient in the clinico-genomic dataset that has a tumor sample with exome data (N = 379 samples, 301 patients). Each box denotes whether the patient has the specified data type. The color of the box denotes either if the data type is available or the number of samples sequenced for each patient. MRD: Medical Record Data; PRD: Patient Reported Data.

**Supplemental Figure S4. Distribution of the following characteristics for all patients included in the clinico-genomic dataset: Age at primary diagnosis, time difference between primary and metastatic diagnosis, and time difference between primary diagnosis and biopsy collection.** (A) Distribution of patients based on their age at primary breast cancer diagnosis. (B) Distribution of patients based on the time difference between their primary and metastatic breast cancer diagnosis. A difference of ≤4 months between primary and metastatic diagnosis is considered *de novo* metastatic breast cancer. (C) Distribution of time difference between primary breast cancer diagnosis and biopsy collection.

**Supplemental Figure S5. Distribution of receptor status obtained from the medical records for each sample with exome data in the clinico-genomic dataset (N = 379 samples, 301 patients).** (A) Distribution for the estrogen receptor (ER). (B) Distribution for the progesterone receptor (PR). (C) Distribution for the HER2 receptor. ctDNA samples (N=104) do not have an associated receptor status.

**Supplemental Figure S6. Concordance between medical record data (MRD) and patient reported data (PRD) for MBCproject participants.** (A) Percentage of concordance/discordance for each clinical parameter. (B) Difference in date of birth. (C) Difference in date of primary breast cancer diagnosis. (D) Difference in date of metastatic breast cancer diagnosis. (E) Difference in time to metastatic breast cancer diagnosis. (F) Precision and sensitivity of PRD data of top 10 most commonly received treatment drugs based on MRD data.

**Supplemental Figure S7. Tumor purity and ploidy for samples with exome data and tumor fraction for ULP-WGS cfDNA samples.** (A) Distribution of tumor samples based on purity. (B) Tumor purity of samples based on tumor location. (C) Distribution of tumor samples based on ploidy. (D) Purity and ploidy for each sample. (E) Distribution of ULP-WGS cfDNA samples based on tumor fraction.

**Supplemental Figure 8. Comparison of the mutational frequency of cancer genes between tumors in the treatment-naive primary setting and treatment-exposed metastatic setting.** (A) Frequency of gene mutations in samples taken in the treatment-naive primary setting versus treatment-exposed metastatic setting. (B) Frequency of gene mutations in samples taken in the treatment-naive primary setting versus treatment-exposed distant metastatic setting. Treatment-naive primary setting is defined as breast and breast-adjacent (axilla or axillary lymph nodes) samples taken prior to diagnosis with metastatic disease that have not been exposed to systemic treatment. Treatment-exposed metastatic is defined as any sample, including breast and axilla samples, obtained on or after metastatic disease diagnosis date that have been exposed to systemic treatment. Treatment-exposed distant metastatic is defined as any sample, other than breast or axilla samples, obtained on or after metastatic disease diagnosis.

**Supplemental Figure S9. Mutational signatures and tumor mutational burden (TMB) in the clinico-genomic dataset with exome data (N = 379 samples, 301 patients).** (A) Distribution of tumor samples based on their dominant mutational signature. (B) Score of COSMIC mutational signatures of samples taken before or after metastatic diagnosis. (C) Score of COSMIC mutational signatures of treatment naive samples taken before metastatic diagnosis and non-treatment naive samples taken after metastatic diagnosis. (D) Tumor mutational burden of samples taken before or after metastatic diagnosis. (E) Tumor mutational burden of treatment naive samples taken before metastatic diagnosis and non-treatment naive samples taken after metastatic diagnosis. (F) Tumor mutational burden of samples before and after metastatic diagnosis based on the dominant mutational signature in each sample.

**Supplemental Figure S10. Quality control metrics, metastatic setting, and distribution of number of fusions detected per tumor for the MBCproject transcriptomic dataset (N = 200, 141 patients).** (A) Distribution of number of genes detected. The 200 tumor samples that passed the quality control threshold of 8000 genes detected are shown. (B) Metastatic disease setting for the tumor samples. Metastatic setting is based on whether the sample was obtained before or after metastatic diagnosis. (C) Distribution of number of genes detected based on biopsy location. (D) Distribution of tumor samples based on the number of fusions identified.

**Supplemental Figure S11. In-frame fusions in kinase genes and putatively novel in-frame fusions.** (A) In-frame fusions in kinase genes. (B) In-frame fusions in cancer genes not found in fusion databases (based on FusionAnnotator and OncoKB). Each line of unit width denotes a unique gene fusion and the ends of the line point to the gene pairs in the fusion.

**Supplemental Figure S12. Distribution of transcriptional subtypes and *ESR1* expression by hormone receptor status and transcriptional subtype.** (A) Distribution of tumor samples based on transcriptional subtypes (research-based PAM50). (B) *ESR1* expression based on hormone receptor status (HR) status. (C) *ESR1* expression based on transcriptional subtype. LumA, Luminal A; LumB, Luminal B; HER2-E, HER2-enriched; NC, Not classified.

### Supplemental Tables

**Supplemental Table S1 Advocacy Partner Organizations.** List of advocacy organizations with which the MBCproject partners, including the organization name and the website URL.

**Supplemental Table S2. Patient-reported breast cancer details (N=3246).** (A) Patient reported data from intake survey questions about receptor and inflammatory status. Distribution of responses for each question. (B) Patient-reported response to the question, “Was your breast cancer identified as any of the following at any time (select all that apply)?” in the follow-up survey. Participants were able to make multiple selections from the list. (C) Patient reported data from intake survey about treatments in the metastatic setting taken for a duration of over 2 years and/or that resulted in non-evaluable disease or a dramatic reduction in tumor size. Distribution of responses for each question.

**Supplemental Table S3. Demographic information.** (A) Patient-reported response to ethnicity question. 3220 of 3246 consented participants completed the question. (B) Patient-reported response to the question, “What is your race?”. Distribution of responses. Participants were able to make multiple selections from the list.

**Supplemental Table S4. Clinical drug ontology database.** Clinical drug ontology database to categorize treatments received by metastatic breast cancer patients as reported in medical records and/or surveys. Preferred drug category is defined based on the drug categories available and the level of detail relevant for the context of metastatic breast cancer treatment, and is the category used .

**Supplemental Table S5. Comparison of Medical Record Data (MRD) with Patient Reported Data (PRD).** (A) Comparison of dates of birth, primary diagnosis date, metastatic diagnosis date, and time from primary diagnosis to metastatic diagnosis as submitted via survey and listed in medical record. (B) Comparison of receptor status (HR-positive, HER2-positive, and triple negative) and histology (IDC/ILC and mixed IDLC). Note that in their survey, patients selected receptor status and histology from any time in their diagnosis, which was compared to all receptor and histological types listed in the medical record. (C) Cohort based query comparisons of metastatic sites at metastatic diagnosis (bone only metastases, visceral metastatic sites, and brain metastases) as submitted in survey and listed in medical record

**Supplemental Table S6. Serial tumor whole-exome sequencing (N=379 tumors, 301 patients).** (A) Number of tumor samples per patient with whole-exome-sequencing (WES). (B) Number of patients with one or more tumor WES samples per number of timepoints (times at which a sample was obtained during a patient’s clinical history).

**Supplemental Table S7. Comparison of dominant mutational signatures based on metastatic setting and treatment exposure.** (A) Comparison of the dominant *de novo* mutational signature of tumors based on their metastatic setting. (B) Comparison of the dominant *de novo* mutational signature of treatment naive tumors taken before metastatic diagnosis and treatment exposed tumors taken after metastatic diagnosis. A two-sided Fisher exact test was used for the comparisons in panels A and B.

**Supplemental Table S8. Comparison of the prevalence of pathogenic/likely-pathogenic germline variants in cancer predisposition genes based on the age at primary diagnosis.** (A) Comparison of the age distribution at primary diagnosis of patients with or without pathogenic/likely-pathogenic germline variants in *BRCA1* or *BRCA2*, or cancer predisposition genes from the TCGA germline study. A two-sided, two-sample Kolmogorov–Smirnov test was used for the comparisons. (B) Comparison of the frequency of pathogenic/likely-pathogenic germline variants based on the patient's age at primary diagnosis (before/at or after the age of 40 years old). A two-sided Fisher exact test was used for the comparisons.

**Supplemental Table S9. Serial tumor RNA sequencing.** (A) Number of tumor samples per patient with RNA sequencing (RNA-seq) from the dataset consisting of 200 tumors from 141 patients. (B) Number of patients with one or more tumor RNA-seq samples per number of timepoints (times at which a sample was obtained during a patient's clinical history). Samples for which either the date it was obtained or the date of primary diagnosis is unknown are not included in the timepoint table for the cases when more than 1 timepoint is available per patient.

**Supplemental Table S10. In-frame fusions in kinase genes and in cancer genes but not in fusion databases.** (A) In-frame fusions involving a kinase gene classified as oncogenic or likely-oncogenic by OncoKB. (B) In-frame fusions in cancer genes not found in cancer-related fusion databases (based on FusionAnnotator and OncoKB). Fusions not explicitly found in OncoKB can still be assigned an alteration effect by OncoKB.

**Supplemental Table S11. Comparison of transcriptional subtypes between MBCproject and TCGA, and difference in transcriptional subtype in paired tumor samples.** (A) Comparison of the frequency in transcriptional subtypes (research-based PAM50) between the MBCproject dataset and TCGA BRCA. (B) Difference in transcriptional subtype in paired patient tumor samples. Paired tumor samples from the 15 out of 35 patients in which there was a difference in transcriptional subtype. Clinical metadata associated with these samples (reference tumor sample used for the comparison, time between primary diagnosis and sample collection, time difference between samples, biopsy site, and receptor subtype) is included. MBCproject: Metastatic Breast Cancer Project; TCGA BRCA: The Cancer Genome Atlas Breast Invasive Carcinoma.

**Supplemental Table S12. Clinical and genomic information for patient 3717's tumor samples.** Clinical features included are metastatic setting, tumor site, time relative to primary diagnosis, and receptor status. Genomic features included are putative resistance driver mutations (from whole-exome sequencing), putative resistance driver fusions, research-based PAM50, *ESR1* expression, and ER transcriptional signature activity (from RNA sequencing). Interquartile range (IQR) for *ESR1* expression and ER transcriptional signature activity for the ER-positive and ER-negative tumors are included as reference.

### Supplemental Files

**Supplemental File 1. MBCproject forms, surveys, and request templates.** The forms contained are the following: (i) tissue consent form, (ii) blood consent form, and (iii) release form. The surveys contained are the following: (i) intake survey (version 1), (ii) intake survey (version 1), (iii) intake survey (version 1), and (iv) follow-up survey. The request forms contained are the following: (i) medical record request template, (ii) pathology report request template, and (iii) tissue request template.

**Supplemental File 2. Data Dictionary.** Data dictionary model used for medical record data abstraction for 40 different clinical variables.
