## Supplemental Figures for "The Metastatic Breast Cancer Project: leveraging patient-partnered research to expand the clinical and genomic landscape of metastatic breast cancer and accelerate discoveries"

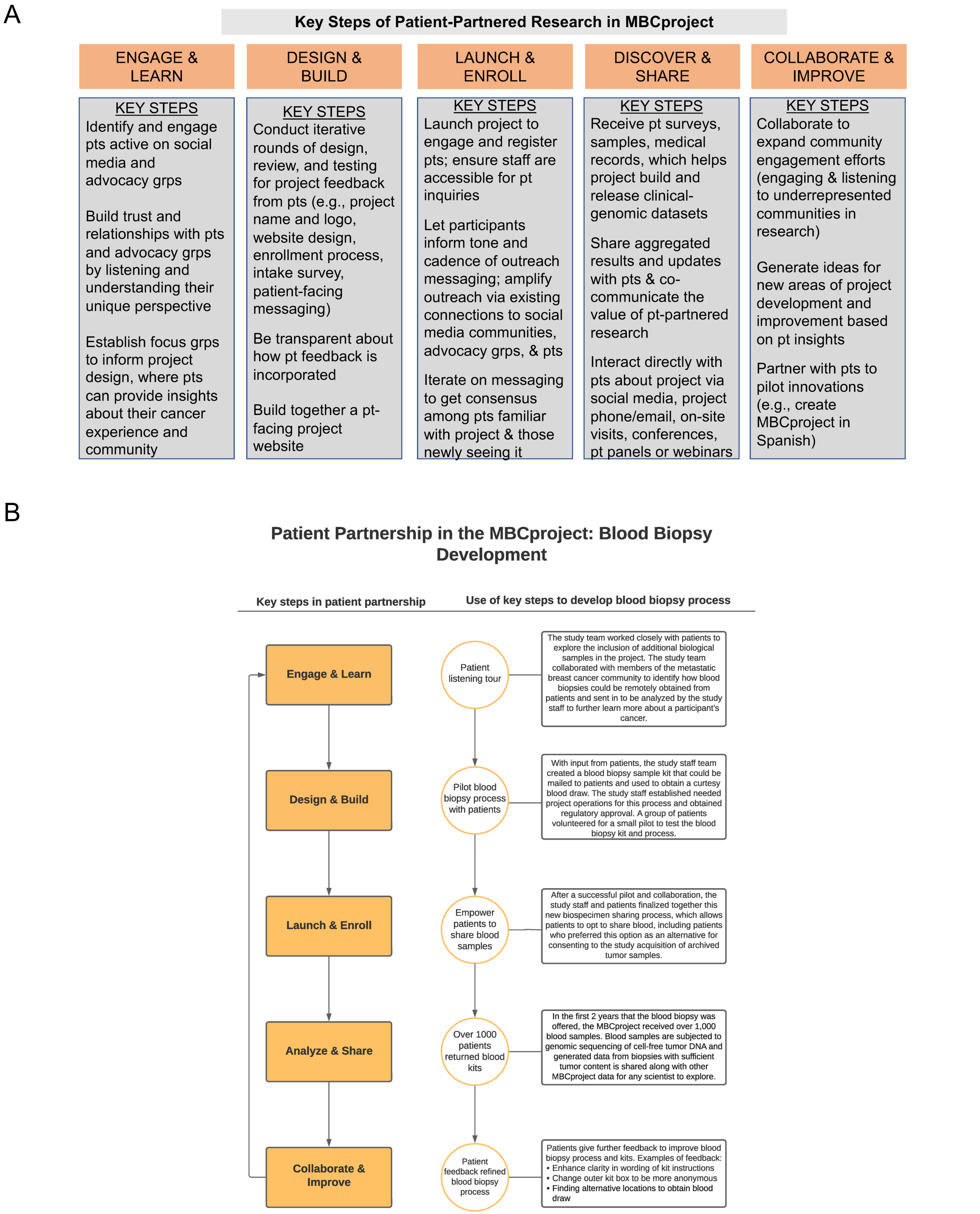


**Supplemental Figure S1. Key steps for carrying out patient-partnered research in MBCproject.** Overview of the key steps (panel A) and their application to develop a process for remote collection of blood samples from participants. (panel B).

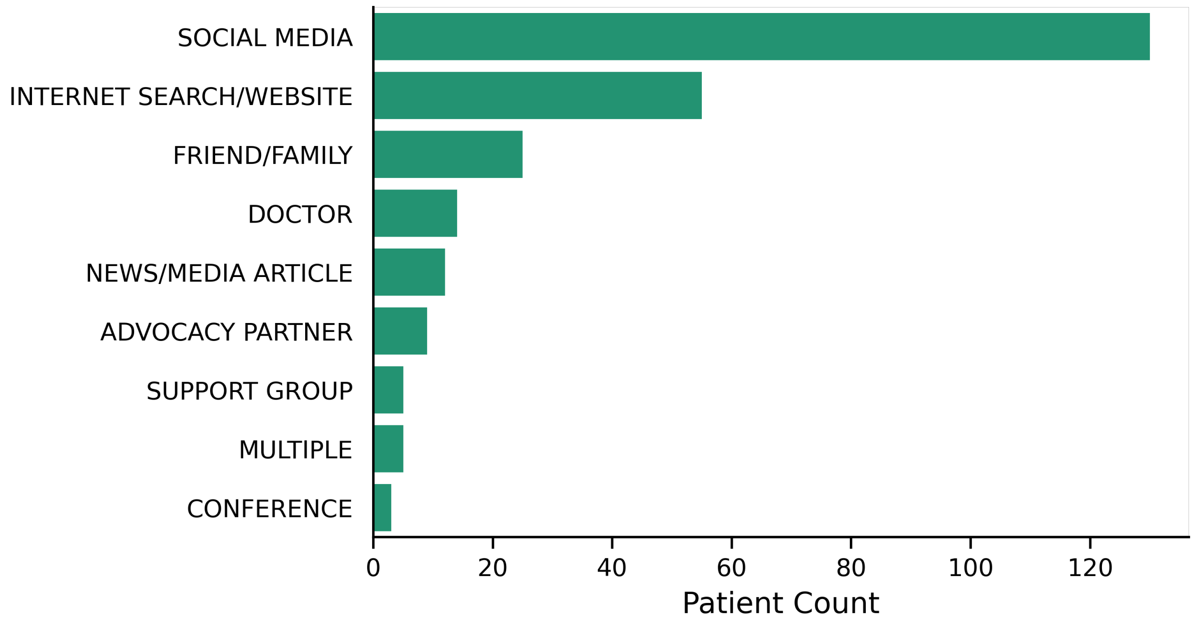


**Supplemental Figure S2. Sources from which participants learned about MBCproject.** Patient-reported response to the question, "How did you hear about the MBCproject?" in the intake survey. Note: This question was added in January 2019. Participants who registered prior to that date did not have this question in their intake survey.

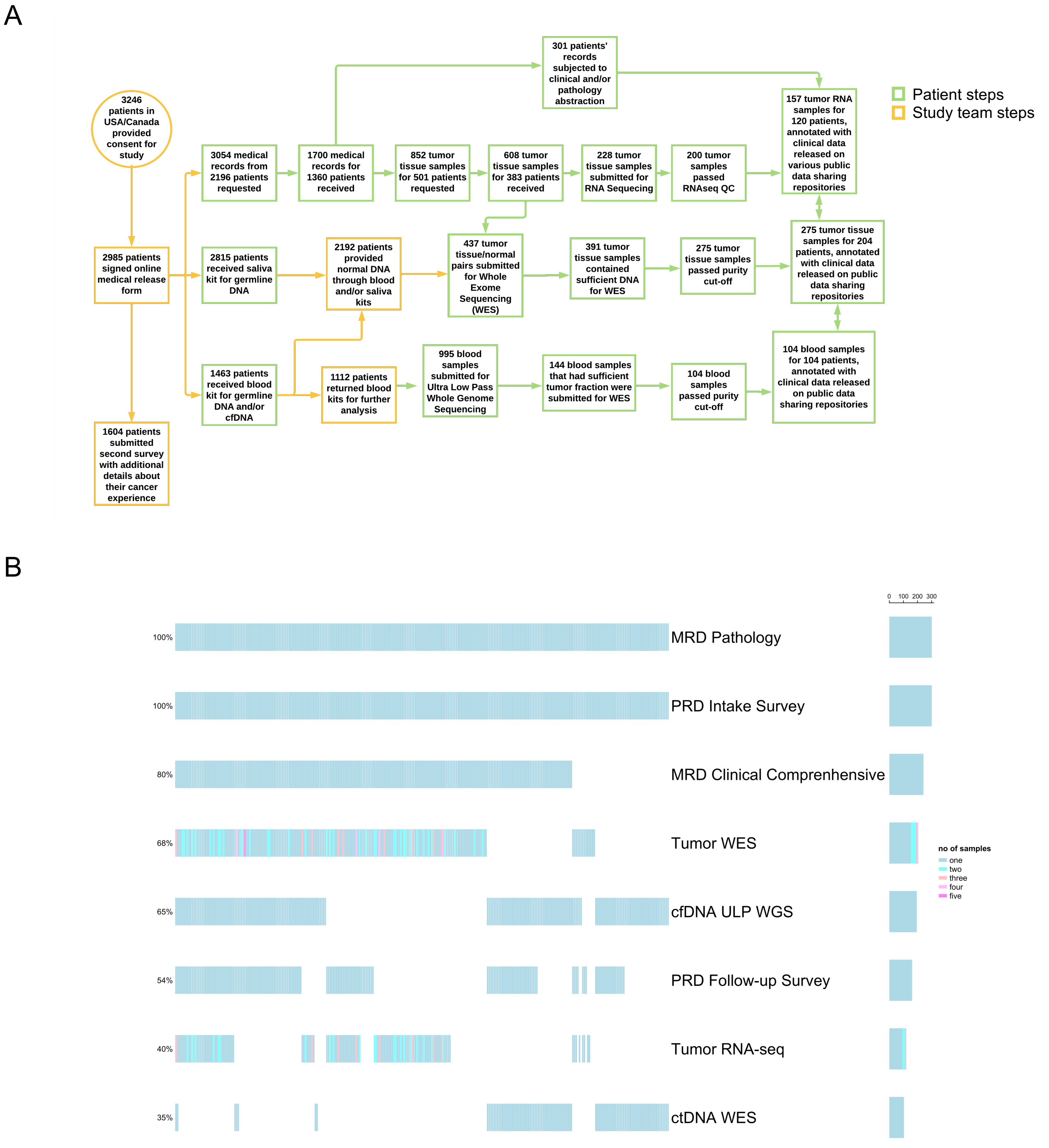


**Supplemental Figure S3. Process overview of all steps in the MBCproject required for data deposition, including attrition at each step** (A) Overview of MBCproject processes. Steps outlined in orange represent steps carried out by patients, while steps outlined in green indicate steps completed by the study team. (B) Data types available for each patient in the clinico-genomic dataset that has a tumor sample with exome data (N = 379 samples, 301 patients). Each box denotes whether the patient has the specified data type. The color of the box denotes either if the data type is available or the number of samples sequenced for each patient. MRD: Medical Record Data; PRD: Patient Reported Data.

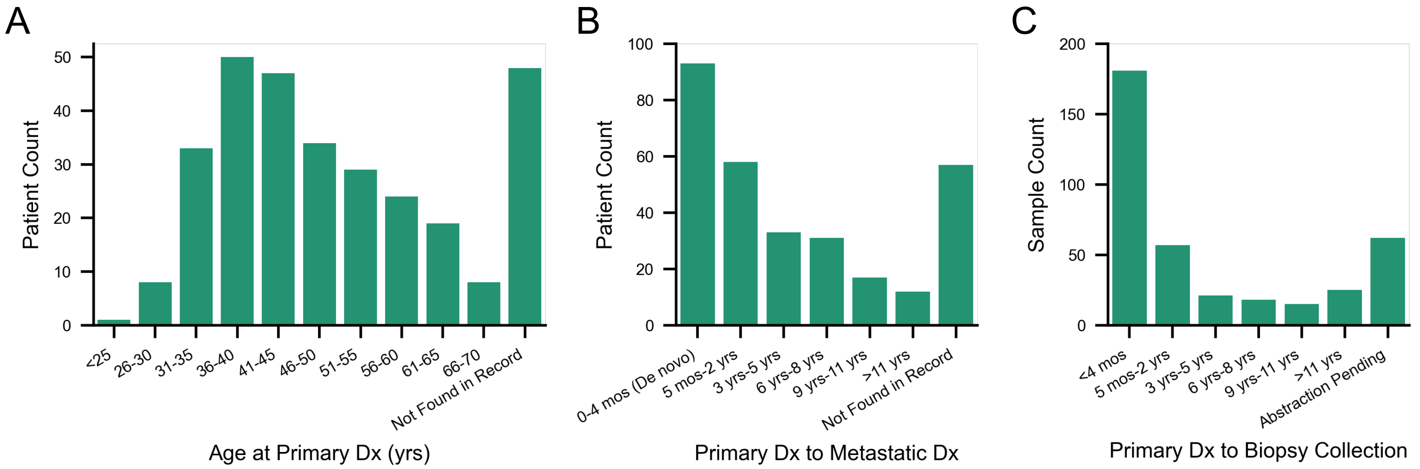


**Supplemental Figure S4. Distribution of the following characteristics for all pts included in the clinico-genomic dataset: Age at primary diagnosis, time difference between primary and metastatic diagnosis, and time difference between primary diagnosis and biopsy collection.** (A) Distribution of patients based on their age at primary breast cancer diagnosis. (B) Distribution of patients based on the time difference between their primary and metastatic breast cancer diagnosis. A difference of ≤4 months between primary and metastatic diagnosis is considered *de novo* metastatic breast cancer. (C) Distribution of time difference between primary breast cancer diagnosis and biopsy collection.

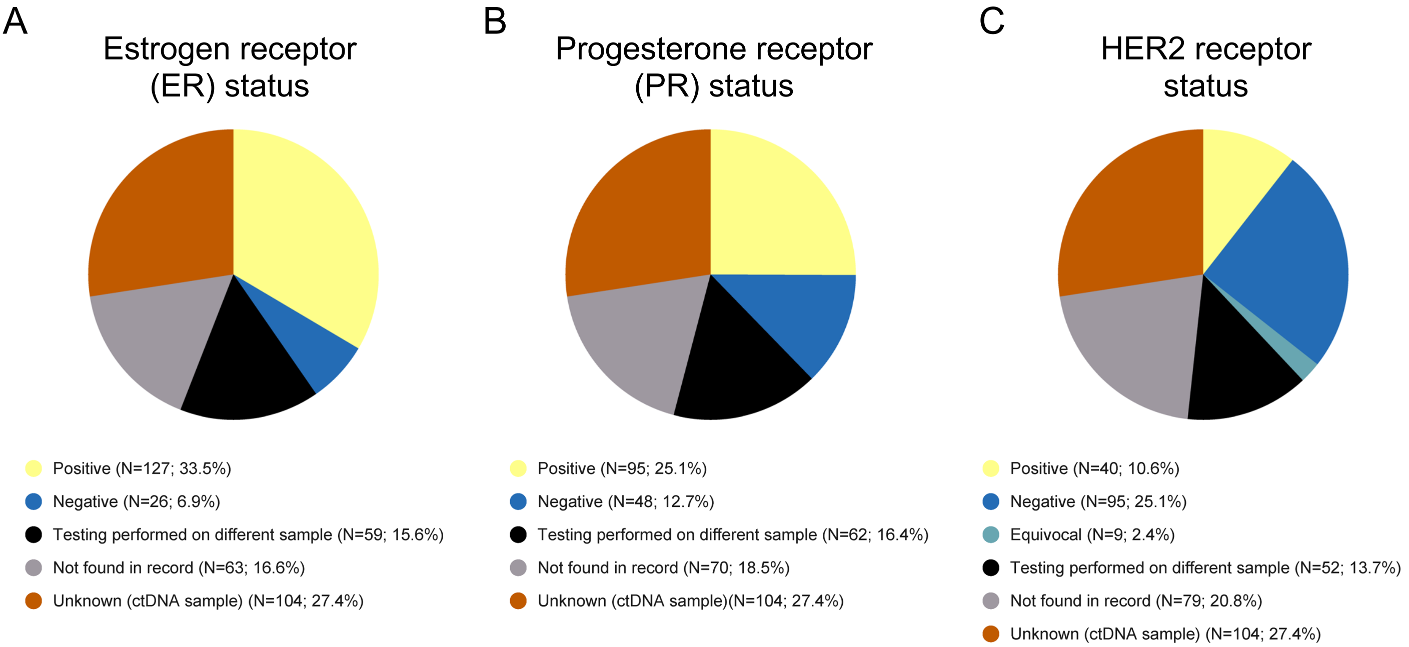


**Supplemental Figure S5. Distribution of receptor status obtained from the medical records for each sample with exome data in the clinico-genomic dataset (N = 379 samples, 301 patients).** (A) Distribution for the estrogen receptor (ER). (B) Distribution for the progesterone receptor (PR). (C) Distribution for the HER2 receptor. ctDNA samples (N=104) do not have an associated receptor status.

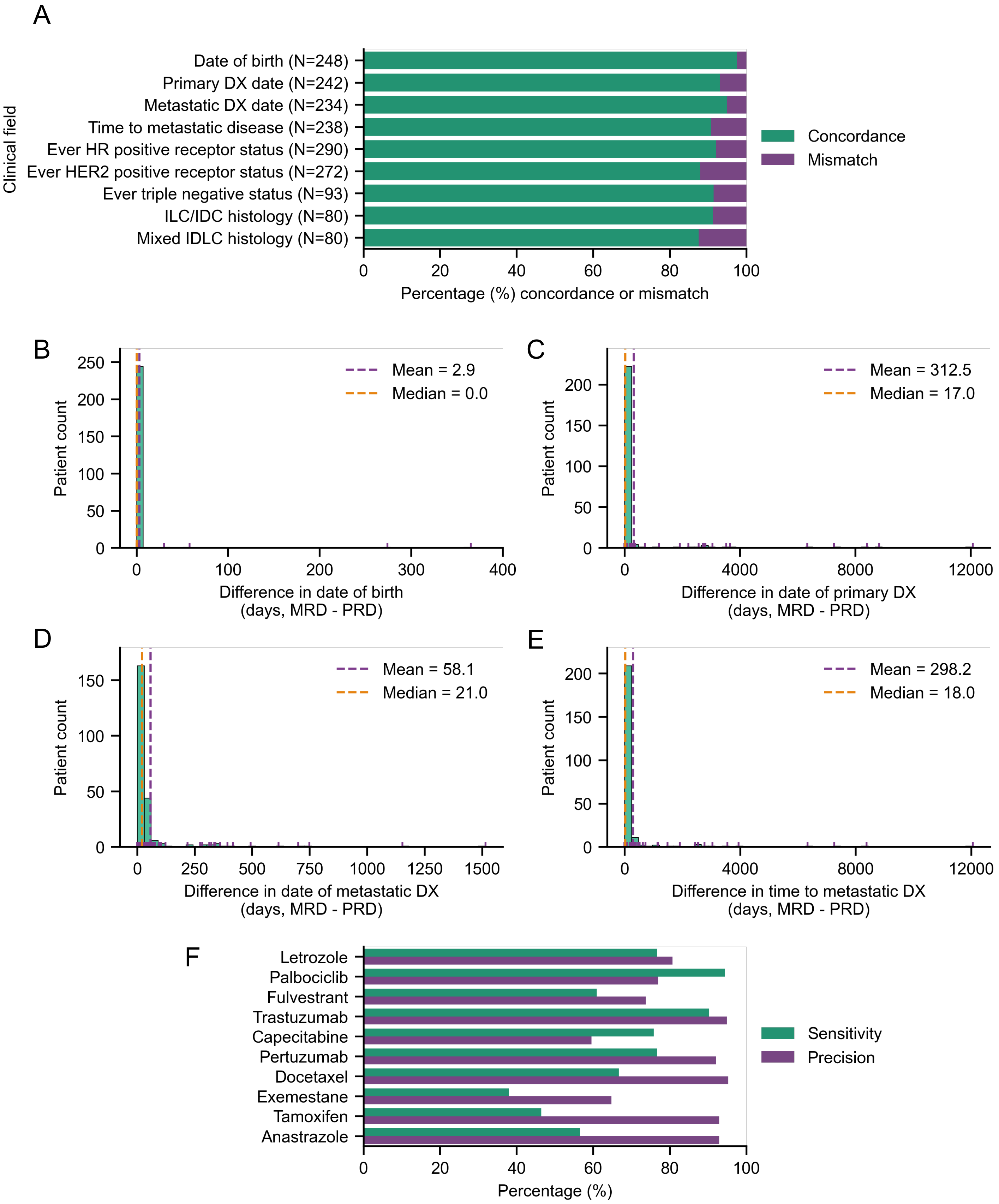


**Supplemental Figure S6. Concordance between medical record data (MRD) and patient reported data (PRD) for MBCproject participants.** (A) Percentage of concordance/discordance for each clinical parameter. (B) Difference in date of birth. (C) Difference in date of primary breast cancer diagnosis. (D) Difference in date of metastatic breast cancer diagnosis. (E) Difference in time to metastatic breast cancer diagnosis. (F) Precision and sensitivity of PRD data of top 10 most commonly received treatment drugs based on MRD data.

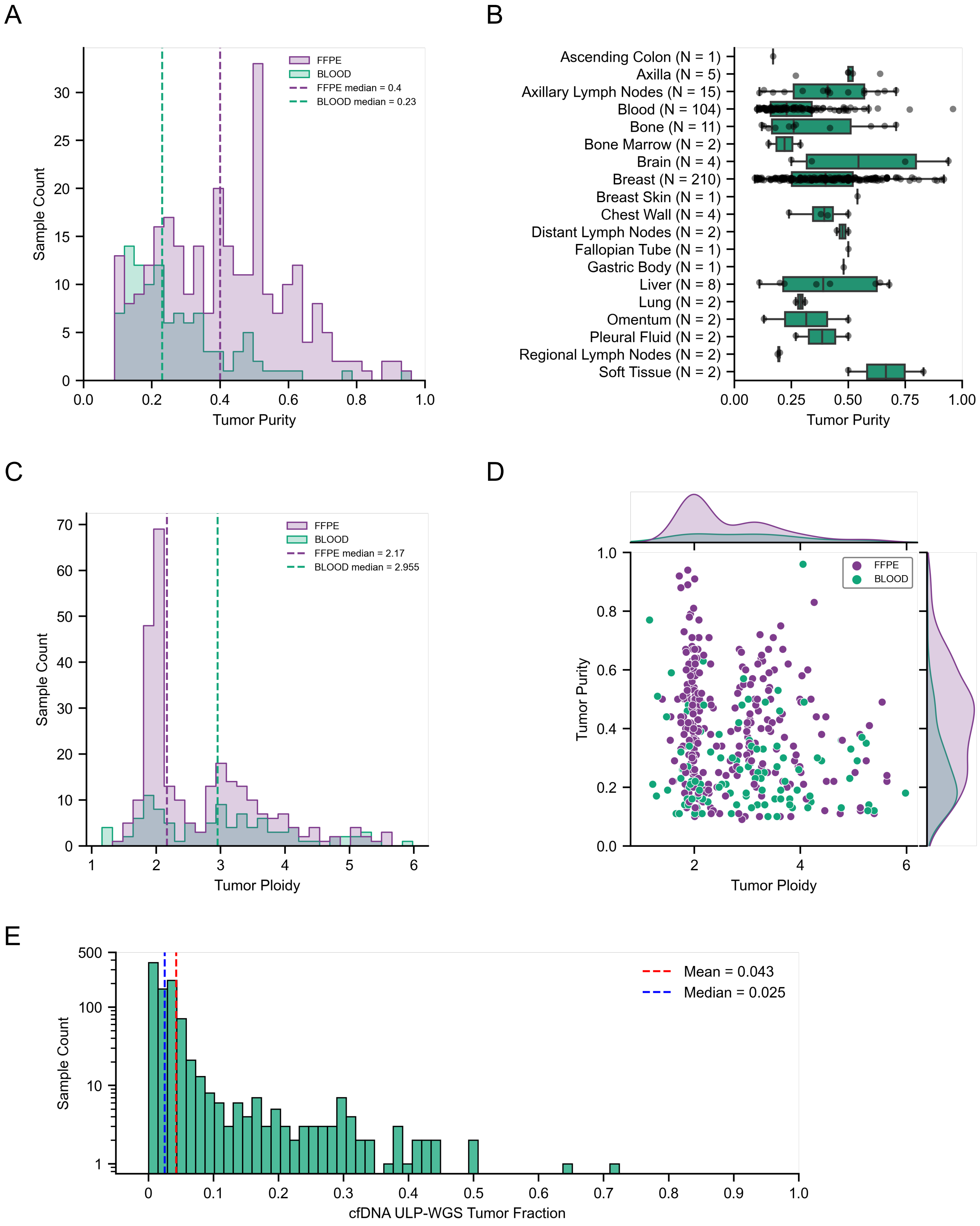


**Supplemental Figure S7. Tumor purity and ploidy for samples with exome data and tumor fraction for ULP-WGS cfDNA samples.** (A) Distribution of tumor samples based on purity. (B) Tumor purity of samples based on tumor location. (C) Distribution of tumor samples based on ploidy. (D) Purity and ploidy for each sample. (E) Distribution of ULP-WGS cfDNA samples based on tumor fraction.

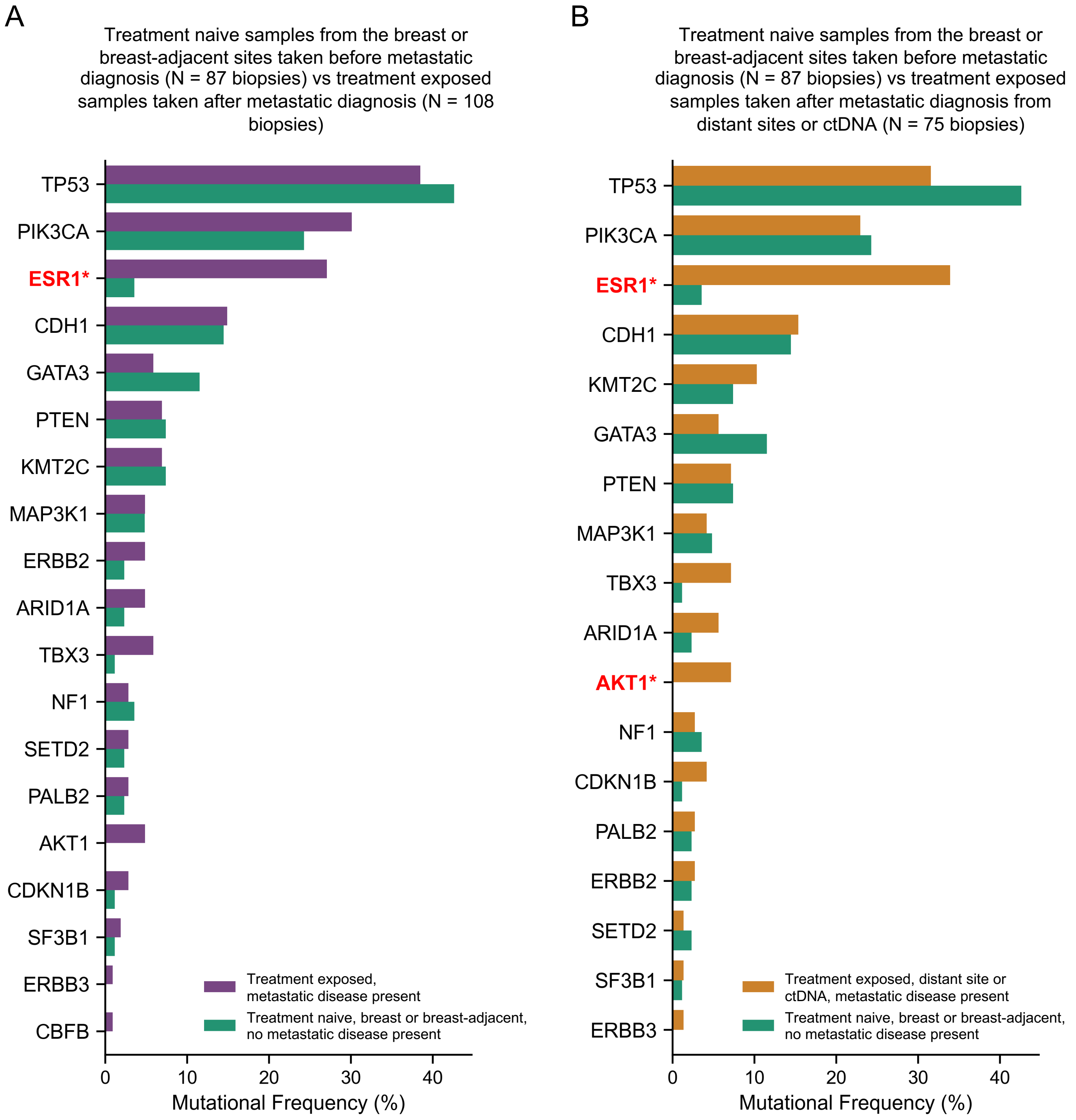


**Supplemental Figure 8. Comparison of the mutational frequency of cancer genes between tumors in the treatment-naive primary setting and treatment-exposed metastatic setting.** (A) Frequency of gene mutations in samples taken in the treatment-naive primary setting versus treatment-exposed metastatic setting. (B) Frequency of gene mutations in samples taken in the treatment-naive primary setting versus treatment-exposed distant metastatic setting. Treatment-naive primary setting is defined as breast and breast-adjacent (axilla or axillary lymph nodes) samples taken prior to diagnosis with metastatic disease that have not been exposed to systemic treatment. Treatment-exposed metastatic is defined as any sample, including breast and axilla samples, obtained on or after metastatic disease diagnosis date that have been exposed to systemic treatment. Treatment-exposed distant metastatic is defined as any sample, other than breast or axilla samples, obtained on or after metastatic disease diagnosis.

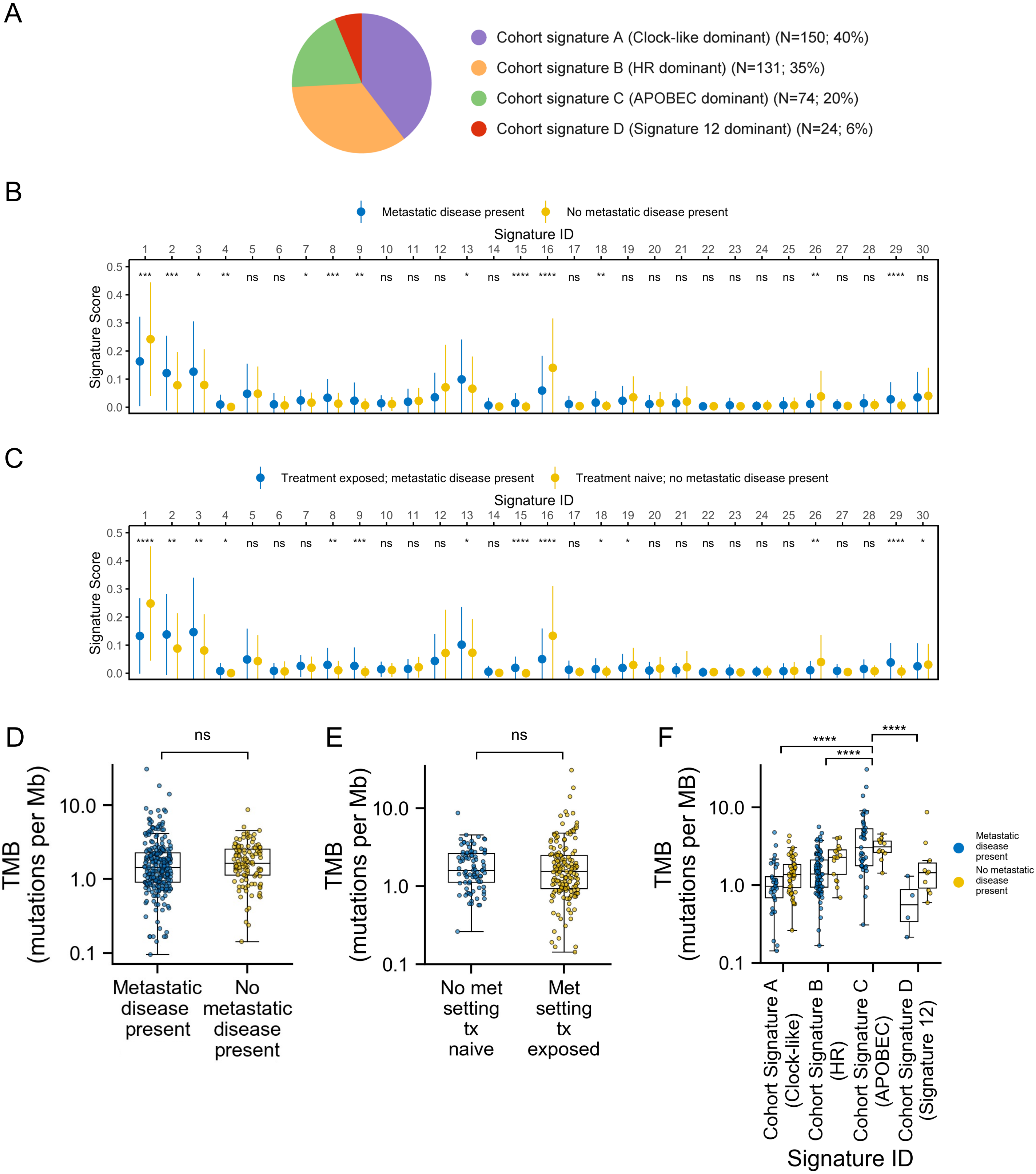


**Supplemental Figure S9. Mutational signatures and tumor mutational burden (TMB) in the clinico-genomic dataset with exome data (N = 379 samples, 301 patients).** (A) Distribution of tumor samples based on their dominant mutational signature. (B) Score of COSMIC mutational signatures of samples taken before or after metastatic diagnosis. (C) Score of COSMIC mutational signatures of treatment naive samples taken before metastatic diagnosis and non-treatment naive samples taken after metastatic diagnosis. (D) Tumor mutational burden of samples taken before or after metastatic diagnosis. (E) Tumor mutational burden of treatment naive samples taken before metastatic diagnosis and non-treatment naive samples taken after metastatic diagnosis. (F) Tumor mutational burden of samples before and after metastatic diagnosis based on the dominant mutational signature in each sample.

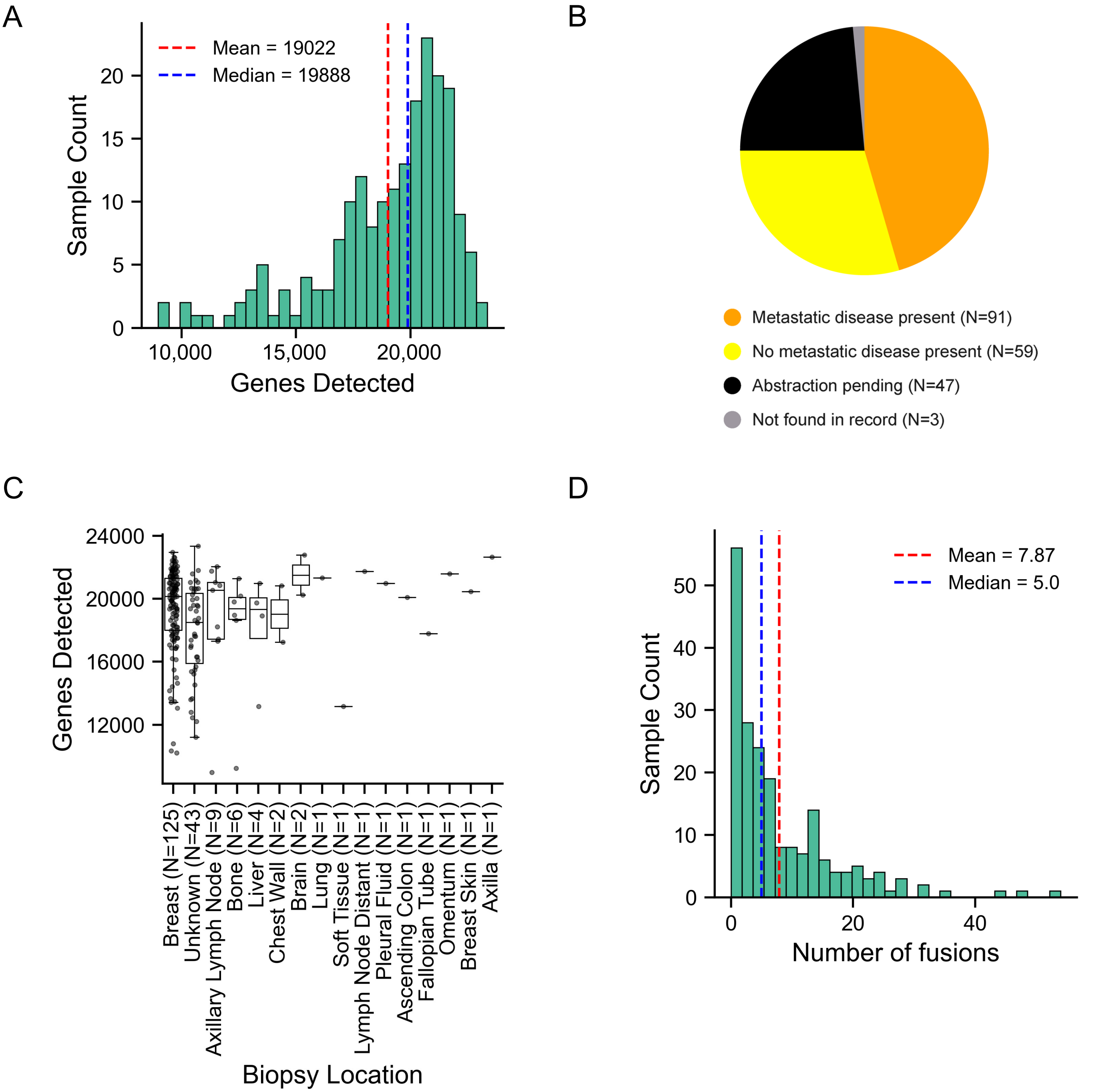


**Supplemental Figure S10. Quality control metrics, metastatic setting, and distribution of number of fusions detected per tumor for the MBCproject transcriptomic dataset (N = 200, 141 patients).** (A) Distribution of number of genes detected. The 200 tumor samples that passed the quality control threshold of 8000 genes detected are shown. (B) Metastatic disease setting for the tumor samples. Metastatic setting is based on whether the sample was obtained before or after metastatic diagnosis. (C) Distribution of number of genes detected based on biopsy location. (D) Distribution of tumor samples based on the number of fusions identified.

A

In-frame fusions in kinase genes

B

In-frame fusions in cancer genes not found in fusion databases

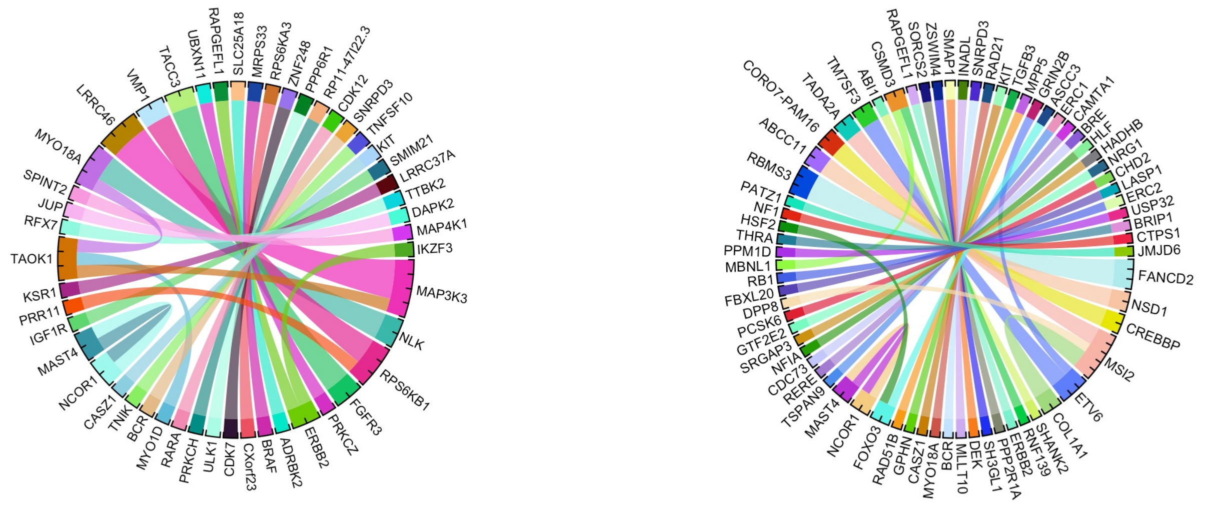


**Supplemental Figure S11. In-frame fusions in kinase genes and putatively novel in-frame fusions.** (A) In-frame fusions in kinase genes. (B) In-frame fusions in cancer genes not found in fusion databases (based on FusionAnnotator and OncoKB). Each line of unit width denotes a unique gene fusion and the ends of the line point to the gene pairs in the fusion.

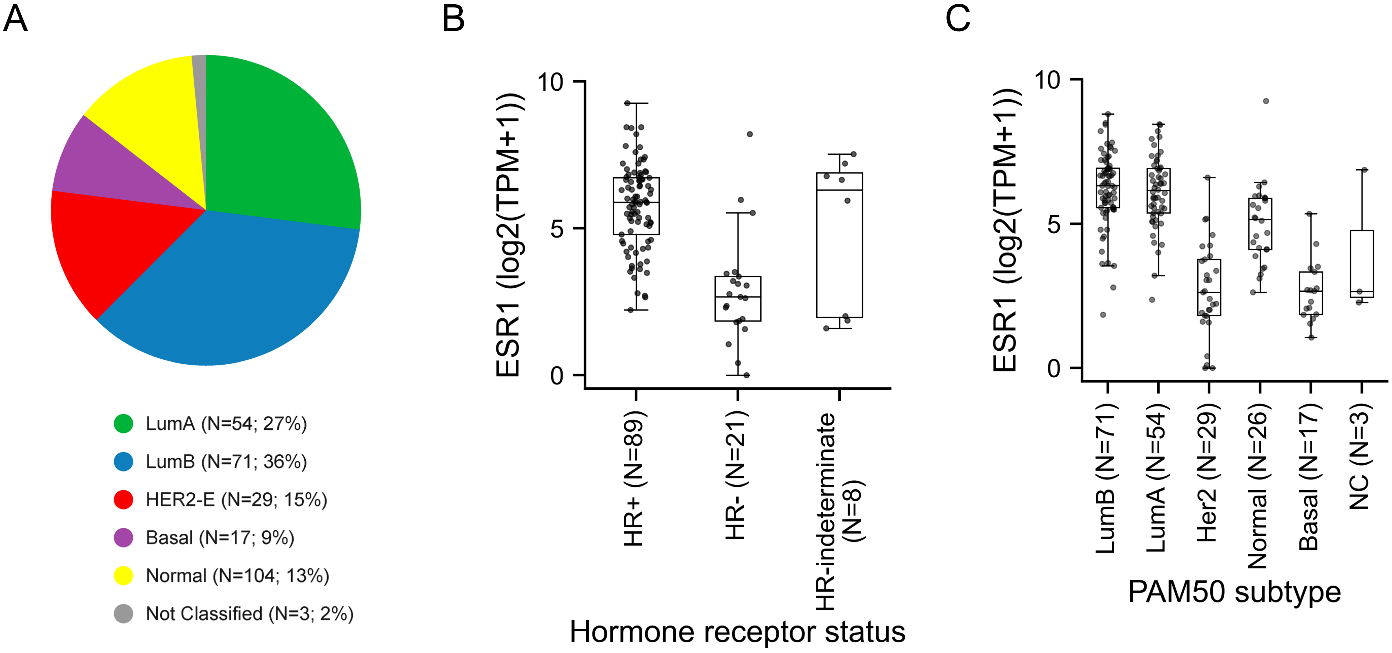


**Supplemental Figure S12. Distribution of transcriptional subtypes and *ESR1* expression by hormone receptor status and transcriptional subtype.** (A) Distribution of tumor samples based on transcriptional subtypes (research-based PAM50). (B) *ESR1* expression based on hormone receptor status (HR) status. (C) *ESR1* expression based on transcriptional subtype. LumA, Luminal A; LumB, Luminal B; HER2-E, HER2-enriched; NC, Not classified.
