## Supplemental Text for "The Metastatic Breast Cancer Project: leveraging patient-partnered research to expand the clinical and genomic landscape of metastatic breast cancer and accelerate discoveries"

### Supplemental Text. Additional information on The Metastatic Breast Cancer Project and Supplemental Methods

The Metastatic Breast Cancer Project (MBCproject) is ongoing and continues to enroll patients. The analyses conducted for this manuscript were performed using information and samples from patients who consented between September 15 , 2015 and March 31, 2020.

#### Patient and public involvement

A core pillar of the Metastatic Breast Cancer Project is patient engagement. There are several ways the Count Me In team engages with patients. These mechanisms include social media, through trusted advocacy organizations, and – for patients who want to be more deeply engaged – through Project Advisory Councils. In addition, Count Me In and the Metastatic Breast Cancer Project have several efforts aimed at connecting with members of the community who have been traditionally underrepresented in research to listen, learn, and build relationships.

Engagement on social media includes having a presence on Facebook, Twitter, and Instagram to share information about the project including updates on enrollment and general research findings, spreading awareness for conferences and/or events, and educating the public on how the project works. In addition to our own channels, patients often share information in private, community-specific groups.

Advocacy partnerships are another important engagement tool. The advocacy organizations that are project partners are trusted groups in the metastatic breast cancer community. By having these groups as partners, patients can have increased confidence that the project is a reputable research option for them. Partnerships involve a bidirectional relationship that fosters sharing of information when possible and appropriate. Partners share information about the MBCproject where it fits into their overall strategy including at conferences and events, via support groups, through blurbs in newsletters, and by posting on their social media platforms.

Project Advisory Councils (PACs) are an important aspect of patient engagement. PACs consist of patients, caregivers and advocates that are part of the metastatic breast cancer community. The goal of PACs is to get feedback on project design, including website language and tone, clarity on project processes, reactions to physical materials, and the build of new processes. Feedback is gathered via Google feedback surveys, email and phone interactions, and group meetings. Informal feedback from any and all project stakeholders is encouraged and often received organically via email messages and phone calls.

The Metastatic Breast Cancer Project and the Count Me In community engagement team has several efforts in place to reach historically marginalized and underrepresented communities in research. In June 2021, the Metastatic Breast Cancer Project was made available in Spanish (www.MBCprojectenEspanol.org) to increase cancer research accessibility to Spanish-speaking Hispanic and Latino/Latina/Latinx/Latine (H/L) people with cancer. All materials were translated/rewritten with iterative feedback from the community to ensure that materials were culturally and community appropriate. Efforts to reach members of the H/L community continue through the development of community alliances, increasing awareness of Count Me In research opportunities, and wider social media engagement. More specifically, these objectives have been achieved through partnerships with local community organizations, continuous collaboration with current Count Me In participants to develop H/L specific engagement materials, and growing Count Me In's H/L's reach through virtual engagement by leveraging social media platforms to share relevant cancer informational and educational posts. Additionally, in March 2022, the *Amplifying Black Voices Across Cancer* digital initiative ('#AmpBlackVoices', www.BlackCancerVoices.org) was launched in direct collaboration with Black cancer patients, loved ones/caregivers, and advocates. Supported by Count Me In, *Voices* aims to amplify and shed light on the lived experiences of Black patients, caregivers/loved ones, and advocates affected by cancer, including the disparities and inequities that exist within this space. Beyond the MBCproject in Spanish and the *Amplifying Black Voices Across Cancer* initiative, there are several other ongoing parallel efforts to reach other historically marginalized communities, including faith-based outreach, engagement with college campuses, physician-focused engagement, and LGBTQI+ initiatives.

#### The Metastatic Breast Cancer Project website

The Metastatic Breast Cancer Project website (MBCproject.org) was developed after extensive collaboration with patients and advocates from the metastatic breast cancer community. The website enables metastatic breast cancer patients across the United States and Canada to learn about the project, register to participate remotely, sign an electronic consent form, and provide information about themselves and their experience via surveys. Over time, the website changed in response to patient feedback. The original website was designed and developed by external collaborators at Playground, Inc. in 2015. Backend data was stored and managed through secure systems including a DatStat study management instance. In 2020, the website was migrated to Count Me In's study platform, Pepper, and backend data management transitioned to Count Me In's custom, secure, internal study management system, the Broad Institute's Data Donation Platform's Data Study Manager. On the new study platform, patients can set up a secure account using their email address that enables them to track their study activities via a dashboard.

#### Patient registration

After visiting MBCproject.org, patients clicked "Count Me In" and were prompted to enter their first and last name, confirm diagnosis of metastatic breast cancer, and enter email address and password to create a study account. Next, patients were directed to complete an 18-question intake survey with acknowledgement that their responses were stored in a secure database, an agreement to be recontacted, and an understanding that they could withdraw from the study at any time, which stops collection of any new data from samples, medical records, or surveys. Information collected prior to consent can be deleted upon withdrawal from the study.

#### Patient consent

All patients who register for the MBCproject are given the option to consent for the study through an electronic consent form. The process for consenting patients has changed over time. At the launch of the study through to the study's migration to the new website platform in May 2020, the study team sent consents out to new project registrants weekly. On the new platform, the consent process is continuous and patients are brought directly to an electronic consent form after completing the intake survey. As of May 2020, participants were provided with two separate consent forms – one to provide consent to allow the study team to collect a saliva sample, copies of medical records, and portions of archived tumor tissue and another to consent to allowing the study team to collect a saliva sample, copies of medical records, and a blood sample. As of May 2020, patients are able to opt-in to sharing archived tissue and/or blood collection on a single consent form. Participants signed the consent form(s) with their full name and date of birth. The blood-only consent also asked participants about the names and duration (start and stop dates) of current and past therapies. Consent also allows for sequencing analysis, and the sharing of de-identified clinical, genomic, and patient-reported data on various public sharing platforms including cBioPortal, dbGaP, and GDC.

#### Patient-reported data (PRD) - follow-up survey-based treatment duration analysis

Treatments (prior and current) received by patients (N = 1511 of 1604 patients that completed the follow-up survey; 93 patients were removed due to missing start/stop dates) from the follow-up survey were cleaned using a fuzzy match algorithm and matched to NCI's drug name database. All drug names were converted into standard naming convention. Each treatment name was assigned a drug category as described in Suppl. Table S4. Patients that exhibited extraordinary responses to a particular drug were identified by selecting for past treatments which patients had received for two years or more after metastatic disease diagnosis.

#### Blood and saliva sample acquisition

Enrolled patients who provided informed consent and completed the release form to indicate their mailing address were mailed saliva collection kits if a valid address in the United States or Canada was provided. Prior to shipment, saliva kits were labeled with a unique two dimensional barcode. Each kit had a business reply label affixed to the back addressed to the Broad Institute's Genomics Platform Sample's Lab in Cambridge, MA. Prior to shipping out each kit, it was scanned to associate it with a participant in the study using the Data Study Manager (DSM), an internal study management platform. Participants were instructed to provide at least 2 mL of saliva into the collection tube (DNA Genotek OGR 600) following the instructions included in each kit. Saliva kits received back at the Broad Institute were scanned using their unique barcode and stored at room temperature until moving them forward for whole exome sequencing (WES). Enrolled participants who provided informed consent to contribute a blood sample, which is optional, and completed the release form to indicate their mailing address were mailed blood collection kits if a valid address in the United States or Canada was provided.

#### Tissue sample acquisition

For enrolled participants who opted to share a portion of their archived tumor tissue, medical records were reviewed in order to record the type of procedure(s) (e.g. mastectomy, core biopsy, lumpectomy), date of procedure, size of procedure's sample, and facility that performed the procedure. Each participant's procedure history was reviewed in order to determine which procedures were requestable. Unless the most recent procedure's sample measured was greater than 1 cm, the most recent samples were not requested in order to avoid exhausting tissue samples that participants may need for future clinical care.

After determining which procedures were requestable, the study staff called the pathology departments associated with each tissue sample in order to confirm the fax number for the requests. Forms were faxed to each pathology department requesting one Hematoxylin and Eosin stain (H&E) slide as well as either 5-μm unstained slides (a minimum of 8 and a maximum of 25 unstained slides) or one Formalin-Fixed Paraffin-Embedded (FFPE) tissue block. Requests explicitly stated sample should not be exhausted in order to fulfill the request.

Tissue samples were received at the Broad Institute (Cambridge, MA) by mail. Tissue samples that were received as FFPE tissue blocks were labeled with a unique numerical identifier associated with the participant it belonged to and sent to the Dana-Farber/Harvard Cancer Center Specialized Histopathology Services - Longwood (SHL) Core, which cut three 30-μm scrolls per block, as well as 3 unstained slides and one H&E for pathology review. SHL only cut the requested tissue after doing a thorough review to determine that it would not exhaust the sample to do so. All scrolls were labeled with unique barcode identifiers. Tissue samples received as unstained slides were logged and labeled with unique barcode identifiers.

Scrolls and unstained slides were submitted to the Broad Institute's Genomics Platform for whole exome sequencing and RNA-sequencing. Germline samples (derived from saliva of blood buffy coat) were submitted in conjunction with tumor samples to make a matched tumor/normal pair.

#### Medical record acquisition

Participants provided the names of their treating institutions and physicians on the medical release form, which was then used by study staff to call each of the hospitals/institutions listed and confirm the fax number of the medical records department. A detailed request was electronically faxed to each facility, asking for medical records that include all clinic notes, treatment data (including radiation and chemotherapy), pathology reports, operative reports, referrals, MD to MD exchanges, and genetic testing reports related to the patient's metastatic breast cancer diagnosis from the data of initial diagnosis with primary breast cancer to present day (the date of the faxed request). Medical records not received for several months were re-requested. Medical records were received in multiple modalities, including by fax, secure electronic message, or physician mail. All medical records were scanned (if required) and saved to a secure server accessible only to specific study staff trained to handle identifiable patient data. All physical medical records were securely shredded and destroyed after being saved to the secure digital server.

#### Medical record data (MRD) - medical record abstraction

For each medical record (MR) acquired by the study team, over 100 clinical fields were abstracted following an established clinical data dictionary model (Suppl. File 2) by the study staff abstraction team. The abstracted fields contain information about clinical and pathological features including pre diagnostic, diagnostic, staging, surgery and treatments details/dates, information on other cancers and genomic tests details. Some of the key fields abstracted from the MRs are described below:

1. **Metastatic disease setting (METASTATIC_DISEASE_PRESENT/NO_METASTATIC_DISEASE_PRESENT).** Metastatic setting of a sequenced tumor sample is defined based on whether the sample was taken before (NO_METASTATIC_DISEASE_PRESENT) or at/after (METASTATIC_DISEASE_PRESENT) the date of metastatic diagnosis. Blood (ctDNA) samples from patients for which a medical record could not be obtained are classified as METASTATIC DISEASE PRESENT since they were presumed to be taken after metastatic diagnosis.
2. **Treatment naive (Yes/No).** A biopsy is classified as treatment naive if the sequenced biopsy was collected before the start of any treatment for breast cancer.
3. **Primary date of diagnosis.** The date of primary breast cancer diagnosis is derived from the date of the diagnostic surgical procedure, which is most often a biopsy but can in rare occasions be a lumpectomy/excision. If the date of primary diagnostic biopsy is not available, date of diagnostic imaging was used instead. All dates are noted as the number of days relative to the date of primary breast cancer diagnosis.
4. **Metastatic date of diagnosis.** The date of metastatic breast cancer diagnosis is derived from the date of the metastatic surgical procedure, which is most often a biopsy. If the date of metastatic diagnostic biopsy is not available, the date of metastatic diagnostic imaging was used instead. All dates are noted as the number of days relative to the date of primary breast cancer diagnosis.
5. **Stage.** Staging is taken explicitly as reported in the MRs. If this is not available, it is annotated as 'NOT FOUND IN RECORD'
    a. Diagnostic stage (DX_stage). Diagnostic staging is taken explicitly if reported in the MR around the patient's primary diagnosis based on biopsy and imaging .
    b. Surgery Stage (SX_stage). Surgery stage information is taken explicitly if reported in the MR around the patient's first curative surgery based on metastatic biopsy and imaging.

6. **Receptor status (ER, PR, HER2OVERALL).** Receptor status is taken explicitly as reported in the MRs. If this is not available, it is annotated as 'NOT FOUND IN RECORD'
    a. Diagnostic receptor status. Diagnostic receptor status is taken explicitly if reported in the MR around the patient's primary diagnosis based on the diagnosis biopsy
    b. Biopsy receptor status. Biopsy level receptor status that is taken explicitly from the pathology reports that is sent with the tumor biopsies.
7. **Treatment response (Response, Mixed, Stable, Progression, NED, Other, Not Found in Record).** Treatment response is defined as a patient's response to a specific treatment within the first 6 months. It is based on the response reported in the provider notes from the MRs.
8. **Discontinuation Reason (PROGRESSION, ADVERSE EFFECTS, SURGERY, FINISHED COURSE, NED, ON TX AT LAST DATE OF ENCOUNTER).** Reason why a drug used during a treatment was discontinued.
9. **Treatment mode.** Treatment modes were classified as pre-diagnosis, neoadjuvant, adjuvant, metastatic, and local recurrence.
    a. Pre-diagnosis. Any treatment received for a prior invasive or *in situ* breast cancer that is presumably unrelated to the invasive breast cancer that led to MBC.
    b. Neoadjuvant. Any treatment received prior to the first curative surgery.
    c. Adjuvant. Any treatment received after the first curative surgery.
    d. Metastatic. Any treatment received after being diagnosed with metastatic disease.
    e. Local Recurrence. Any treatment received when diagnosed with local recurrence.

#### Additional information on the attrition of genomic and clinical data in collection and processing

In this section we give additional information on the attrition of genomic and clinical data given in the result section "*Generating a clinco-genomic dataset for the MBCproject"* of the main text and in Table 1, Table 2, and Suppl. Fig. S3.

As of March 31, 2020, 3246 patients had registered and consented to participate in the study. 2985 of these patients signed the online medical release form. 1604 out of the 3246 patients filled out the follow-up survey, which could have been because the survey was sent out approximately 3 years after project enrollment began and patients were no longer engaged with the study, because patients passed away, or because of some other reason. For the 2985 patients that signed the medical release form, 3054 medical records were requested for 2196 patients. Of the 3054 medical records requested, 1700 were received and 1354 were not received for the following reasons: records were not available at the institutions listed, records were unable to be released for authorization reasons, or no response was ever received from the institution.

Of the 852 tissue samples requested, 608 samples were received and 244 have not yet been received. Of the 608 tissue samples, 437 were sent for whole exome sequencing (WES) and 171 were not for the following reasons: the block did not have sufficient material for cutting scrolls, the sample was requested back from the pathology department before it could be sequenced, the pathology department sent the incorrect tissue, or tissue was still being processed.

Of the 437 tissue samples sent for WES, 275 were included in the final WES cohort because they had enough DNA for WES and passed the tumor purity cutoff of 10%, 46 samples did not contain sufficient DNA for WES and 116 samples did not pass the tumor purity cutoff of 10%. Of the 228 samples sent for RNA sequencing (RNA-seq), 200 samples passed the RNA-seq QC metrics and were included in the final RNA-seq cohort, while 28 samples did not pass the QC metrics and were excluded.

Of the 1463 patients who were sent blood kits, 1112 returned a blood kit and 351 did not return them. Of these 1112 returned blood samples, cfDNA from 995 samples was submitted for ULP-WGS while 117 samples remained in the queue for ULP-WGS. Of the 995 cfDNA samples submitted for ULP-WGS, 144 cfDNA samples were submitted for WES and 851 were not submitted for WES because tumor fraction was determined to be <5%. Of the 144 cfDNA samples submitted for WES, 104 cfDNA samples were submitted for WES, passed the tumor purity cutoff of 10%, and were included in the final WES cohort, while 40 cfDNA samples did not pass the tumor purity cutoff of 10%.

Overall, the MBCproject clinico-genomic dataset consists of WES for 275 FFPE tumor tissue samples from 204 patients (with matching germline WES samples), WES for 104 ctDNA samples from 104 patients (with matching germline WES samples), RNA-seq for 200 FFPE tissue samples from 141 patients, and 377 germline samples from 377 patients.

The 379 WES tumors (275 tumor tissue samples and 104 ctDNA samples) from 301 patients and 157 RNA-Seq tumors ( (those with both WES and RNA-seq data) from 120 patients were submitted to cBioPortal (https://www.cbioportal.org/study/summary?id=brca_mbcproject_2022) along with associated patient-reported and clinical data. A total of 237 WES tumors with associated germline WES from 180 patients and 203 RNA-Seq tumors from 140 patients were submitted to the National Cancer Institute's Genomic Data Commons (GDC) (https://portal.gdc.cancer.gov/projects/CMI-MBC). A total of 237 WES tumors with associated germline WES from 180 patients and 228 tumors from 154 patients were submitted to dbGaP (Study Accession phs001709). As of May 1st 2023, updating the Genomic Data Commons and dbGaP data repositories with the genomic data for all the tumor samples used in this study (379 WES tumors with associated germline WES from 301 patients, 200 RNA-Seq tumors from 141 patients, and germline WES for a total of 377 patients) is in progress.

#### Supplemental genomic and sample processing methods

##### *Ultra Low Pass Whole Genome Sequence data processing and quality control for blood biopsies*

Each of the obtained cfDNA samples are subjected to ultra low coverage whole genome sequencing to estimate the tumor content (using ichorCNA) for each sample (Adalsteinsson et al. 2017). Samples with >= 5% estimated tumor fraction were then submitted for whole exome sequencing.

##### *Somatic alterations assessment*

A custom made cancer genomics analysis pipeline was used to identify somatic alterations using the Terra platform (https://app.terra.bio/). We have utilized the CGA WES Characterization pipeline developed at the Broad Institute to call, filter and annotate somatic mutations and copy number variation (available in the Terra platform public workspace *broad-fc-getzlab-workflows/CGA_WES_Characterization_OpenAccess*) (documentation available in https://docs.google.com/document/d/1VO2kX_fgfUd0x3mBS9NjLUWGZu794WbTepBel3cBg08). The pipeline employs the following tools: MuTect (Cibulskis et al. 2013), ContEst (Cibulskis et al. 2011), Strelka (Saunders et al. 2012), Orientation Bias Filter (Costello et al. 2013), DeTiN (Taylor-Weiner et al. 2018), AllelicCapSeg (Landau et al. 2013), MAFPoNFilter (Lawrence et al. 2014), BLAT realignment filter, ABSOLUTE (Carter et al. 2012), GATK (McKenna et al. 2010), GATK CNV (Van der Auwera and O'Connor 2020), Picard Tools *CrosscheckFingerprints* and *CollectMultipleMetrics* (https://broadinstitute.github.io/picard/), Variant Effect Predictor (Shamsani et al. 2019), and Oncotator (Ramos et al. 2015). To annotate known oncogenic mutations, the OncoKB (Chakravarty et al. 2017) annotator was used (https://github.com/oncokb/oncokb-annotator).

For the comutation plots, variants classified as silent or intron are not included. Variants included in the comutation plot that are not in a hotspot region and not assigned an oncogenic effect by OncoKB are classified as either missense (if classified as a "Missense_Mutation"), putative loss of function (if classified as either of these mutations: "Splice_Site" ,"Frame_Shift_Del", "Frame_Shift_Ins", "Nonsense_Mutation", "Nonstop_Mutation", "In_Frame_Del", "Start_Codon_SNP", "In_Frame_Ins", "Start_Codon_Ins", "Stop_Codon_Del", or "Stop_Codon_Ins"), or other mutation (otherwise). The multi-hit classification is used if there are multiple alterations in the gene (excluding silent or intron variants).

MutSig2CV (Lawrence et al. 2013) was used to infer significantly recurrent mutated genes for specific sub cohorts. MutSig2CV uses patient- and gene-specific mutation rates to estimate a background model of predicted mutation incidence across the genome, and factors in biological covariates such as replication timing and gene-expression level on a gene-by-gene basis to account for the increased mutational rate of certain classes of genes. Genes were called significantly mutated if $q<0.10$. Mutations in cancer genes that were significantly mutated were manually checked to verify if they appeared to be artifacts and genes with a large number of mutations that appeared to be artifacts were removed.

Genes with mutations that appeared to be artifacts had the same mutation across different tumors and patients in genes without or with few known oncogenic mutations, were exclusive to ctDNA samples, and/or had relatively few mutations (8 or less). These genes were *USP8*, *CDH17*, *RAD9A*, *DROSHA*, *RGPD3*, *EZR*, and *PSIP1*. Other genes with mutations that appeared to be artifacts were *ACTB*, *HIST3H2BB*, *FANCD2*, and *POLD1*; it was not completely clear if the mutations in these genes were artifacts, but these genes were removed out of caution. The mutations in the genes *STAG2* and *POLA1* were not included because they were removed by the Mutation Validator tool (Ellrott et al. 2018).

The original number of significantly mutated genes ($q<0.10$) in the metastatic tumors (N=249) and the treatment naive tumors (N=157) were 117 and 15 genes, respectively. When restricted to cancer genes the number of genes were 27 and 15 genes, respectively. When restricted to cancer genes and removing genes with artifact mutations the number of genes were 17 and 11 genes, respectively.

##### *DNA and RNA extraction for tumors and whole blood*

DNA extraction was performed as described previously (Fisher et al. 2011). For whole blood, DNA was extracted using magnetic bead–based chemistry using the Chemagic DNA Blood Kit-96 or the QIAsymphony DSP DNA midi kit in conjunction with the Chemagic MSM I instrument (Perkin Elmer) or the QIAsymphony SP instrument (Qiagen). Following red blood cell lysis, magnetic beads bind to the DNA and were removed from solution using electromagnetized rods. Several wash steps followed to eliminate cell debris and protein residue from DNA bound to the magnetic beads. DNA was then eluted in TE buffer. For FFPE tumor tissues, DNA and RNA were extracted using Qiagen's AllPrep DNA/RNA FFPE Kit. All DNA was quantified using Picogreen.

##### *DNA extraction for saliva*

DNA was extracted using magnetic bead–based chemistry using the Chemagic DNA Blood Kit-96 in conjunction with the Chemagic MSM I instrument (Perkin Elmer). Saliva samples were incubated at 50°C for 2 hours. The saliva was then transferred to a deep well plate placed on the Chemagic MSM I. The following steps were automated on the MSM I. M-PVA Magnetic Beads were added to the saliva. Lysis buffer was added to the solution and mixed. The bead-bound DNA was then removed from solution via a 96-rod magnetic head and washed in three Ethanol-based wash buffers. The beads were then washed in a final water wash buffer. Finally, the beads were dipped in an elution buffer to resuspend the DNA sample in solution. The beads were then removed from solution, leaving purified DNA eluate. DNA samples were quantified using a fluorescence-based PicoGreen assay.

##### *DNA extraction for cfDNA from whole blood*

Blood tubes were centrifuged at 1900 g for 10 minutes and plasma was transferred to a second tube before further centrifugation at 15000 g for 10 minutes. Supernatant plasma was stored at -80°C until cfDNA extraction. cfDNA was extracted using the QIAsymphony DSP Circulating DNA Kit according to the manufacturer's instructions, with 6.3 mL of plasma as input and with a 60 µL DNA elution (Qiagen, 2017).

##### *Library construction (exomes)*

**Tumors, whole blood, and saliva.** DNA libraries for massively parallel sequencing were generated as described previously (Fisher et al. 2011) with the following modifications: the initial genomic DNA input into the shearing step was reduced from 3 µg to 10–100 ng in 50 µL of solution. For adapter ligation, Illumina paired-end adapters were replaced with palindromic forked adapters (purchased from Integrated DNA Technologies) with unique dual indexed 8 base index molecular barcode sequences included in the adapter sequence to facilitate downstream pooling. Kapa HyperPrep reagents in 96-reaction kit format were used for end repair/A-tailing, adapter ligation, and library enrichment PCR. In addition, during the post-enrichment solid-phase reversible immobilization (SPRI) bead cleanup, elution volume was reduced to 30 µL to maximize library concentration, and a vortexing step was added to maximize the amount of template eluted.

**cfDNA.** Initial DNA input was normalized to be within the range of 25-52.5 ng in 50 µL of TE buffer (10 mM Tris HCl 1 mM EDTA, pH 8.0) according to picogreen quantification. Library preparation was performed using a commercially available kit provided by KAPA Biosystems (KAPA HyperPrep Kit with Library Amplification product KK8504) and IDT's duplex UMI adapters. Unique 8-base dual index sequences embedded within the p5 and p7 primers (purchased from IDT) were added during PCR. Enzymatic clean-ups were performed using Beckman Coultier AMPure XP beads with elution volumes reduced to 30 µL to maximize library concentration. Library quantification was performed using the Invitrogen Quant-It broad range dsDNA quantification assay kit (Thermo Scientific Catalog: Q33130) with a 1:200 PicoGreen dilution. Following quantification, each library is normalized to a concentration of 25 ng/µL, using Tris-HCl, 10mM, pH 8.0.

##### *Solution-phase hybrid selection (exomes)*

After library construction, hybridization and capture were performed using the relevant components of Illumina's Nextera Rapid Capture Exome Kit or TruSeq Rapid Exome Kit following the manufacturer's suggested protocol, with the following exceptions: first, all libraries within a library construction plate were pooled prior to hybridization. Second, the Midi plate from Illumina's Exome Kit was replaced with a skirted PCR plate to facilitate automation. All hybridization and capture steps were automated on the Agilent Bravo liquid handling system.

##### *Preparation of libraries for cluster amplification and sequencing (exomes)*

After post-capture enrichment, library pools were then quantified using quantitative PCR (KAPA Biosystems) with probes specific to the ends of the adapters; this assay was automated using Agilent's Bravo liquid handling platform. Based on qPCR quantification, libraries were normalized to 2 nM, then denatured using 0.1 or 0.2 N NaOH on the Hamilton Starlet.

##### *Cluster amplification and sequencing (exomes)*

**Tumors, whole blood, and saliva.** Cluster amplification of denatured templates was performed according to the manufacturer's protocol (Illumina) using HiSeq 4000 or exclusion amplification cluster chemistry and HiSeq 4000 or HiSeq X flowcells. Flowcells were sequenced on v1 Sequencing-by-Synthesis chemistry for HiSeq 4000 flowcells or v2.5

Sequencing-by-Synthesis chemistry for HiSeq X flowcells. The flowcells were then analyzed using RTA v.1.18.64 or later. Each pool of whole exome libraries was run on paired 76 bp runs, reading the dual-indexed sequences to identify molecular indices and sequenced across the number of lanes needed to meet coverage for all libraries in the pool.

**cfDNA.** Cluster amplification of library pools was performed according to the manufacturer's protocol (Illumina) using exclusion amplification cluster chemistry and HiSeq X flowcells. Flowcells were sequenced on v2 Sequencing-by-Synthesis chemistry for HiSeq X flowcells. The flowcells are then analyzed using RTA v.2.7.3 or later. Each pool of libraries was run on paired 151 bp runs, reading the dual-indexed sequences to identify molecular indices and sequenced across the number of lanes needed to meet coverage for all libraries in the pool.

##### *cDNA library construction (transcriptomes)*

**Transcriptome capture (FFPE tissue).** Total RNA was assessed for quality using the Caliper LabChip GX2. The percentage of fragments with a size greater than 200nt (DV200) was calculated using software. An aliquot of 200ng of RNA was used as the input for first strand cDNA synthesis using Illumina's TruSeq RNA Access Library Prep Kit. Synthesis of the second strand of cDNA was followed by indexed adapter ligation. Subsequent PCR amplification enriched for adapted fragments. The amplified libraries were quantified using an automated PicoGreen assay.

200 ng of each cDNA library, not including controls, were combined into 4-plex pools. Capture probes that target the exome were added, and hybridized for recommended time. Following hybridization, streptavidin magnetic beads were used to capture the library-bound probes from the previous step. Two wash steps effectively remove any nonspecifically bound products. These same hybridization, capture and wash steps are repeated to assure high specificity. A second round of amplification enriches the captured libraries. After enrichment the libraries were quantified with qPCR using the KAPA Library Quantification Kit for Illumina Sequencing Platforms and then pooled equimolarly. The entire process is in 96-well format and all pipetting is done by either Agilent Bravo or Hamilton Starlet.

##### *Illumina sequencing (transcriptomes)*

Pooled libraries were normalized to 2 nM and denatured using 0.1 NaOH prior to sequencing. Flowcell cluster amplification and sequencing were performed according to the manufacturer's protocols using HiSeq 2000 or HiSeq 2500. Each run was a 76 bp paired-end with an eight-base index barcode read. Data was analyzed using the Broad Picard Pipeline which includes de-multiplexing and data aggregation.

##### *Figure and plot generation and editing*

Plots were generated using R (version 4.0.3) and the packages ggplot2 (Wickham 2016), ggpubr (U.S. Census Bureau 2021), and ComplexHeatmap (Gu 2022). Figures were edited using Microsoft PowerPoint and Inkscape.

Scalable, Fully Automated Process for Construction of Sequence-Ready Human Exome Targeted Capture Libraries.” *Genome Biology* 12 (1): R1.

Gu, Zuguang. 2022. “Complex Heatmap Visualization.” *iMeta* 1 (3). https://doi.org/10.1002/imt2.43.

Landau, Dan A., Scott L. Carter, Petar Stojanov, Aaron McKenna, Kristen Stevenson, Michael S. Lawrence, Carrie Sougnez, et al. 2013. “Evolution and Impact of Subclonal Mutations in Chronic Lymphocytic Leukemia.” *Cell* 152 (4): 714–26.

Lawrence, Michael S., Petar Stojanov, Craig H. Mermel, James T. Robinson, Levi A. Garraway, Todd R. Golub, Matthew Meyerson, Stacey B. Gabriel, Eric S. Lander, and Gad Getz. 2014. “Discovery and Saturation Analysis of Cancer Genes across 21 Tumour Types.” *Nature* 505 (7484): 495–501.

Lawrence, Michael S., Petar Stojanov, Paz Polak, Gregory V. Kryukov, Kristian Cibulskis, Andrey Sivachenko, Scott L. Carter, et al. 2013. “Mutational Heterogeneity in Cancer and the Search for New Cancer-Associated Genes.” *Nature* 499 (7457): 214–18.

McKenna, Aaron, Matthew Hanna, Eric Banks, Andrey Sivachenko, Kristian Cibulskis, Andrew Kernytsky, Kiran Garimella, et al. 2010. “The Genome Analysis Toolkit: A MapReduce Framework for Analyzing next-Generation DNA Sequencing Data.” *Genome Research* 20 (9): 1297–1303.

Ramos, Alex H., Lee Lichtenstein, Manaswi Gupta, Michael S. Lawrence, Trevor J. Pugh, Gordon Saksena, Matthew Meyerson, and Gad Getz. 2015. “Oncotator: Cancer Variant Annotation Tool.” *Human Mutation* 36 (4): E2423–29.

Saunders, Christopher T., Wendy S. W. Wong, Sajani Swamy, Jennifer Becq, Lisa J. Murray, and R. Keira Cheetham. 2012. “Strelka: Accurate Somatic Small-Variant Calling from Sequenced Tumor–normal Sample Pairs.” *Bioinformatics* 28 (14): 1811–17.

Shamsani, Jannah, Stephen H. Kazakoff, Irina M. Armean, Will McLaren, Michael T. Parsons, Bryony A. Thompson, Tracy A. O’Mara, Sarah E. Hunt, Nicola Waddell, and Amanda B. Spurdle. 2019. “A Plugin for the Ensembl Variant Effect Predictor That Uses MaxEntScan to Predict Variant Spliceogenicity.” *Bioinformatics* 35 (13): 2315–17.

Taylor-Weiner, Amaro, Chip Stewart, Thomas Giordano, Mendy Miller, Mara Rosenberg, Alyssa Macbeth, Niall Lennon, et al. 2018. “DeTiN: Overcoming Tumor-in-Normal Contamination.” *Nature Methods* 15 (7): 531–34.

U.S. Census Bureau. 2021. “Racial and Ethnic Diversity in the United States: 2010 Census and 2020 Census.” https://www.census.gov/library/visualizations/interactive/racial-and-ethnic-diversity-in-the-united-states-2010-and-2020-census.html.

Van der Auwera, Geraldine A., and Brian D. O’Connor. 2020. *Genomics in the Cloud: Using Docker, GATK, and WDL in Terra*. O’Reilly Media.

Wickham, Hadley. 2016. “Data Analysis.” In *ggplot2: Elegant Graphics for Data Analysis*, edited by Hadley Wickham, 189–201. Cham: Springer International Publishing.
