## Supplemental File 1 for "The Metastatic Breast Cancer Project: leveraging patient-partnered research to expand the clinical and genomic landscape of metastatic breast cancer and accelerate discoveries"

#### Forms and Surveys

---


| Forms | Page Number |
| --- | --- |
| Tissue Consent Form | 1 |
| Blood Consent Form | 13 |
| Release Form | 26 |
| **Surveys** | |
| Intake Survey (version 1) | 29 |
| Intake Survey (version 2) | 30 |
| Intake Survey (version 3) | 31 |
| Follow-Up Survey | 39 |
| **Request Templates** | |
| Medical Record Request Template | 42 |
| Pathology Report Request Template | 44 |
| Tissue Request Template | 46 |

415 Main Street
Cambridge, MA 02142
T 617-714-7000
www.joincountmein.org

#### The Metastatic Breast Cancer Project

#### RESEARCH CONSENT FORM – KEY POINTS

"The Metastatic Breast Cancer Project" is a patient-driven movement that empowers metastatic breast cancer patients to directly transform research and treatment of disease by sharing part of their stored tumor tissue and copies of their medical records. Because we are enrolling patients across the country regardless of where they are being treated, this study will allow many more patients to contribute to research than has previously been possible.

##### 1. What is the purpose of this study?

We want to understand metastatic breast cancer better so that we can develop more effective therapies. By partnering directly with patients, we are able to study many more aspects of cancer than would otherwise be possible.

##### 2. What will I have to do if I agree to participate in this study?

Participation requires little effort. You will answer some questions about your cancer and medical care and send a saliva sample to us in a pre-stamped package that we will provide. We will take care of obtaining copies of your medical records as well as small amounts of your stored tumor tissues from the hospitals or centers where you receive your medical care.

##### 3. Do I have to participate in this study?

No. Taking part in this study is voluntary. Even if you decide to participate, you can always change your mind and leave the study.

##### 4. Will I benefit from participating?

While taking part in this study may not improve your own health, the information we collect will aid in our research efforts to provide better cancer treatment and prevention options to future patients. We will provide updates about key research discoveries made possible by your participation on our website.

##### 5. What are the risks of taking part in this research?

There may be a risk that your information (which includes your genetic information and information from your medical records) could be seen by unauthorized individuals. However, we have procedures and security measures in place designed to minimize this risk and protect the confidentiality of your information

415 Main Street
Cambridge, MA 02142
T 617-714-7000
www.joincountmein.org

**6. Will it cost me anything to participate in this study?**
No.

**7. Who will use my samples and see my information?**
Your samples and health information will be available to researchers at the Broad Institute of MIT and Harvard, a not-for-profit biomedical research institute. After removing your name and other readily identifiable information, we will share results obtained from your participation with the greater research community as well as central data banks at the National Institutes of Health.

**8. Can I stop taking part in this research study?**
Yes, you can withdraw from this research study at any time, although any of your information that has already been entered into our system cannot be withdrawn. Your information would be removed from future studies.

**9. What if I have questions?**

415 Main Street
Cambridge, MA 02142
T 617-714-7000
www.joincountmein.org

**RESEARCH CONSENT FORM – KEY POINTS**

#### A. Introduction

You are being invited to participate in a research study that will collect and analyze samples and health information of patients with metastatic breast cancer, also called Stage IV or advanced breast cancer. This study will help doctors and researchers better understand why metastatic breast cancer occurs and develop ways to better treat and prevent it.

Cancers occur when the molecules that control normal cell growth (genes and proteins) are altered. Changes in the genes of tumor cells and normal tissues are called "alterations." Several alterations that occur in certain types of cancers have already been identified and have led to the development of new drugs that specifically target those alterations. However, the vast majority of tumors from patients have not been studied, which means there is a tremendous amount of information still left to be discovered. Our goal is to discover more alterations, and to better understand those that have been previously described. We think this could lead to the development of additional therapies and cures.

Genes are composed of DNA "letters," which contain the instructions that tell the cells in our bodies how to grow and work. We would like to use your DNA to look for alterations in cancer cell genes using a technology called "sequencing."

Gene sequencing is a way of reading the DNA to identify alterations in genes that may contribute to the behavior of cells. Some changes in genes occur only in cancer cells. Others occur in normal cells as well, in the genes that may have been passed from parent to child. This research study will examine both kinds of genes.

You are being asked to participate in the study because you have metastatic breast cancer. Other than providing a sample of saliva, participating in the study involves no additional tests or procedures.

This form explains why this research study is being done, what is involved in participating, the possible risks and benefits of the study, alternatives to participation, and your rights as a participant. The decision to participate is yours. We encourage you to ask questions about the study now or in the future.

#### B. Why is this research study being done?

We want to understand cancer better so that we can develop more effective therapies. By partnering directly with patients, we will be able to study many more aspects of cancer than has previously been possible. In addition, because we are enrolling patients across the country regardless of where they are being treated, this study will allow many more patients to directly contribute to research than might otherwise be feasible.

#### C. What other options are there?

Taking part in this research study is voluntary – you may choose not to participate. Your decision not to participate will not affect your medical care in any way or result in any penalty or loss of benefits.

#### D. What is involved in the research study?

Participation will require that you answer some questions about your cancer and medical care, send a saliva sample to us in a pre-stamped package that we will provide. With your consent, we will obtain a portion of your tumor tissues from hospitals or centers where you received your medical care.

We will analyze the genes in your cancer cells (obtained from biopsies or surgical specimens stored in a hospital pathology department) and your normal samples (obtained from the saliva sample). No additional procedures will be required. The results of this analysis will be used to try to develop better ways to treat and prevent cancers.

We will link the results of the gene tests on your cancer and saliva with medical information that has been generated during the course of your treatment. We are asking your permission to obtain a copy of your medical record from places where you have received care for your cancer.

In some cases, a research doctor may contact you to find out if you would be interested in participating in a different or future research study based on information that may have been found in your tissue or saliva samples.

415 Main Street
Cambridge, MA 02142
T 617-714-7000
www.joincountmein.org

To allow sharing of information with other researchers, the National Institutes of Health (NIH) and other organizations have developed central data (information) banks that analyze information and collect the results of certain types of genetic studies. These central banks will store your genetic and medical information and provide the information to qualified researchers to do more studies. We will also store your genetic and medical information at the Broad Institute of MIT and Harvard and share your information with other qualified researchers. Therefore, we are asking your permission to share your results with these special banks and other researchers, and have your information used for future research studies, including studies that have not yet been designed, studies involving diseases other than cancer, and/or studies that may be for commercial purposes (such as the development or approval of new drugs). Your information will be sent to central banks and other researchers only with a code number attached. Your name, social security number, and other information that could readily identify you will not be shared with central banks or other researchers. We will never sell your readily identifiable information to anyone under any circumstances.

#### E. How long will I be in this research study?

You will be asked to give a sample of saliva at the beginning of the study. We will keep your tissue and saliva samples and medical records indefinitely until this study is finished unless you inform us that you no longer wish to participate. You may do this at any time. More information about how to stop being in the study is below in paragraph I.

Once the study is finished, any left over tissue samples that we have will be returned to the pathology department at the hospital or other place where you received treatment and any left over saliva samples and your medical records will be destroyed.

#### F. What kind of information could be found in this study and will I be able to see it?

The gene tests in this study are being done to add to our knowledge of how genes and other factors affect cancer. This information will be kept confidential and while you will not receive information about your personal results obtained from studying your tissue and saliva, we will provide general results and major discoveries to all participants. We will do this by regularly updating the website that you used to enroll in this study. Furthermore, we will publish important discoveries found through these studies in the scientific literature so

415 Main Street
Cambridge, MA 02142
T 617-714-7000
www.joincountmein.org

that the entire research community can work together to better understand cancer. Your individual data will not be published in a way in which you could be readily identified. Abstracts, which are plain language summaries of the published reports, will be available to you and the general public.

#### G. What are the risks or discomforts of the research study?

There is a small risk that by participating in this study, the gene test results, including the identification of genetic changes in you or your cancer, could be seen by unauthorized individuals. We have tried to minimize this risk by carefully limiting access to the computers that would house your information to the staff of this research study.

There is a small but real risk that if your samples are used for this research study, they might not be available for clinical care in the future. However, we have attempted to minimize this risk in the following way: the pathologists in the department of pathology where your specimens are kept will not release your specimen unless they believe that the material remaining after the research test is performed is sufficient for any future clinical needs.

#### H. What are the benefits of the research study?

Taking part in this research study may not directly benefit you. By joining this study, you will help us and other researchers understand how to use gene tests to improve the care of patients with cancer in the future. We will provide study participants updates on our project website about key research discoveries made possible by your participation

#### I. Can I stop being in the research study and what are my rights?

You can stop being in the research study at any time. We will not be able to withdraw all the information that already has been used for research. If you tell us that you want to stop being in the study, we will return any remaining tumor samples from where we obtained them, and destroy any saliva samples or DNA samples we have. We will not perform any additional tests on the samples. Additionally, we will not collect any additional medical records and we will destroy the medical records we already have.

415 Main Street
Cambridge, MA 02142
T 617-714-7000
www.joincountmein.org

However, we will keep the results from the tests we did before you stopped being in the study. We will also keep the information we learned from reviewing your medical records before you stopped being in the study, We will not be able to take back the information that already has been used or shared with other researchers, central data banks, or that has been used to carry out related activities such as oversight, or that is needed to ensure quality of the study.

To withdraw your permission, you must do so in writing by contacting the researcher listed below in the section: "Whom do I contact if I have questions about the research study?" If you choose to not participate, or if you are not eligible to participate, or if you withdraw from this research study, this will not affect your present or future care and will not cause any penalty or loss of benefits to which you are otherwise entitled.

##### J. Will I be paid to take part in this research study?

There is no financial compensation for participation in this study.

##### K. What are the costs?

There are no costs to you to participate in this study.

##### L. What happens if I am injured or sick because I took part in this research study?

There is virtually no chance that you will become injured or sick by taking part in this study. Please contact us if you think you have been injured as a result of participating in this study.

##### M. What about confidentiality?

We will take rigorous measures to protect the confidentiality and security of all your information, but we are unable to guarantee complete confidentiality. When we receive your tissues and saliva samples, your name, social security number, and other information that could be used to readily identify you will be removed and replaced by a code. If we send your samples to our collaborators for gene testing, the samples will be identified using only this code. The medical records that we receive will be reviewed by our research team to confirm that you are eligible for the study and to obtain information about your medical condition and treatment.

We will store all of your identifiable information related to the study (including your medical records) in locked file cabinets and in password-protected computer files at the Broad Institute and we will limit access to such files. We may share your identifiable information or coded information, as necessary, with regulatory or oversight authorities (such as the Office for Human Research Protections), ethics committees reviewing the conduct of the study, or as otherwise required by law.

When we send the results of the gene tests and yourmedical information to central data banks or other researchers, they will not contain your name, social security number, or other information that could be used to readily identify you.

The results of this research study or future research studies using the information from this study may be published in research papers or included in presentations that will become part of the scientific literature. You will not be identified in publications or presentations.

#### N. Whom do I contact if I have questions about the research study?

- Nikhil Wagle, MD (Principle Investigator of the Study)
- Corrie Painter, PhD

For questions about your rights as a patient, please contact a representative of the Office for Human Research Studies at (617) 632-3029. This can include questions about your participation in the study, concerns about the study, a research related injury, or if you feel/felt under pressure to enroll in this research study or to continue to participate in this research study. Please keep a copy of this document in case you want to read it again.

#### O. AUTHORIZATION TO USE YOUR HEALTH INFORMATION FOR RESEARCH

**PURPOSES**

Because information about you and your health is personal and private, it generally cannot be used in this research study without your written authorization. Federal law requires that your health care providers and healthcare institutions (hospitals, clinics, doctor's offices) protect the privacy of information that identifies you and relates to your past, present, and future physical and mental health conditions.

If you sign this form, it will provide your health care providers and healthcare institutions the authorization to disclose your protected health information to the Broad Institute for use in this research study. The form is intended to inform you about how your health information will be used or disclosed in the study. Your information will only be used in accordance with this authorization form and the informed consent form and as required or allowed by law. Please read it carefully before signing it.

1. **What personal information about me will be used or shared with others during this research?**
    - Your medical records
    - Your tissue samples relevant to this research study and related records
    - New health information created from study-related tests and/or questionnaires
2. **Why will protected information about me be used or shared with others?**

    The main reasons include the following:

    - To conduct and oversee the research described earlier in this form;
    - To ensure the research meets legal, institutional, and accreditation requirements;
    - To conduct public health activities (including reporting of adverse events or situations where you or others may be at risk of harm)
3. **Who will use or share protected health information about me?**

    The Broad Institute and its researchers and affiliated research staff will use and/or share your personal health information in connection with this research study.
4. **With whom outside of the Broad Institute may my personal health information be shared?**

415 Main Street
Cambridge, MA 02142
T 617-714-7000
www.joincountmein.org

While all reasonable efforts will be made to protect the confidentiality of your protected health information, it may also be shared with the following entities:

- Federal and state agencies (for example, the Department of Health and Human Services, the Food and Drug Administration, the National Institutes of Health, and/or the Office for Human Research Protections), or other domestic or foreign government bodies if required by law and/or necessary for oversight purposes. A qualified representative of the FDA and the National Cancer Institute may review your medical records.
- Outside individuals or entities that have a need to access this information to perform functions relating to the conduct of this research such as data storage companies.

Some who may receive your personal health information may not have to satisfy the privacy rules and requirements. They, in fact, may share your information with others without your permission.

5. **For how long will protected health information about me be used or shared with others?**

   There is no scheduled date at which your protected health information that is being used or shared for this research will be destroyed, because research is an ongoing process.

6. **Statement of privacy rights:**
   - You have the right to withdraw your permission for the doctors and researchers to use or share your protected health information. We will not be able to withdraw all the information that already has been used or shared with others to carry out related activities such as oversight, or that is needed to ensure quality of the study. To withdraw your permission, you must do so in writing by contacting the researcher listed above in the section: “Whom do I contact if I have questions about the research study?”
   - You have the right to request access to your personal health information that is used or shared during this research and that is related to your treatment or payment for your treatment. To request this information, please contact your doctor who will request this information from the study directors.

415 Main Street
Cambridge, MA 02142
T 617-714-7000
www.joincountmein.org

#### P. Participation Information

If you decide to sign this consent form, we will ask you for information about contacting your physicians and the hospitals that you were treated at for your cancer. We will not disclose details about the results of your participation in this study with any of the individuals that we contact, but rather ask them to provide us with your medical history and your tissue samples.

#### Q. Documentation of Consent

This is what I agree to:

- You can request my stored tissue samples from my physicians and the hospitals and other places where I received my care, perform (or collaborate with others to perform) gene tests on the samples, and store the samples until this research study is complete.
- You can perform (or collaborate with others to perform) gene tests on the saliva sample that I will send you and store the sample until this research study is complete.
- You can request my medical records from my physicians and the hospitals and other places where I received and/or continue to receive my treatment and link results of the gene tests you perform on my tissue and saliva samples with my medical information from my medical records.
- You can use the results of the gene tests and my medical information for future research studies, including studies that have not yet been designed, studies for diseases other than cancer, and/or studies that may be for commercial purposes.
- You can share the results of the gene tests and my medical information with central data banks (e.g., the NIH) and with other qualified researchers in a manner that does not include my name, social security number, or any other information that could be used to readily identify me, to be used by other qualified researchers to perform future research studies, including studies that have not yet been designed, studies for diseases other than cancer, and studies that may be for commercial purposes.

My full name below indicates:

- I have had enough time to read the consent and think about agreeing to participate in this study;

- I have had all of my questions answered to my satisfaction;
- I am willing to participate in this research study;
- I have been told that my participation is voluntary and if I decide not to participate it will have no impact on my medical care;
- I have been told that if I decide to participate now, I can decide to stop being in the study at any time.
- I acknowledge that a copy of the signed consent form will be sent to my email address.

________________________
Full Name

________________________
Date

________________________
Email

________________________
Date of Birth

415 Main Street
Cambridge, MA 02142
T 617-714-7000
www.joincountmein.org

**The Metastatic Breast Cancer Project**

**RESEARCH CONSENT FORM (BLOOD DRAW) – KEY POINTS**

“The Metastatic Breast Cancer Project” is a patient-driven movement that empowers metastatic breast cancer patients to directly transform research and treatment of disease by sharing clinical information and tissue/blood samples with researchers in order to accelerate the pace of discovery. Because we are enrolling patients across the country regardless of where they are being treated, this study will allow many more patients to contribute to research than has previously been possible.

**1. What is the purpose of this study?**

We want to understand metastatic breast cancer better so that we can develop more effective therapies. By partnering directly with patients, we are able to study many more aspects of cancer than would otherwise be possible.

**2. What will I have to do if I agree to participate in this study?**

Participation requires little effort. We will ask you to have a sample of blood (1 tube or 2 teaspoons) drawn at your physician’s office, local clinic, or nearby lab facility – we will provide detailed instructions on how to do this. We’ll then ask you to send the sample to us in a pre-stamped package that we will provide. We will take care of obtaining copies of your medical records from the hospitals or centers where you receive your medical care. Participating in the study involves no tests or procedures beyond those required for your care.

**3. Do I have to participate in this study?**

No. Taking part in this study is voluntary. Even if you decide to participate, you can always change your mind and leave the study.

**4. Will I benefit from participating?**

While taking part in this study may not improve your own health, the information we collect will aid in our research efforts to provide better cancer treatment and prevention options to future patients. We will provide updates about key research discoveries made possible by your participation on our website.

**5. What are the risks of taking part in this research?**

There are small risks associated with obtaining the additional tube of blood. You may experience slight pain and swelling at the site of the blood draw. These complications are

415 Main Street
Cambridge, MA 02142
T 617-714-7000
www.joincountmein.org

rare and should resolve within a few days. If they do not, you should contact your doctor. There may be a risk that your information (which includes your genetic information and information from your medical records) could be seen by unauthorized individuals. However, we have procedures and security measures in place designed to minimize this risk and protect the confidentiality of your information.

**6. Will it cost me anything to participate in this study?**
No.

**7. Who will use my samples and see my information?**
Your samples and health information will be available to researchers at the Broad Institute of MIT and Harvard, a not-for-profit biomedical research institute. After removing your name and other readily identifiable information, we will share results obtained from your participation with the greater research community as well as central data banks at the National Institutes of Health.

**8. Can I stop taking part in this research study?**
Yes, you can withdraw from this research study at any time, although any of your information that has already been entered into our system cannot be withdrawn. Your information would be removed from future studies.

**9. What if I have questions?**

415 Main Street
Cambridge, MA 02142
T 617-714-7000
www.joincountmein.org

**RESEARCH CONSENT FORM – KEY POINTS**

#### A. Introduction

You are being invited to participate in a research study that will collect and analyze samples and health information of patients with metastatic breast cancer, also called Stage IV or advanced breast cancer. This study will help doctors and researchers better understand why metastatic breast cancer occurs and develop ways to better treat and prevent it.

Cancers occur when the molecules that control normal cell growth (genes and proteins) are altered. Changes in the genes of tumor cells and normal tissues are called "alterations." Several alterations that occur in certain types of cancers have already been identified and have led to the development of new drugs that specifically target those alterations. However, the vast majority of tumors from patients have not been studied, which means there is a tremendous amount of information still left to be discovered. Our goal is to discover more alterations, and to better understand those that have been previously described. We think this could lead to the development of additional therapies and cures.

Genes are composed of DNA "letters," which contain the instructions that tell the cells in our bodies how to grow and work. We would like to use your DNA to look for alterations in cancer cell genes using a technology called "sequencing."

Gene sequencing is a way of reading the DNA to identify alterations in genes that may contribute to the behavior of cells. Some changes in genes occur only in cancer cells. Others occur in normal cells as well, in the genes that may have been passed from parent to child. This research study will examine both kinds of genes.

You are being asked to participate in the study because you have metastatic breast cancer. Other than providing a sample of blood (1 tube or 2 teaspoons), participating in the study involves no additional tests or procedures.

This form explains why this research study is being done, what is involved in participating, the possible risks and benefits of the study, alternatives to participation, and your rights as a participant. The decision to participate is yours. We encourage you to ask questions about the study now or in the future.

415 Main Street
Cambridge, MA 02142
T 617-714-7000
www.joincountmein.org

**B. Why is this research study being done?**

We want to understand cancer better so that we can develop more effective therapies. By partnering directly with patients, we will be able to study many more aspects of cancer than has previously been possible. In addition, because we are enrolling patients across the country regardless of where they are being treated, this study will allow many more patients to directly contribute to research than might otherwise be feasible.

**C. What other options are there?**

Taking part in this research study is voluntary – you may choose not to participate. Your decision not to participate will not affect your medical care in any way or result in any penalty or loss of benefits.

**D. What is involved in the research study?**

Participation will require that you answer some questions about your cancer and medical care. We will ask you to have a sample of blood (1 tube or 2 teaspoons) drawn at your physician's office, local clinic, or nearby lab facility – we will provide detailed instructions on how to do this. We'll then ask you to send the sample to us in a pre-stamped package that we will provide. With your consent, we may also obtain copies of your medical records from hospitals or centers where you received your medical care.

We will analyze the genes in your cancer cells (obtained from the sample of your blood) and your normal cells (obtained from the sample of your blood or from your saliva sample if you previously sent one to us). No additional procedures will be required. The results of this analysis will be used to try to develop better ways to treat and prevent cancers.

We will link the results of the gene tests on your cancer cells and normal cells with medical information that has been generated during the course of your treatment. We are asking your permission to obtain a copy of your medical record from places where you have received care for your cancer.

In some cases, a research doctor may contact you to find out if you would be interested in participating in a different or future research study based on information that may have

415 Main Street
Cambridge, MA 02142
T 617-714-7000
www.joincountmein.org

been found in your blood samples.

To allow sharing of information with other researchers, the National Institutes of Health (NIH) and other organizations have developed central data (information) banks that analyze information and collect the results of certain types of genetic studies. These central banks will store your genetic and medical information and provide the information to qualified researchers to do more studies. We will also store your genetic and medical information at the Broad Institute of MIT and Harvard and share your information with other qualified researchers. Therefore, we are asking your permission to share your results with these special banks and other researchers, and have your information used for future research studies, including studies that have not yet been designed, studies involving diseases other than cancer, and/or studies that may be for commercial purposes (such as the development or approval of new drugs). Your information will be sent to central banks and other researchers only with a code number attached. Your name, social security number, and other information that could readily identify you will not be shared with central banks or other researchers. We will never sell your readily identifiable information to anyone under any circumstances.

#### E. <u>How long will I be in this research study?</u>

You will be asked to give a sample of blood after you cosnet to enrolling in this study. We will keep your blood samples and medical records indefinitely until this study is finished unless you inform us that you no longer wish to participate. You may do this at any time. More information about how to stop being in the study is below in paragraph I.

Once the study is finished, any left over blood samples and your medical records will be destroyed.

#### F. <u>What kind of information could be found in this study and will I be able to see it?</u>

The gene tests in this study are being done to add to our knowledge of how genes and other factors affect cancer. This information will be kept confidential and while you will not receive information about your personal results obtained from studying your blood sample, we will provide general results and major discoveries to all participants. We will do this by regularly updating the website that you used to enroll in this study. Furthermore, we will publish important discoveries found through these studies in the scientific literature so that the entire research community can work together to better understand cancer. Your individual data will not be published in a way in which you could be readily identified. Abstracts, which are plain language summaries of the published reports, will be available to you and the general public.

#### G. What are the risks or discomforts of the research study?

There are small risks associated with obtaining the tube of blood. You may experience slight pain and swelling at the site of the blood draw. These complications are rare and should resolve within a few days. If they do not, you should contact your doctor.

There is a small risk that by participating in this study, the gene test results, including the identification of genetic changes in you or your cancer, could be seen by unauthorized individuals. We have tried to minimize this risk by carefully limiting access to the computers that would house your information to the staff of this research study.

#### H. What are the benefits of the research study?

Taking part in this research study may not directly benefit you. By joining this study, you will help us and other researchers understand how to use gene tests to improve the care of patients with cancer in the future. We will provide study participants updates on our project website about key research discoveries made possible by your participation.

#### I. Can I stop being in the research study and what are my rights?

You can stop being in the research study at any time. We will not be able to withdraw all the information that already has been used for research. If you tell us that you want to stop being in the study, we will destroy any remaining blood or DNA samples we have. We will not perform any additional tests on the samples. Additionally, we will not collect any additional medical records and we will destroy the medical records we already have. However, we will keep the results from the tests we did before you stopped being in the study. We will also keep the information we learned from reviewing your medical records before you stopped being in the study, We will not be able to take back the information that

415 Main Street
Cambridge, MA 02142
T 617-714-7000
www.joincountmein.org

already has been used or shared with other researchers, central data banks, or that has been used to carry out related activities such as oversight, or that is needed to ensure quality of the study.

To withdraw your permission, you must do so in writing by contacting the researcher listed below in the section: “Whom do I contact if I have questions about the research study?” If you choose to not participate, or if you are not eligible to participate, or if you withdraw from this research study, this will not affect your present or future care and will not cause any penalty or loss of benefits to which you are otherwise entitled.

#### J. Will I be paid to take part in this research study?

There is no financial compensation for participation in this study.

#### K. What are the costs?

There are no costs to you to participate in this study.

#### L. What happens if I am injured or sick because I took part in this research study?

There is virtually no chance that you will become injured or sick by taking part in this study. There are no plans for this project to pay you or give you other compensation for any injury. You do not give up your legal rights by signing this form. If you think you have been injured as a result of taking part in this research study, please tell the person in charge of this research study as soon as possible. The research doctor’s contact information is listed in this consent form.

#### M. What about confidentiality?

We will take rigorous measures to protect the confidentiality and security of all your information, but we are unable to guarantee complete confidentiality.

When we receive your blood sample, your name, social security number, and other information that could be used to readily identify you will be removed and replaced by a code. If we send your samples to our collaborators for gene testing, the samples will be identified using only this code. The medical records that we receive will be reviewed by our research team to confirm that you are eligible for the study and to obtain information about your medical condition and treatment.

We will store all of your identifiable information related to the study (including your medical records) in locked file cabinets and in password-protected computer files at the Broad Institute and we will limit access to such files. We may share your identifiable information or coded information, as necessary, with regulatory or oversight authorities (such as the Office for Human Research Protections), ethics committees reviewing the conduct of the study, or as otherwise required by law.

When we send the results of the gene tests and your medical information to central data banks or other researchers, they will not contain your name, social security number, or other information that could be used to readily identify you.

The results of this research study or future research studies using the information from this study may be published in research papers or included in presentations that will become part of the scientific literature. You will not be identified in publications or presentations.

#### N. Whom do I contact if I have questions about the research study?

- Nikhil Wagle, MD (Principle Investigator of the Study)
- Corrie Painter, PhD

For questions about your rights as a patient, please contact a representative of the Office for Human Research Studies at (617) 632-3029. This can include questions about your participation in the study, concerns about the study, a research related injury, or if you feel/felt under pressure to enroll in this research study or to continue to participate in this research study. Please keep a copy of this document in case you want to read it again.

#### O. AUTHORIZATION TO USE YOUR HEALTH INFORMATION FOR RESEARCH

**PURPOSES**

Because information about you and your health is personal and private, it generally cannot be used in this research study without your written authorization. Federal law requires that your health care providers and healthcare institutions (hospitals, clinics, doctor's offices) protect the privacy of information that identifies you and relates to your past, present, and future physical and mental health conditions.

If you sign this form, it will provide your health care providers and healthcare institutions the authorization to disclose your protected health information to the Broad Institute for use in this research study. The form is intended to inform you about how your health information will be used or disclosed in the study. Your information will only be used in accordance with this authorization form and the informed consent form and as required or allowed by law. Please read it carefully before signing it.

1. **What personal information about me will be used or shared with others during this research?**
   - Your medical records
   - Your blood sample
   - New health information created from study-related tests and/or questionnaires
2. **Why will protected information about me be used or shared with others?**

   The main reasons include the following:

   - To conduct and oversee the research described earlier in this form;
   - To ensure the research meets legal, institutional, and accreditation requirements;
   - To conduct public health activities (including reporting of adverse events or situations where you or others may be at risk of harm)
3. **Who will use or share protected health information about me?**

   The Broad Institute and its researchers and affiliated research staff will use and/or share your personal health information in connection with this research study.
4. **With whom outside of the Broad Institute may my personal health information be shared?**

415 Main Street
Cambridge, MA 02142
T 617-714-7000
www.joincountmein.org

While all reasonable efforts will be made to protect the confidentiality of your protected health information, it may also be shared with the following entities:

- Federal and state agencies (for example, the Department of Health and Human Services, the Food and Drug Administration, the National Institutes of Health, and/or the Office for Human Research Protections), or other domestic or foreign government bodies if required by law and/or necessary for oversight purposes. A qualified representative of the FDA and the National Cancer Institute may review your medical records.
- Outside individuals or entities that have a need to access this information to perform functions relating to the conduct of this research such as data storage companies.

Some who may receive your personal health information may not have to satisfy the privacy rules and requirements. They, in fact, may share your information with others without your permission.

5. **For how long will protected health information about me be used or shared with others?**

   There is no scheduled date at which your protected health information that is being used or shared for this research will be destroyed, because research is an ongoing process.

6. **Statement of privacy rights:**
   - You have the right to withdraw your permission for the doctors and researchers to use or share your protected health information. We will not be able to withdraw all the information that already has been used or shared with others to carry out related activities such as oversight, or that is needed to ensure quality of the study. To withdraw your permission, you must do so in writing by contacting the researcher listed above in the section: “Whom do I contact if I have questions about the research study?”
   - You have the right to request access to your personal health information that is used or shared during this research and that is related to your treatment or payment for your treatment. To request this information, please contact your doctor who will request this information from the study directors.

415 Main Street
Cambridge, MA 02142
T 617-714-7000
www.joincountmein.org

**P. Participation Information**

If you decide to sign this consent form, we will ask you for information about contacting your physicians and the hospitals that you were treated at for your cancer. We will not disclose details about the results of your participation in this study with any of the individuals that we contact, but rather ask them to provide us with your medical history and your tissue samples.

**Q. Documentation of Consent**

This is what I agree to:

- You can work with me to arrange a sample of blood to be drawn at my physician's office, local clinic, or nearby lab facility.
- You can perform (or collaborate with others to perform) gene tests on the blood sample that I will send you and store the sample until this research study is complete.
- You can request my medical records from my physicians and the hospitals and other places where I received and/or continue to receive my treatment and link results of the gene tests you perform on my blood samples with my medical information from my medical records.
- You can use the results of the gene tests and my medical information for future research studies, including studies that have not yet been designed, studies for diseases other than cancer, and/or studies that may be for commercial purposes.
- You can share the results of the gene tests and my medical information with central data banks (e.g., the NIH) and with other qualified researchers in a manner that does not include my name, social security number, or any other information that could be used to readily identify me, to be used by other qualified researchers to perform future research studies, including studies that have not yet been designed, studies for diseases other than cancer, and studies that may be for commercial purposes.

My full name below indicates:

- I have had enough time to read the consent and think about agreeing to participate in this study;
- I have had all of my questions answered to my satisfaction;

- I am willing to participate in this research study;
- I have been told that my participation is voluntary and if I decide not to participate it will have no impact on my medical care;
- I have been told that if I decide to participate now, I can decide to stop being in the study at any time.
- I acknowledge that a copy of the signed consent form will be sent to my email address.

YOUR CONTACT INFORMATION:

First Name: ________________________________

Last Name: ________________________________

Current Mailing Address: ________________________________

City: ________________ State: ________________ Zip: ________________

Phone: ________________________________

YOUR TREATMENT INFORMATION:

Current Treatments:

Current Treatment Since:

Past Treatments:

415 Main Street
Cambridge, MA 02142
T 617-714-7000
www.joincountmein.org

Full Name

Date

Date of Birth

415 Main Street
Cambridge, MA 02142
T 617-714-7000
www.broadinstitute.org

Thank you very much for your consent to participate in The Metastatic Breast Cancer Project. To proceed with our study, we have asked you for information about:

1. Your contact information, including your current mailing address, so that we can send you a saliva kit
2. The name and contact information for the physician(s) who has/have cared for you throughout your experiences with breast cancer, so we can obtain copies of your medical records
3. The names of the hospitals / institutions where you've had biopsies and surgeries, so we can obtain some of your stored tumor samples

Printed below is the information you have provided to us:

YOUR CONTACT INFORMATION:

First Name: ______________________________

Last Name: ______________________________

Current Mailing Address: ______________________________

City: ________________ State: ________________ Zip: ________________

Phone: ______________________________

YOUR PHYSICIANS' NAMES:

Physician Name: ______________________________

Institution (if any): ______________________________

Address: ______________________________

City: ________________ State: ________________ Zip: ________________

415 Main Street
Cambridge, MA 02142
T 617-714-7000
www.broadinstitute.org

Phone: ______________________________

YOUR HOSPITAL / INSTITUTION NAMES:

Your initial biopsy for breast cancer was performed:

Institution: ______________________________

City: ________________ State: ________________

Any other biopsies or surgeries for your breast cancer (i.e. biopsy, lumpectomy, partial mastectomy, mastectomy) were performed:

Institution: ______________________________

City: ________________ State: ________________

By completing this information, you are agreeing to allow us to contact these physician(s) and hospital(s) / institution(s) to obtain your records.

- I have already read and signed the informed consent document for this study, which describes the use of my personal health information (Section O), and hereby grant permission to Nikhil Wagle, MD, Dana-Farber Cancer Institute, 450 Brookline Ave, Boston, MA, 02215, or a member of the study team to examine copies of my medical records pertaining to my breast cancer diagnosis and treatment, and to obtain tumor tissue for research studies. I acknowledge that a copy of this completed form will be sent to my email address.

________________ ________________
Full Name Date

415 Main Street
Cambridge, MA 02142
T 617-714-7000
www.broadinstitute.org

______________________

Date of Birth

### Join the movement: tell us about yourself

Thank you for providing your contact information The Metastatic Breast Cancer Project is open to patients in the United States or Canada. Please help us understand more about your breast cancer by answering the questions below

As you fill out the questions below your answers will be automatically saved. If you've previously entered information here and want to pick up where you left off, please use the link we sent you via email to return to this page.

If you decide not to complete this form and would like the information entered below to be deleted from the database, now or in the future, you can email info@mbcproject org to request that your responses be removed.

#### About You

Please fill out as much as you can. All questions are optional. You can return at any time with the link sent to you by email.

1. When were you first diagnosed with breast cancer? (i)

   Month ⬍ Year ⬍

2. When were you first diagnosed with metastatic breast cancer (also known as advanced or stage IV breast cancer)? (i)

   Month ⬍ Year ⬍

3. At any time, was your breast cancer found to be hormone receptor positive (HR+, ER+ and/or PR+) (i)
   - ○ Yes
   - ○ No
   - ○ Dont know

4. At any time, was your breast cancer found to be HER2 positive (HER2+)? (i)
   - ○ Yes
   - ○ No
   - ○ Dont know

5. At any time, was your breast cancer found to be triple negative (e.g. NOT ER+ PR+ or HER2+)? (i)
   - ○ Yes
   - ○ No
   - ○ Dont know

6. At any time, were you diagnosed with *inflammatory breast cancer*? (i)
   - ○ Yes
   - ○ No
   - ○ Dont know

7. Since your diagnosis with metastatic breast cancer have you been on any of your cancer therapies for more than 2 years? (i)
   - ○ Yes
   - ○ No
   - ○ Dont know

7a. If YES, please list the cancer therapies you have been on for more than 2 years if you know their names. For a list of commonly used therapies, click here. (i)

Describe treatments

8. Have any of your therapies worked extraordinarily well — made your cancer disappear completely (resulting in no evidence of disease, NED) or resulted in a dramatic reduction in tumor size — for any period of time? (i)
   - ○ Yes
   - ○ No
   - ○ Dont know

8a. If YES, please list the cancer therapies that have worked extraordinarily well, if you know their names. For a list of commonly used therapies, click here. (i)

Describe treatments

9. If you have had an extraordinary response to therapy, tell us more about it. (i)

   Describe response/therapy

10. When was your most recent biopsy of your cancer? (i)

    Month ⬍ Year ⬍

11. Tell us anything else you would like to about yourself or your cancer (i)

12. In what year were you born?

    Year

13. * What country do you live in?

    Select a country ▾

13a. What is your ZIP or postal code?

ZIP code

14. Do you consider yourself Hispanic, Latino/a or Spanish?
    - ○ Yes
    - ○ No
    - ○ Dont know

15. What is your race (select all that apply)?
    - ☐ American Indian or Native American
    - ☐ Japanese
    - ☐ Chinese
    - ☐ Other East Asian
    - ☐ South East Asian or Indian
    - ☐ Black or African American
    - ☐ Native Hawaiian or other Pacific Islander
    - ☐ White
    - ☐ I Prefer Not to Answer
    - ☐ Other

    (other - please specify)

I understand that the information I entered here will be stored in a secure database for research studies conducted by the Metastatic Breast Cancer Project. If I am in the United States or Canada, I agree to be contacted about possibly participating.

I understand that if I do not sign the research consent form to participate in the Metastatic Breast Cancer Project and would like the information I entered above deleted from the database, now or in the future, I can to request that my information be removed.

I understand that if I sign the research consent form and would like to withdraw from the research study I can contact the study team at at any time, although any of my information that has already been entered into the system cannot be withdrawn. My information would be removed from future studies.

SUBMIT



Contact Us:

617 800 1622

Count Me In
415 Main St Cambridge, MA
02142 United States

### Join the movement: tell us about yourself

Thank you for providing your contact information. The Metastatic Breast Cancer Project is open to patients in the United States or Canada. Please help us understand more about your breast cancer by answering the questions below.

As you fill out the questions below, your answers will be automatically saved. If you've previously entered information here and want to pick up where you left off, please use the link we sent you via email to return to this page.

If you decide not to complete this form and would like the information entered below to be deleted from the database, now or in the future, you can to request that your responses be removed.

#### About You

Please fill out as much as you can. All questions are optional. You can return at any time with the link sent to you by email.

1. When were you first diagnosed with breast cancer? (i)

   Month | Year

2. When were you first diagnosed with metastatic breast cancer (also known as advanced or stage IV breast cancer)? (i)

   Month | Year

3. At any time, was your breast cancer found to be hormone receptor positive (HR+, ER+ and/or PR+) (i)
   - ○ Yes
   - ○ No
   - ○ Don't know

4. At any time, was your breast cancer found to be HER2 positive (HER2+)? (i)
   - ○ Yes
   - ○ No
   - ○ Don't know

5. At any time, was your breast cancer found to be triple negative (e.g, NOT ER+,PR+, or HER2+)? (i)
   - ○ Yes
   - ○ No
   - ○ Don't know

6. At any time, were you diagnosed with *inflammatory breast cancer*? (i)
   - ○ Yes
   - ○ No
   - ○ Don't know

7. Since your diagnosis with metastatic breast cancer, have you been on any of your cancer therapies for more than 2 years? (i)
   - ○ Yes
   - ○ No
   - ○ Don't know

7a. If YES, please list the cancer therapies you have been on for more than 2 years, if you know their names. For a list of commonly used therapies, click here. (i)

   Describe treatments

8. Have any of your therapies worked extraordinarily well — made your cancer disappear completely (resulting in no evidence of disease, NED) or resulted in a dramatic reduction in tumor size — for any period of time? (i)
   - ○ Yes
   - ○ No
   - ○ Don't know

8a. If YES, please list the cancer therapies that have worked extraordinarily well, if you know their names. For a list of commonly used therapies, click here. (i)

   Describe treatments

9. If you have had an extraordinary response to therapy, tell us more about it. (i)

   Describe response/therapy

10. When was your most recent biopsy of your cancer? (i)

    Month | Year

11. Tell us anything else you would like to about yourself or your cancer: (i)

12. In what year were you born?

    Year

13. * What country do you live in?

    Select a country ▼

13a. What is your ZIP or postal code?

    ZIP code

14. Do you consider yourself Hispanic, Latino/a or Spanish?
    - ○ Yes
    - ○ No
    - ○ Don't know

15. What is your race (select all that apply)?
    - ☐ American Indian or Native American
    - ☐ Japanese
    - ☐ Chinese
    - ☐ Other East Asian
    - ☐ South East Asian or Indian
    - ☐ Black or African American
    - ☐ Native Hawaiian or other Pacific Islander
    - ☐ White
    - ☐ I Prefer Not to Answer
    - ☐ Other

    (other - please specify)

16. How did you hear about the MBCproject?

I understand that the information I entered here will be stored in a secure database for research studies conducted by the Metastatic Breast Cancer Project. If I am in the United States or Canada, I agree to be contacted about possibly participating.

I understand that if I do not sign the research consent form to participate in the Metastatic Breast Cancer Project and would like the information I entered above deleted from the database, now or in the future, I can to request that my information be removed.

I understand that if I sign the research consent form and would like to withdraw from the research study, I can contact the study team at at any time, although any of my information that has already been entered into the system cannot be withdrawn. My information would be removed from future studies.

SUBMIT



Contact Us:

617-800-1622

Count Me In
415 Main St, Cambridge, MA
02142, United States

Metastatic Breast Cancer Project

Learn More

### Join the movement: tell us about yourself

Thank you for providing your contact information. The Metastatic Breast Cancer Project is open to patients in the United States or Canada. Please help us understand more about your metastatic breast cancer by answering the questions below.

As you fill out the questions below, your answers will be automatically saved. If you've previously entered information here and want to pick up where you left off, please use the link we sent you via email to return to this page.

If you decide not to complete this form and would like the information entered below to be deleted from the database, now or in the future, you can to request that your responses be removed.

#### About You

Please fill out as much as you can. All questions are optional. You can return at any time with the link sent to you by email.

1. When were you first diagnosed with breast cancer?

   Choose month... | Choose year...

2. When were you first diagnosed with metastatic breast cancer (also known as advanced or stage IV breast cancer)?

   Choose month... | Choose year...

3. At any time, was your breast cancer found to be hormone receptor positive (HR+, ER+ and/or PR+)?

   - ○ Yes
   - ○ No
   - ○ I don't know

4. At any time, was your breast cancer found to be HER2 positive (HER2+)?

   - ○ Yes
   - ○ No

- I don't know

5. At any time, was your breast cancer found to be triple negative (e.g, NOT ER+, PR+ or HER2+)?
   - Yes
   - No
   - I don't know

6. At any time, were you diagnosed with *inflammatory breast cancer*?
   - Yes
   - No
   - I don't know

7. Since your diagnosis with metastatic breast cancer, have you been on any of your cancer therapies for more than 2 years?
   - Yes
   - No
   - I don't know

   Please list the cancer therapies you have been on for more than 2 years, if you know their names.

   Choose therapy...

   + Add Another Therapy

8. Have any of your therapies worked extraordinarily well - made your cancer disappear completely (resulting in no evidence of disease, NED) or resulted in a dramatic reduction in tumor size - for any period of time?
   - Yes
   - No
   - I don't know

   Please list the cancer therapies that have worked extraordinarily well, if you know their names.

   Choose therapy...

   + Add Another Therapy

9. If you have had an extraordinary response to therapy, tell us more about it.

   Describe response/therapy

10. When was your most recent biopsy of your cancer?

Choose month... | Choose year...

11. Tell us anything else you would like to about yourself or your cancer.

12. In what year were you born?

Choose year...

13. What country do you live in? *

Choose country...

14. What is your ZIP or postal code?

Zip Code

15. Which categories describe you? Select all that apply. Note, you may select more than one group.

- [x] American Indian or Alaskan Native

  **(For example: Aztec, Blackfeet Tribe, Mayan, Navajo Nation, Native Village of Barrow (Utqiagvik) Inupiat Traditional Government, Nome Eskimo Community, etc.)**

  - [ ] American Indian
  - [ ] Alaska Native
  - [ ] Central or South American Indian
  - [ ] None of these fully describe me

- [x] Asian

  **(For example: Asian Indian, Chinese, Filipino, Japanese, Korean, Vietnamese, etc.)**

  - [ ] Asian Indian
  - [ ] Cambodian
  - [ ] Chinese
  - [ ] Filipino
  - [ ] Hmong
  - [ ] Japanese
  - [ ] Korean
  - [ ] Pakistani
  - [ ] Vietnamese
  - [ ] None of these fully describe me

- [x] Black, African American, or African

(For example: African American, Ethiopian, Haitian, Jamaican, Nigerian, Somali, etc.)

- [ ] African American
- [ ] Barbadian
- [ ] Caribbean
- [ ] Ethiopian
- [ ] Ghanaian
- [ ] Haitian
- [ ] Jamaican
- [ ] Liberian
- [ ] Nigerian
- [ ] Somali
- [ ] South African
- [ ] None of these fully describe me

- [x] Hispanic, Latino, or Spanish

(For example: Colombian, Cuban, Dominican, Mexican or Mexican American, Puerto Rican, Salvadoran, etc.)

- [ ] Colombian
- [ ] Cuban
- [ ] Dominican
- [ ] Ecuadorian
- [ ] Honduran
- [ ] Mexican or Mexican American
- [ ] Puerto Rican
- [ ] Salvadoran
- [ ] Spanish
- [ ] None of these fully describe me

- [x] Middle Eastern or North African

(For example: Algerian, Egyptian, Iranian, Lebanese, Moroccan, Syrian, etc.)

- [ ] Afghan
- [ ] Algerian
- [ ] Egyptian
- [ ] Iranian
- [ ] Iraqi
- [ ] Israeli
- [ ] Lebanese
- [ ] Moroccan

- [ ] Syrian
- [ ] Tunisian
- [ ] None of these fully describe me

- [x] Native Hawaiian or other Pacific Islander

(For example: Chamorro, Fijian, Marshallese, Native Hawaiian, Tongan, etc.)

- [ ] Chamorro
- [ ] Chuukese
- [ ] Fijian
- [ ] Marshallese
- [ ] Native Hawaiian
- [ ] Palauan
- [ ] Samoan
- [ ] Tahitian
- [ ] Tongan
- [ ] None of these fully describe me

- [x] White

(For example: English, European, French, German, Irish, Italian, Polish, etc.)

- [ ] Dutch
- [ ] English
- [ ] European
- [ ] French
- [ ] German
- [ ] Irish
- [ ] Italian
- [ ] Norwegian
- [ ] Polish
- [ ] Scottish
- [ ] Spanish
- [ ] None of these fully describe me

- [x] None of these fully describe me

Please specify

500 characters remaining

- [ ] Prefer not to answer

Since you selected "Hispanic, Latino, or Spanish" in the previous question, we would like to ask some follow up questions about your racial and ethnic identity.

Do you consider yourself to be mixed race, that is belonging to more than one racial group, such as Mestizo, Mulatto or some other mixed race, or not?

- Yes
- No
- I don't know
- Prefer not to answer

Do you consider yourself to be Afro-Latino, Afro-Hispanic, Afro-Caribbean, or not?
(An Afro-Latino or Afro-Hispanic is a Latino or Hispanic with black African ancestry)

- Yes
- No
- I don't know
- Prefer not to answer

Do you consider yourself to be indigenous or Native American
(such as Purepecha, Mixteco, Zapoteco, Nahua, Maya, Tzotzil, Taino, Arawak, Quiche, Aymara, Quechua, Kwichua, Mapuche, Guarani, or some other indigenous or Native American origin), or not?

- Yes
- No
- I don't know
- Prefer not to answer

16. What is your gender identity? Select all that apply

- [ ] Man
- [ ] Woman
- [ ] Transgender
- [ ] Nonbinary
- [ ] A gender not listed here
- [ ] Prefer not to answer

17. What sex were you assigned at birth?

- Male
- Female
- Intersex
- Prefer not to answer

18. How did you hear about the MBCproject?

I understand that the information I entered here will be stored in a secure database for research studies conducted by the Metastatic Breast Cancer Project. If I am in the USA or Canada, I agree to be contacted about possibly participating in the Metastatic Breast Cancer Project.

I understand that if I do not sign the research consent form to participate in the Metastatic Breast Cancer Project and would like the information entered above to be deleted from the database, now or in the future, I can to request that my information be removed.

I understand that if I sign the research consent form and would like to withdraw from the research study, I can contact the study team at at any time, although any of my information that has already been entered into the system cannot be withdrawn. My information would be removed from future studies.

##### Citation information for race & ethnicity questions

Survey question 15 uses some of the language developed for race & ethnicity questions and responses created either as part of the *All of Us* Research Program from the National Institutes of Health or as part of the Pew Research Center 2014 National Survey of Latinos. Below is the citation information for these questions:

Cronin RM, Jerome RN, Mapes B, Andrade R, Johnston R, Ayala J, Schlundt D, Bonnet K, Kripalani S, Goggins K, Wallston KA, Couper MP, Elliott MR, Harris P, Begale M, Munoz F, Lopez-Class M, Cella D, Condon D, AuYoung M, Mazor KM, Mikita S, Manganiello M, Borselli N, Fowler S, Rutter JL, Denny JC, Karlson EW, Ahmedani BK, O'Donnell CJ; Vanderbilt University Medical Center Pilot Team, and the Participant Provided Information Committee. Development of the Initial Surveys for the All of Us Research Program. Epidemiology. 2019 Jul;30(4):597-608. doi: 10.1097/EDE.0000000000001028. PMID: 31045611; PMCID: PMC6548672.

Mapes BM, Foster CS, Kusnoor SV, Epelbaum MI, AuYoung M, Jenkins G, Lopez-Class M, Richardson-Heron D, Elmi A, Surkan K, Cronin RM, Wilkins CH, Pérez-Stable EJ, Dishman E, Denny JC, Rutter JL; All of Us Research Program. Diversity and inclusion for the All of Us research program: A scoping review. PLoS One. 2020 Jul 1;15(7):e0234962. doi: 10.1371/journal.pone.0234962. PMID: 32609747; PMCID: PMC7329113.

"Multiracial in America." Pew Research Center, Washington, D.C. (June 11, 2015). https://www.pewresearch.org/social-trends/2015/06/11/multiracial-in-america

SUBMIT

Contact Us:

617-800-1622

Count Me In
415 Main St, Cambridge, MA
02142, United States

Metastatic Breast Cancer Project

### Follow-up survey #1: Additional details about your cancer & treatments

Please help us understand more about your metastatic breast cancer by answering the six questions below.

As you fill out the questions, your answers will be automatically saved. If you would like to leave the survey and complete it at another time, please use the link we sent you via email to return to this page.

If you would like your information deleted from our database, please let us know by emailing and we will remove your name and email address and the answers to any questions you may have answered.

Questions 1-3 refer to your experiences since your diagnosis with metastatic breast cancer.

1. Please select all of the places in your body where you **currently** have metastatic breast cancer to the best of your knowledge (select all that apply). If you don't have any detectable disease please select No Evidence of Disease (NED).

- ☐ Breast(s)
- ☐ Axillary Lymph Nodes (lymph nodes in the armpit)
- ☐ Lymph Nodes anywhere other than the axilla/armpit
- ☐ Bone (may include ribs, sternum, pelvis, vertebrae, skull)
- ☐ Chest Wall
- ☐ Liver
- ☐ Lung(s)
- ☐ Brain
- ☐ Pleural Effusion (fluid in the lung)
- ☐ Ascites (fluid in the abdomen)
- ☐ Skin
- ☐ Ovary
- ☐ Other
- ☐ No Evidence of Disease (NED)
- ☐ I don't know

2. When you were **first diagnosed** with **metastatic** breast cancer, where were all of the places in your body that it was detected (select all that apply)?

- ☐ Breast(s)
- ☐ Axillary Lymph Nodes (lymph nodes in the armpit)
- ☐ Lymph Nodes anywhere other than the axilla/armpit
- ☐ Bone (may include ribs, sternum, pelvis, vertebrae, skull)
- ☐ Chest Wall
- ☐ Liver
- ☐ Lung(s)
- ☐ Brain
- ☐ Pleural Effusion (fluid in the lung)
- ☐ Ascites (fluid in the abdomen)

- [ ] [illegible]
- [ ] Skin
- [ ] Ovary
- [x] Other

Please provide details

- [ ] I don't know

3. Please select all of the places in your body that metastatic breast cancer has been found **at any time** (select all that apply).

- [ ] Breast(s)
- [ ] Axillary Lymph Nodes (lymph nodes in the armpit)
- [ ] Lymph Nodes anywhere other than the axilla/armpit
- [ ] Bone (may include ribs, sternum, pelvis, vertebrae, skull)
- [ ] Chest Wall
- [ ] Liver
- [ ] Lung(s)
- [ ] Brain
- [ ] Pleural Effusion (fluid in the lung)
- [ ] Ascites (fluid in the abdomen)
- [ ] Skin
- [ ] Ovary
- [ ] Other
- [ ] I don't know

4. Was your breast cancer identified as any of the following **at any time** (select all that apply)?

- [ ] Invasive Ductal Carcinoma (IDC)
- [ ] Invasive Lobular Carcinoma (ILC)
- [ ] Mixed Invasive Ductal/Lobular Carcinoma (IDLC)
- [x] Other Rare Subtypes (please specify in the following question)
- [ ] I don't know

Please choose the other rare subtypes (select all that apply)

- [ ] Mucinous (Colloid)
- [ ] Metaplastic
- [ ] Tubular
- [ ] Medullary
- [ ] Papillary
- [ ] Sarcoma
- [ ] Lymphoma
- [ ] Phyllodes
- [ ] Other
- [ ] I don't know

Please fill out the following treatment information.
You may have filled out previous forms for the
[illegible]

MBCproject, but we would like to make sure that we have your most current information.

5. Are you currently receiving any medications/chemotherapies for treatment of your metastatic breast cancer?

- [x] Yes
- [ ] No
- [ ] I don't know

Please list all medications/chemotherapies that you are currently receiving for treatment of your metastatic breast cancer (list all that apply). If known, please enter the start date of these medications. If you are receiving multiple medications/chemotherapies at the same time, please enter each individually.

- [ ] I don't know the names of the medications

Start typing a Medication/Therapy...

Start Date

Choose month... | Choose year...

- [ ] This was part of a clinical trial

+ ADD ANOTHER MEDICATION/CHEMOTHERAPY

6. Have you received any other medications/chemotherapies in the past for treatment of your metastatic breast cancer?

- [x] Yes
- [ ] No
- [ ] I don't know

Please list all other medications/chemotherapies that you have received in the past for your metastatic breast cancer (list all that apply). If known, please enter the start and stop dates of these medications. If you received multiple medications/chemotherapies at the same time, please enter each individually.

- [ ] I don't know the names of the medications

Start typing a Medication/Therapy...

Start Date

Choose month... | Choose year...

End Date

Choose month... | Choose year...

- [ ] This was part of a clinical trial

+ ADD ANOTHER MEDICATION/CHEMOTHERAPY

SUBMIT

**Nikhil Wagle, MD**
Broad Institute of MIT & Harvard
MBCProject, Cancer Program
415 Main St, 105B-[546]
Cambridge, MA 02142
P: 617-714-8836
F: 617-395-2631
E:

### FAX

**To:**

**Fax:**

**From:** Nikhil Wagle, MD

**Phone Number:** 617-714-8836

**Fax Number:** 617-395-2631

**Number of Pages Including Cover Page:** #

**Date:**

**NOTES**: The following materials are enclosed for a patient enrolled on DFCI Protocol 15-057, and we are requesting the patient's oncology medical records as part of the study procedure. The following items are enclosed as part of our request.

1. Medical Record Request Letter (i.e., outline of requested materials)
2. Electronically-signed patient consent form
3. Electronically-signed Medical Record Release Form
4. Latest IRB approval memo

**Nikhil Wagle, MD**
**Broad Institute**
**MBCProject, Cancer Program**
**415 Main Street, 105B-[546]**
**Cambridge, MA 02142**

**IMPORTANT:** Please note that consent & release forms **DO NOT EXPIRE**. Participants sign consent and complete the medical record release form only once when they are enrolled. Participants are never re-consented for this minimal risk study and consent forms remain valid with annual study renewal per IRB approval (included). Please call Tania Hernandez, the study's Clinical Research Coordinator, at 617-714-8836 if you have questions.

**CONFIDENTIAL**

The documents accompanying this fax transmission may contain confidential patient information belonging to the sender that is legally privileged. This authorized recipient of this information is prohibited from disclosing this information to any other party. If you have received this transmission in error, please notify the sender immediately. Thank you.

Dear

We are writing to request the medical records for the following patient:

**Patient:**

**DOB:**

We are requesting the following documents from the patients' medical record:

**From:**

**Through:**

- ☐ All clinic notes from treating providers, including medical oncologists, residents, fellows, radiation oncologists, surgeons, nurse practitioners, etc.
- ☐ Breast cancer treatment data (including radiation, chemotherapy and hormonal therapy)
- ☐ Pathology reports
- ☐ Operative reports
- ☐ Referrals
- ☐ MD to MD exchange
- ☐ Genetic testing reports
- ☐ DICOM format images

We are reviewing this patient's medical records as part of their participation in **The Metastatic Breast Cancer Project** at Dana-Farber Cancer Institute and the Broad Institute of MIT and Harvard. If you have any questions, please feel free to contact Clinical Research Coordinator Tania Hernandez at 617-714-8836.

Thank you very much for your help with our study!

CONFIDENTIAL

The documents accompanying this fax transmission may contain confidential patient information belonging to the sender that is legally privileged. This authorized recipient of this information is prohibited from disclosing this information to any other party. If you have received this transmission in error, please notify the sender immediately. Thank you.

**Nikhil Wagle, MD**
Broad Institute
MBCProject, Cancer Program
105 Broadway, Room 554
Cambridge, MA 02142
P: 617-714-8836 F: 617-395-2631
E:

### Fax

To: Institution ABC
Re: **Pathology Report Request**

To: Pathology Department
Fax Number: XXX-XXX-XXXX #
Pages including cover page: X
Phone Number: XXX-XXX-XXXX
Date: XX/XX/XXXX

From Fax Number: 617-395-2631

The patient detailed below is enrolled on DFCI research protocol #15-057B, and we are requesting the patient's pathology report from the procedure indicated on the following page as part of study procedures.

---

The report can be faxed to the above number or mailed to the above address. The research study will pay for any administrative fees. Please feel free to contact Clinical Research Coordinator Tania Hernandez at 617-714-8836 with any questions. The patient's information is below:

**Confidential**

The documents accompanying this fax transmission may contain confidential patient information belonging to the sender that is legally privileged. This authorized recipient of this information is prohibited from disclosing this information to any other party. If you have received this transmission in error, please notify the sender immediately. Thank you.

**Patient: FIRST NAME LAST NAME**
DOB: XX/XX/XXXX

1) **PROCEDURE NAME, PROCEDURE DATE, ACCESSION #, ANY OTHER RELEVANT INFO**

Thank you!

**Nikhil Wagle, MD**
Broad Institute of MIT & Harvard
MBCProject, Cancer Program
320 Charles St, 105B-[546]
Cambridge, MA 02142
P: 617-714-8836
F: 617-395-2631
E:

DANA-FARBER CANCER INSTITUTE

BROAD INSTITUTE

Count Me In

### FAX

**To:**

**Re: Tissue Request**

**ATTN:** Pathology Dept

**Phone Number:**

**Fax Number:**

**From Fax Number:** 617-395-2631

**Number of Pages Including Cover Page:** #

**Date:**

The patient detailed below is enrolled in research protocol #15-057B at DFCI, and I am requesting the patient's tissue(s) that needs to be analyzed. Please send:

- 25 (5- micron thick) sections on unstained slides AND 1 H&E slide containing invasive carcinoma from the procedure listed on the following page.
  - Please air dry on uncharged slides and mount all tissue in the same direction.
  - Unstained slides and H&Es will not be returned.
  - **Please do not exhaust the tissue available when sending.**

If the above is not able to be sent, please send one of the following alternative options:

- Option 1: Minimum of 5 unstained slides **AND** 1 H&E slide
  - Unstained slides and H&Es will not be returned.
- Option 2: Minimum of 6 unstained slides
  - Unstained slides will not be returned.
- Option 3: Alternatively, you may send tissue blocks that we will cut at our pathology facility.
  - All blocks will be returned in a timely manner.

**Please include a pathology report with the sample(s).**

CONFIDENTIAL

The documents accompanying this fax transmission may contain confidential patient information belonging to the sender that is legally privileged. This authorized recipient of this information is prohibited from disclosing this information to any other party. If you have received this transmission in error, please notify the sender immediately. Thank you.

The tissue can be mailed to me at the above address. The research study will pay for any tissue cutting or administrative fees.

IMPORTANT: Please note that consent & release forms DO NOT EXPIRE. Participants sign consent and complete the medical record release form only once when they are enrolled. Participants are never re-consented for this minimal risk study and consent forms remain valid with annual study renewal per IRB approval (included). Please feel free to contact the project's Clinical Research Coordinator, Tania Hernandez, at 617-714-8836 or email us at with any questions. The patient's information is below:

- **Patient:**
- **DOB:**

**1)**

**Acc#:**

**Date of service:**

**2)**

**Acc#:**

**Date of service:**

**3)**

**Acc#:**

**Date of service:**

**4)**

**Acc#:**

**Date of service:**

**Thank you!**

CONFIDENTIAL

The documents accompanying this fax transmission may contain confidential patient information belonging to the sender that is legally privileged. This authorized recipient of this information is prohibited from disclosing this information to any other party. If you have received this transmission in error, please notify the sender immediately. Thank you.
